# Measuring implementation of clinical guidelines through the COVID-19 pandemic, using linked national health records: a national study of type 2 diabetes in England

**DOI:** 10.64898/2026.08.07.26359950

**Authors:** Pardis Biglarbeigi, Caroline Dale, Andrew Lambarth, Andrew Mason, Rohan Takhar, Giulia Ballabio, John Minshull, Mamas A Mamas, Christopher Tomlinson, Shaun Rowark, Gerry Rayman, Ewan R. Pearson, Kamlesh Khunti, Naveed Sattar, Reecha Sofat, the CVD-COVID-UK/CVD-IMPACT Consortium

## Abstract

**Objectives:** To examine the conformance to type 2 diabetes NICE guidelines across cardiovascular risk strata; and to quantify geographical variation in treatment pathways following the COVID-19 pandemic, encompassing guideline changes.

**Design:** We carried out a retrospective observational study using linked electronic health records across England. Process mining, a data driven method that can reconstruct clinical treatment pathways, was applied to map 12-month treatment trajectories after treatment initiation. Conformance with NICE NG28 (2022) was quantified using a structural similarity index. Further, behavioural and entropy-based similarity (capturing treatment variability and complexity) measures were used to assess sequencing and heterogeneity of treatment.

**Setting:** Primary and secondary care in England datasets within the National Health Service England Secure Data Environment (NHSE SDE), analysed first at national level and then across 42 Integrated Care Boards (ICBs) which are the devolved health care geographical delivery regions in England.

**Participants:** 822,650 individuals with newly diagnosed T2DM between 1-February-2022 and 1-November-2025, stratified into low cardiovascular risk (LR-C; QRISK3<10), high risk (HR-C; QRISK3>=10 or receiving statins/blood pressure lowering treatment), and established cardiovascular disease (eCVD-C). Participants were followed for 12 months after first dispensed glucose lowering therapy.

**Main outcome measure:** First line therapy, treatment intensification and switching within 12 months; change in glycated haemoglobin (HbA1c); quantified conformance to NICE recommended pathways; and regional variation in broader similarity measures.

**Results:** Metformin monotherapy was the dominant initiation strategy in LR-C and HR-C cohorts (92.4% and 90.2%, respectively), whereas eCVD-C showed lower uptake of metformin (68.9%) and higher uptake of SGLT2 inhibitors (26.3%). Intensification from metformin to combination therapy was infrequent across all cohorts (<1%), although HR-C demonstrated the highest treatment transitions and switching behaviour. Dispensed SGLT2 inhibitor use was nearly threefold higher in eCVD-C (26.7%) than in LR-C (9.0%) or HR-C (10.7%). Overall, conformance to NICE-recommended pathways remained modest nationally, particularly in LR-C and HR-C. Across 42 ICBs, substantial regional heterogeneity in treatment pathways and guideline conformance was observed, with conformance ranging from 0.29 to 1.00 in LR-C pathways, 0.40 to 1.00 in HR-C pathways, and 0.54 to 0.92 in eCVD pathways.

**Conclusion:** National T2DM treatment pathways for post-pandemic showed higher alignment to NICE guidelines in eCVD-C compared to the other risk groups, with ongoing gaps and large regional variations in other risk groups. Process mining offers a scalable approach to monitor implementation of guideline recommended care that could support learning health systems. Using T2DM during and post COVID-19 pandemic as a case study, this work demonstrates how these methods can assess the use of existing and innovative therapies, identify gaps and guide future adoption to ensure recommended treatments reach the right patient groups.

## 1. Introduction

Healthcare systems increasingly aim to improve quality and consistency of care by using routinely collected data to evaluate clinical pathways, identify variation and support evidence-based practice. In England, the Getting it Right First Time (GIRFT) programme[1] exemplifies this approach, combining population-level analysis with clinician feedback to review service delivery and promote best practice. For best practice in pharmacological intervention, GIRFT works to ensure that treatment guidelines outlined by National Institute for Health and Care Excellence (NICE) are adhered to. The COVID-19 pandemic caused widespread disruption to healthcare delivery, creating a natural experiment to examine the resilience of clinical pathways, including appropriate preventative pharmacological management. We have previously demonstrated the indirect effects of the COVID-19 pandemic on cardiovascular (CVD) prevention, through a medicine’s lens[2], including blood pressure and lipid lowering medications. In Type 1 and Type 2 Diabetes Mellitus (T1DM, T2DM), lockdowns led to a substantial reduction in the overall use of medicines to treat cardiovascular risk factors[2].

Here, we examine how type 2 diabetes mellitus (T2DM) was managed across England following the COVID-19 pandemic, and in the context of the February 2022 update to NICE guideline NG28, introduced while health services were still recovering from the pandemic [3], [4],[5]. T2DM is a high-burden condition associated with increased risk of myocardial infarction, stroke, heart failure, and premature mortality; early optimisation of treatment is essential, as delays in intensification are linked to poorer outcomes [6]. However, risk is heterogeneous, and treatment benefits vary across patient groups[7]. Reflecting this, the 2022 NICE T2DM update moved beyond glycaemic control to prioritise therapies with proven cardiovascular (CVD) and chronic kidney disease (CKD) benefits, recommending the preferential use of sodium-glucose co-transporter 2 inhibitors (SGLT2i) and glucagon-like peptide 1 receptor agonists (GLP-1RA) in individuals with established or high CVD risk[8]. These recommendations were implemented during a period of ongoing service disruption in the wake of the pandemic recovery, making their uptake and alignment with practice uncertain.

In this context, ensuring that patients receive the right treatment at the right time, coined “getting it right first time”, is a core principle of modern diabetes care. Using process mining [9], [10] of linked electronic health records, we evaluated how first-line therapy initiation and subsequent treatment intensification evolved during and after the pandemic, and the extent to which practice in post COVID-19 aligned with NICE recommendations (NG28, 2022) across CVD risk groups and geographic regions in England. Our objectives were threefold: to assess adherence to the 2022 NICE NG28 guideline [8]; to examine patterns of treatment intensification among individuals with low risk, high risk, and established CVD; and to evaluate regional variation in care in the context of guideline change instituted in the wake of COVID-19 when services were still disrupted. Moreover, the methods described here can be applied across guidance where clinical pathways require treatment intensification. The T2DM NICE guidance has been updated subsequent to the post-Covid-19 period described here, supported by an expanding set of technology appraisals. As treatment pathways are becoming increasingly complex, there is a need for robust methods, such as process mining, to assess implementation, understand variation, and support NICE, in improving uptake of effective therapies and reducing unwarranted variation in outcomes.

## 2. Methods

### Dataset

We conducted a nationwide retrospective observational study using three linked population-level NHS data sources within NHS England (NHSE) Secure Data Environment (SDE)[11], accessed through the British Heart Foundation (BHF) Data Science Centre’s CVD-COVID-UK/COVID-IMPACT Consortium. The linked datasets used include Hospital Episode Statistics for Admitted Patient Care (HES-APC), General Practice Extraction Service Extract (GPES) for Pandemic Planning and Research (GDPPR), and NHS Business Service Authority (NHSBSA) medicine dispensing data, which capture electronic and paper prescriptions submitted monthly for reimbursement in primary care[12]. Analyses were conducted on a monthly timescale, as dispensing data are available only per calendar-month, with all records assigned to the first day of the corresponding reimbursement month. In the absence of exact dispensing dates, the reimbursement submission date was used as a proxy for the dispensing date. Data were linked for individual patients via a pseudo-identifier [11].

### Study Population

The study considered three temporal periods, preDCOVID-19 (1-Oct-2018 – 31-March-2020), COVID-19 (1-April-2020 – 31-January-2022), and postDCOVID-19 (1-Feb-2022 - 1-Nov-2025), to capture changes in dispensing patterns. However, given the substantial updates to clinical guidelines in the postDCOVID-19 period, with the release of NICE NG28 (February 2022), the conformance assessment and primary analyses were restricted to the postDCOVID-19 period.

Accordingly, the conformance study population comprised individuals with a new diagnosis of T2DM, excluding gestational diabetes mellitus (GDM), pre-existing type 2 diabetes mellitus in pregnancy, T1DM and those at risk of diabetes, from 1-Feb-2022 to 1-Nov-2025. Individuals were considered from their first recorded dispensed T2DM drug in NHSBSA (t_0_) (Supplementary Methods 1.1), and T2DM diagnosis in HES or GDPPR, using T2DM phenotyping code lists[13]. Individuals were followed up for 12 months after their first recorded T2DM drug as treatment initiation (t_0_), thus constructing a standardised 12-month observation window for the cohort.

NICE NG28 2022 guidelines specify intensification and or switching treatments across T2DM drug classes with recommendations based on CVD risk categories and kidney function[8]. Accordingly, the study population was divided into three groups: i) a cohort with chronic heart failure or established atherosclerotic CVD (eCVD-C), (Supplementary Methods 1.2), ii) a high risk of CVD cohort (HR-C), defined by a QRISK3 score of ≥ 10% [14] , and iii) the remainder of the population who are at low risk, (LR-C). In addition, individuals who were in the LR-C but receiving statins or blood pressure lowering medication, were included in the HR-C group since these medications suggest a clinical risk of CVD (agreed through consensus of contributing authors and specialists in T2DM care). It is important to note that CKD stages 3-5 are already considered in the QRISK3 score calculation. However, to understand if QRISK3 is appropriately handing these cases, we verified how individuals with CKD were classified and found only 81/882k (∼0.009%) to be classified in the LR-C group, with a relatively young mean age of 42.64 (±8.81); therefore, we decided not to re-classify these individuals into the HR-C group (again agreed through consensus). Calculation of QRISK3 score and codes to define the subgroups are provided in Supplementary Methods 1.3.

### Type 2 diabetes pharmacological therapies and data processing

T2DM medications were identified using the British National Formulary (BNF) Chapter 6 Section 1 (BNF Chapter 6: Endocrine System section 6.1: “Drugs used in diabetes” (Supplementary Methods 1.1). Drug classes were mapped to current NICE guidelines [8]. To address discontinuities in NHSBSA prescription records due to bulk dispensing (e.g. monthly, 2-month and 3-month prescriptions) and medication cessation (i.e. with no prescription for >3m), a 3-month moving average of prescribed quantities was applied to produce a consistent, continuous time series of medication use, detailed in Supplementary Methods 1.4.

### HbA1c data processing

HbA1c measurements were used to assess glycaemic response to treatment. For each individual, HbA1c values recorded up to 6 months before treatment initiation (*t*_0_) were identified. As HbA1c is not measured at regular intervals, missing months were imputed using the most recent measurement (last observation carried forward). For each treatment transition or intensification, the percentage change in HbA1c between consecutive measurements was calculated, and the mean change across the study population was used to summarise the glycaemic response. These summary statistics were incorporated into process maps to contextualise treatment pathways with corresponding changes in glycaemic control (Supplementary Methods 1.5).

### Process Mining

We applied process mining [9], [10] , a data-driven method for analysing sequences of time stamped events, to identify and visualise longitudinal trajectories of drugs used to treat T2DM. Using the timestamped event logs of EHRs to reconstruct and analyse clinical care pathways, we mapped drug utilisation patterns and treatment pathways for each cardiovascular risk group, during the 12 months following initiation of T2DM therapy.

For the analyses, in a pre-processing step, the 10-year risk of CVD amongst individuals without prevalent CVD, aged 25 to 84, was calculated using QRISK3 algorithm [14] at the time of onset of T2DM (*t*_0_). We then used Python’s Process Mining library (pm4py) [15] to generate process maps that represent the observed sequences of drug utilisation events across the study population. To enhance interpretability and reduce visual complexity, the process maps were optimised using frequency-based filtering to remove infrequent drug transitions and to emphasise the most representative drug utilisation pathways. Process maps were validated by comparing them to actual treatment sequences recorded in EHR data and their performance was measured using the F-score, which balances fitness (how well the map reproduces observed data) and precision (how well it avoids generating unobserved sequences) [16]. The F-score ranges from 0 to 1, with higher values indicating better performance. This standard process mining approach supports transparent and reproducible map selection. More details on applying process mining are detailed in Supplementary Methods 1.6.

Process maps were visualised inspected using directly flow graphs (DFGs), where nodes represent treatments annotated with mean HbA1c during that therapy, and edges show transition probabilities along with percentage changes in mean HbA1c between treatments, reflecting the difference between the therapy-period averages of the prior and subsequent therapies. To support comparison and clinical interpretation, three complementary similarity measures were calculated to compare treatment pathways.

1. **Structural similarity** quantifies the degree to which two pathways share the same therapeutic options and transitions. Low values indicate differences in observed medications or combinations.
2. **Behavioural similarity** assesses the relative frequency and sequencing of treatment transitions, low values reflect differences in how clinicians progress patients through therapy (e.g., probabilities of intensification or switching) even when the same medicines are present, thereby revealing real-world prescribing strategies and potential gaps in care standardisation.
3. **Entropy-based similarity** captures the overall variability and predictability of prescribing patterns. Lower values denote greater dispersion in therapeutic choices and a higher proportion of low-frequency or inconsistent transitions, identifying areas of non-uniform clinical decision-making.

Together, these three measures provide a clinically interpretable framework to compare treatment pathways, highlighting both differences in medication selection and patterns of therapeutic management across patient subgroups, more details in Supplementary Methods 1.7, with clinical interpretations presented in Table S4.

Given the objectives of this study, analyses primarily focused on the structural similarity index. This property enables direct comparison of observed therapeutic sequences with guideline-defined pathways and therefore supports assessment of conformance to NICE recommendations (defined in Supplementary Methods 1.8). Structural similarity was used to quantify the degree of alignment between real-world process maps and NICE guidance. Conformance to NICE guidelines is defined on a scale of [0, 1], where a value of 1 indicates perfect alignment with NICE-recommended pathways and lower values indicates greater deviation from recommended care.

Given that the reported NICE guideline conformance scores are based on structural similarity between the observed and guideline-defined pathways, these scores are deterministic measures derived from the DFGs and, as such, do not include formal statistical uncertainty. To complement this, we also assessed pathway differences using behavioural and entropy-based measures, which capture variability and heterogeneity in treatment sequences and allow comparison of distributions across subgroups. Behavioural and entropy-based similarity indices incorporate information on the frequency and probability of transitions and prescribing patterns, which are not specified within NICE guidelines and therefore cannot be directly compared to guideline recommendations. All three similarity measures were calculated to compare treatment pathways across ICBs with comparison to one reference ICB, that shows the highest conformance to NICE guidelines.

This analysis was performed according to a prespecified analysis plan published on GitHub, along with the phenotyping and analysis code (https://github.com/BHFDSC/CCU014_05).

## 3. Results

We examined temporal trends in T2DM drug dispensing across the pre-, during, and post-COVID-19 periods, stratified by cardiovascular risk groups (LR-C, HR-C, eCVD-C). Total percentage of dispensed T2DM drug categories evolved substantially over time, with a notable increase in the uptake of newer medications, notably SGLT2 inhibitors, in the post-COVID period, particularly across higher-risk groups (Figure 1). Accordingly, the proportion of all dispensed SGLT2 inhibitors in post COVID-19 was nearly 2.5-3 higher in the eCVD-C (26.7%) compared to LR-C (9.0%) or HR-C (10.73%) group. These trends likely reflect both the disruption to healthcare delivery during the pandemic and subsequent changes in clinical guidance and practice following the release of updated recommendations. Given these observed shifts and the introduction of NICE NG28 (2022), we mainly focused on the postDCOVID period to evaluate guideline-concordant care. The treatment pathways for pre-COVID-19 and during COVID-19 periods are presented in Supplementary Results Figures S1, and S2, respectively.

**Figure 1:**
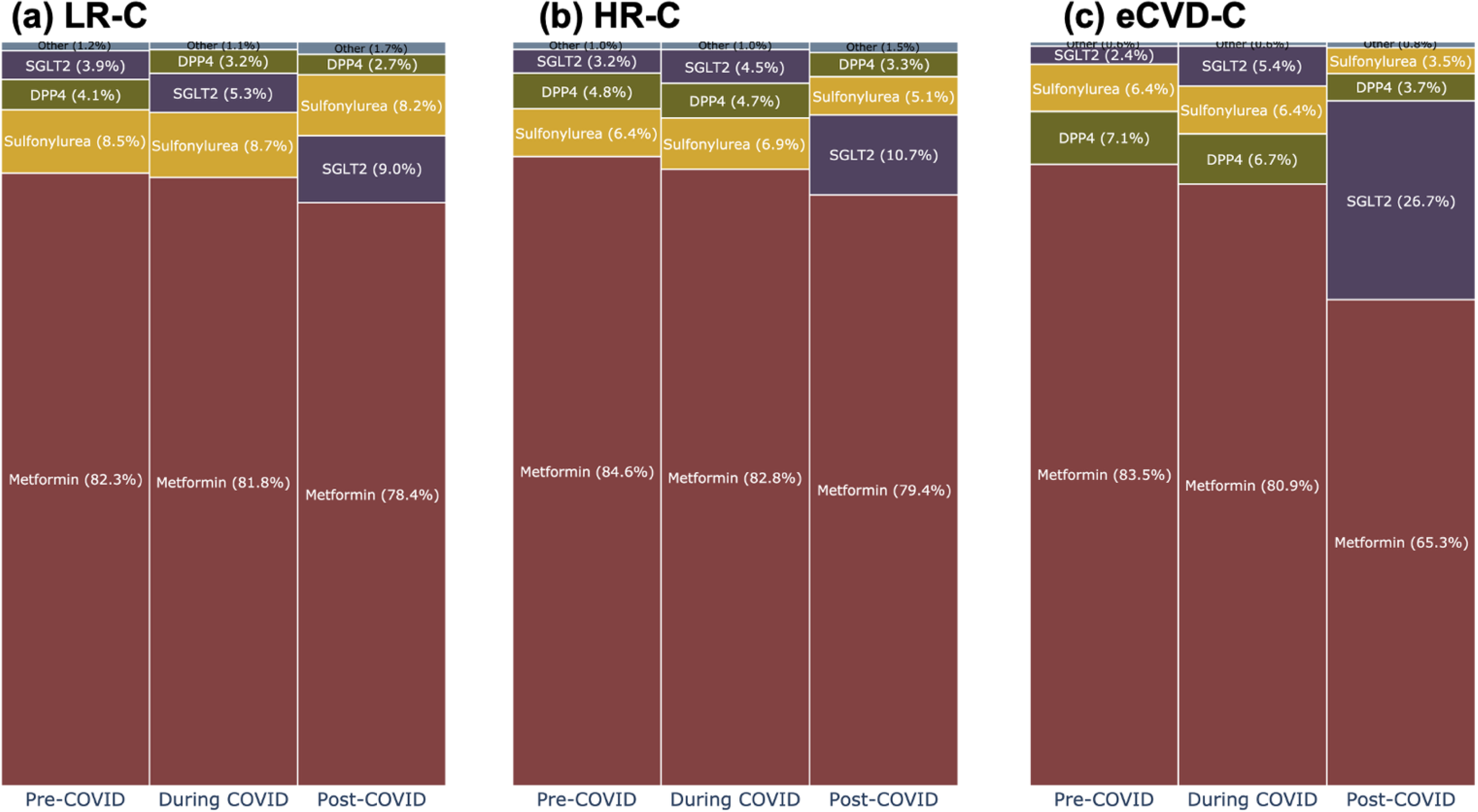
Total percentage of dispensed T2DM drug categories for (a) Low risk – LR-C, (b) High risk – HR-C, and (c) established CVD -eCVD-C cohorts, during different phases of COVID-19.

We reproduced NICE NG28 (2022) T2DM treatment recommendations [8] as a DFG (Supplementary Results Figure S3) stratifying patients into the three broad CVD risk cohorts (LR-C, HR-C, e-CVD-C). According to NICE guidelines, metformin is recommended as first line therapy, unless contraindicated, with early initiation of SGLT2 inhibitors for those with high CVD risk or established CVD.

### National level analysis

A total of 822,650 individuals met inclusion criteria; their characteristics are summarised in Table 1. Optimal process maps for LR-C, eCVD-C, and HR-C demonstrate high-performance scores for all risk groups (with F-scores of 0.97, 0.98, and 0.98, respectively, Figure 2), indicating strong alignment between mapped and observed behaviours.

**Figure 2:**
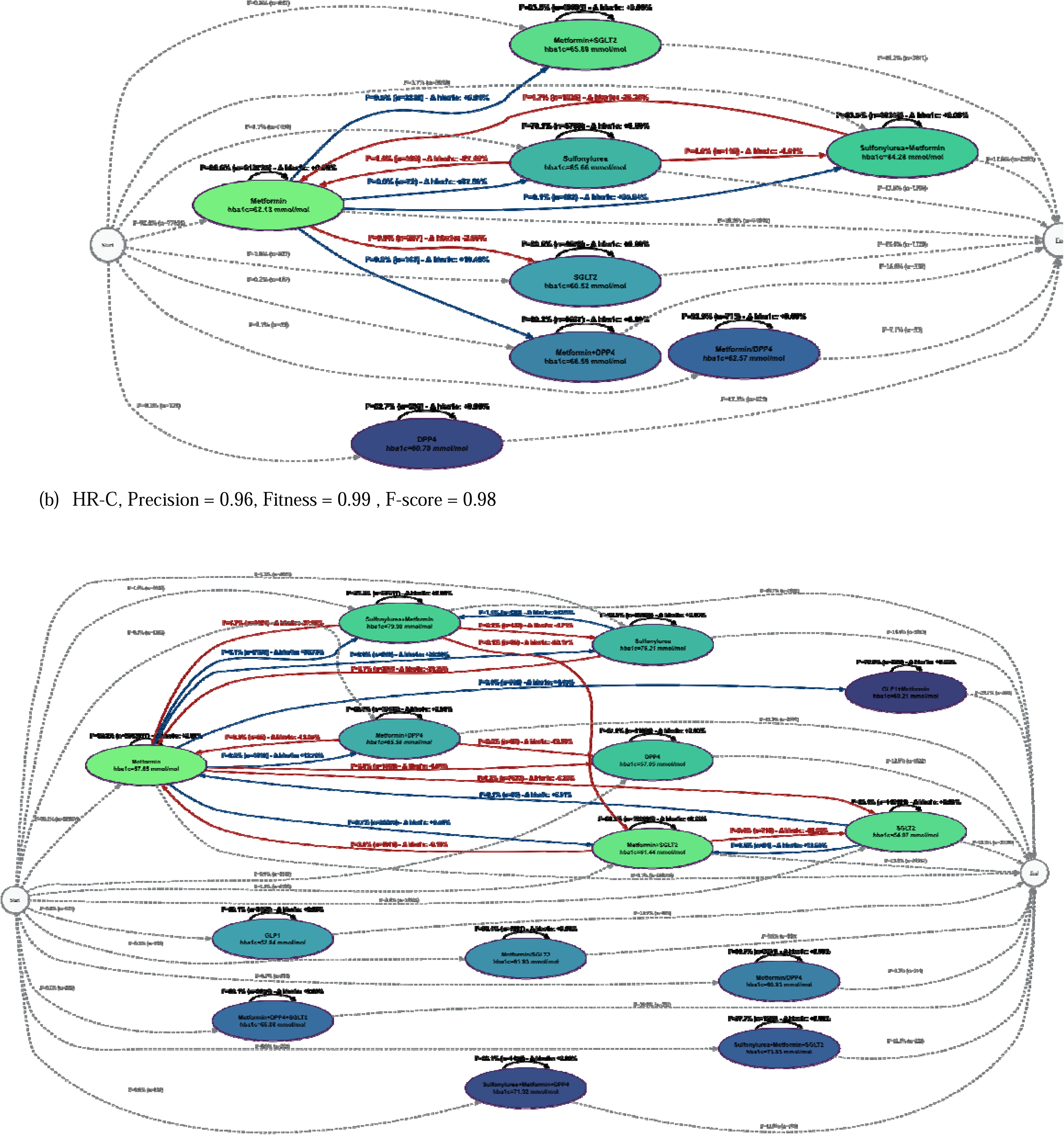
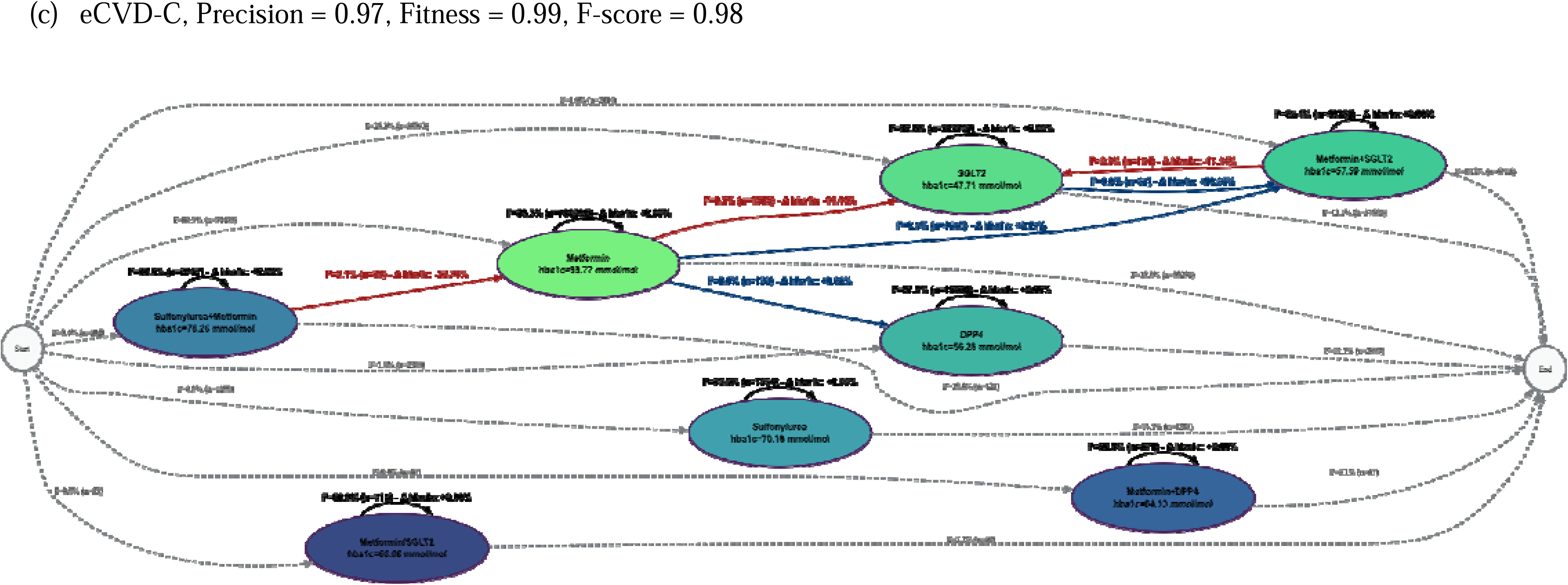
First-line treatment and further interventions for patients with T2DM shown as optimal process maps in the format of Directly-Follows Graph (DFG), in (a) low risk of CVD cohort (LR-C), (b) high risk of CVD cohort (HR-C), and (c) patients with established CVD (eCVD-C).

**Table 1:** The count and age range of patients with T2DM for the three cardiovascular risk groups, according to NICE NG28 2022 guidelines [8], in England.

|  |  | LR-C | HR-C | eCVD-C |
| --- | --- | --- | --- | --- |
| **Population**<br>Feb 2022 – Nov<br>2025 | Count (% female) | 111,215 (47.96%) | 543,575 (43.28%) | 167,860 (33.57%) |
| | Mean Age (±std) | 42.38 (±8.35) | 59.47 (±11.44) | 67.19 (±10.63) |
|  | Mean QRISK3 | 5.11 | 18.74 | n/a |

Taking each of the strata in turn:

- In LR-C, _∼_ 92.4% initiated metformin monotherapy, of whom 99.4% remained on metformin throughout the following 12 months. For those initiating metformin, intensification was rare (*<* 1%) and was most often with addition of SGLT2 inhibitor (0.5%), sulfonylurea (*<* 0.1%) or DPP-4 (*<* 0.01%). Intensification was associated with rising HbA1c. Non-metformin monotherapy starting regimes were observed in _∼_7.6%, with the alternative monotherapies being sulfonylureas (1.7%), SGLT2 (1.0%), DPP4 inhibitors (0.1%), or fixed dose combinations (0.1%). Dual initial therapy occurred in _∼_ 4.7% and was accounted for by metformin plus either SGLT2 (0.8%), DPP4 (0.2%), or sulfonylurea (3.7%). Those patients on dual therapy of sulfonylurea + metformin had the highest probability (4.7%) to transition to single therapy of metformin (Figure 2.a).
- In HR-C, approximately 90.2% of patients initiated with metformin monotherapy. Intensification among those initiated on metformin again occurred with low probability and was most commonly addition of an SGLT2 inhibitor (0.7%), a DPP-4 inhibitor (<0.01%), or a sulfonylurea (0.1%). A small proportion (0.2%) transitioned to SGLT2 monotherapy. Non-metformin monotherapy starting regimens were observed in approximately 9.8% of the cohort; alternative monotherapies included SGLT2 inhibitors (3.6%), sulfonylureas (1.2%), DPP-4 inhibitors (0.9%), and GLP1-RA (0.2%). Dual initial therapy accounted for approximately 3.6% of initiations and comprised metformin + SGLT2 (1.4%), metformin + DPP-4 (0.3), or metformin + sulfonylurea (1.9%). Other drug initiations such as fixed dose treatments, or triple therapy occurred with low probability (0.3%). Patients on the sulfonylurea + metformin dual regimen had notably elevated HbA1c (79.98 mmol/mol) and demonstrated a 5% probability of transitioning to alternative regimens, including a large subset (4.7%) reverting to metformin monotherapy. GLP1-RA monotherapy, though rare (0.2%), was associated with the lowest mean HbA1c of 52.94 mmol/mol across all initiated regimens (Figure 2.b). Overall, HR-C showed the highest transitions between treatments compared to LR-C and eCVD-C.
- In the eCVD-C, a much lower proportion of initiations (68.9%) were on metformin monotherapy, with 26.3% starting directly on SLGT2 inhibitors. While initial treatment with an SGLT2 was far more common, those who started on metformin had a low probability of intensifying with SGLT2 (0.5%) or switching (e.g. to DPP-4 or SGLT2 _∼_ 0.2%). Besides metformin and SGLT2, the other 4.8% of this cohort initiated treatment with DPP4 (1.9%), sulfonylurea (0.9%), or dual therapy (metformin + sulfonylurea (0.4%), metformin + SGLT2 (1.6%), or metformin + DPP4 (<0.01%)). There was also a very low probability of initiation with fixed dose combination. Those on dual therapy of sulfonylurea + metformin had the highest probability (2.1%) to transition to single therapy of metformin (Figure 2.c).

To complement the DFGs, we constructed alluvial plots illustrating drug utilisation variants across consecutive 3-month intervals (presented in the Supplementary Information Results Figures S4-S6). Unlike the DFGs, which apply frequency-based filtering to retain only the most common treatment transitions and simplify pathway visualization, the Sankey plots display all observed patient transitions without path filtering. Consequently, the proportions shown in the Sankey diagrams reflect the full study population at each time interval, whereas the DFG visualisations represent only the most frequent pathways after filtering. This methodological difference explains the variation in percentages between the two visualisation approaches.

**Conformance to NICE:** The DFGs representing treatment pathways, were further compared with the corresponding NICE-recommended treatment pathways. Conformance to guideline-recommended care was quantified using the structural similarity index, with higher values indicating closer alignment. The resulting similarity metrics for risk group are summarised in Table 2.

**Table 2:** The conformance of national level treatment pathways to NICE guidelines, based on *structural similarity index*. Structural similarity is defined in range of [0, 1], with lower similarity indicating differences in medication or combinations appear in the pathway.

|  | LR-C | HR-C | eCVD-C |
| --- | --- | --- | --- |
| **Conformance to NICE NG28, 2022** | 0.22 | 0.21 | 0.39 |

Overall conformance, at national level, was low across all groups, with the LR-C (0.22) and HR-C (0.21) groups showing comparable and similarly poor alignment with guideline-recommended treatment sequences. Conformance was notably higher in the eCVD-C (0.39), though still reflected only modest adherence to the recommended pathway.

### Phase 2: Post-pandemic geographical variation in treatment pathways

At the national level, the process map demonstrates substantially greater variability, reflecting the wide range of different care delivery practices across the nation. This variation results in a higher number of treatment pathway variants, and is associated with lower conformance to guidelines, as a greater proportion of patient journeys deviated from the expected sequence of care. At the ICB level, however, there is more consistency in how care is organised and delivered. More consistency results and substantially reduced variation, hence producing clearer and more interpretable process maps reflecting clinical practice and conformance to NICE guidance. Therefore ICB-level analyses provide a more reliable basis for evaluating treatment pathways and identifying opportunities for quality improvement, while national-level analyses are better suited to detecting broader variation in care delivery across the healthcare system.

**Conformance to NICE:** Optimal DFGs were generated for all 42 ICBs for patients receiving one year of treatment after February 2022 (counts and age ranges are summarised in Supplementary Results Table S5). Figure 3 represents the conformance of each ICB with NICE guidelines, based on their structural similarity.

**Figure 3:**
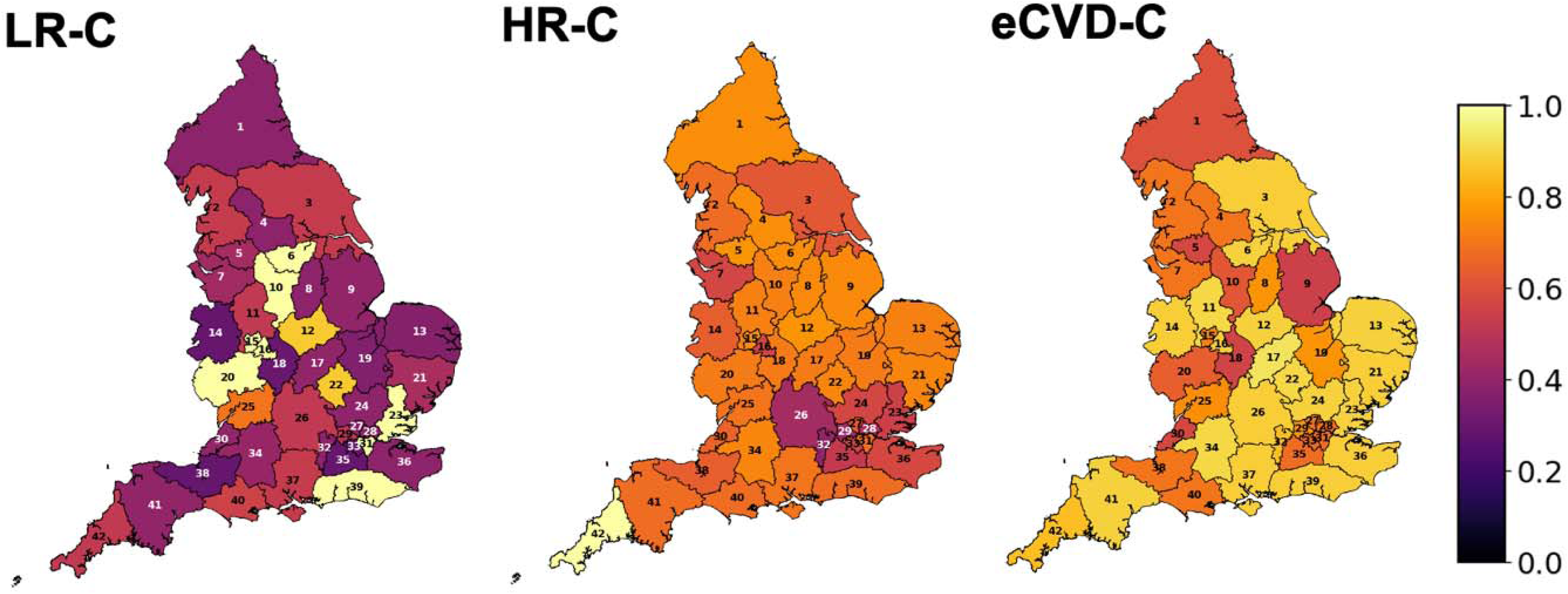
Conformance to NICE guidelines for each cardiovascular risk cohort. The conformance is calculated based on structural similarity criteria and is reported in range of [0, 1], with lower conformance indicating more variability in medication or combinations in treatment pathways compared to NICE guidelines.

Taking each strata in turn:

- For LR-C, conformance to NICE guidelines, varied considerably across the 42 ICBs. The majority of ICBs (n=24, 57.1%) demonstrated low conformance, with structural similarity scores of ≤0.50, indicating substantial divergence from the recommended treatment pathways. A further 8 ICBs (19.0%) showed moderate conformance, with scores between 0.50 and 0.75, suggesting partial alignment with NICE guidance. Only 10 ICBs (23.8%) achieved high conformance, with structural similarity scores exceeding 0.75. Despite the low performance of ICBs in LR-C, 8 out of 10 ICBs showed 100% alignment with NICE guidelines.
- In HR-C moderate conformance to NICE guidelines was observed. The majority of ICBs (n=35, 83.3%) demonstrated structural similarity scores between 0.50 and 0.75, indicating partial but incomplete alignment with the recommended treatment pathways. A small number of ICBs (n=4, 9.5%) showed low conformance, with scores of ≤0.50, and only 3 ICBs (7.1%) achieved high conformance, with structural similarity scores exceeding 0.75, suggesting that close adherence to the recommended pathway was uncommon in this cohort. In this cohort, only 1 ICB (i.e. ICB-42) showed 100% alignment with NICE guidelines.
- In eCVD-C, conformance to NICE guidelines was notably higher compared to other cohorts. No ICBs (0%) demonstrated low conformance, indicating that clear divergence from NICE-recommended pathways was absent in this cohort. The majority of ICBs (n=23, 54.8%) achieved high conformance, with structural similarity scores exceeding 0.75. A further 19 ICBs (45.2%) demonstrated moderate conformance, with scores between 0.50 and 0.75. Although, eCVD-C showed the best conformance to NICE guidelines, no ICB showed 100% alignment.

Altogether, across the three cohorts, a clear gradient in guideline conformance was observed. The LR-C cohort showed a markedly different pattern, with the majority of ICBs (57.1%) falling below the 0.50 threshold, indicating lowest conformance despite this group representing the lowest clinical complexity. Conformance was low in the HR-C cohort, where the distribution was concentrated in the moderate range (83.3% of ICBs scoring between 0.50 and 0.75) with very few ICBs achieving high conformance (7.1%). In contrast, the eCVD cohort demonstrated the strongest adherence to NICE-recommended pathways, with no ICBs in the low conformance band and the majority (54.8%) exceeding the 0.75 threshold. (total percentage of dispensed drugs in each ICB during the post pandemic are presented in Supplementary Results Figure S7).

**Variation in observed treatment pathways:** It is important to note that, although ICBs may demonstrate same level of adherence to clinical guidelines, this does not imply that their treatment pathways are identical. Differences can still exist in the frequency and probability of transitions between treatment states. These differences can be quantified by using one ICB as the reference and comparing the transition frequencies and probabilities of the remaining ICBs against it.

As such, despite the relatively high conformance, for eCVD-C none of the ICBs showed full conformance to NICE guidelines (Figure 3). Among several ICBs which showed the highest similarity score, we selected ICB-17 (0.93), as it shows the highest conformances to NICE guidelines in eCVD-C, as reference ICB. Figure 4.a represents the optimal DFG for the eCVD cohort in this region, illustrating that with a probability of _∼_ 68% the medication pathway starts from metformin, and with _∼_ 32% with SGLT2. In this ICB, patients, who initiated with metformin had a low probability (0.3%) of intensifying to metformin + SGLT2. However, those who initiated the treatment with SGLT2, stayed on SGLT2 until the end of 12 months of study period. Comparison of treatment pathways in ICB-17 with those of other ICBs revealed substantial heterogeneity across regions. Only 2 ICBs (i.e. ICB-22 and ICB-34) demonstrated identical treatment pathways to ICB-17. These are characterised by use of the same therapeutic options (high structural similarity), highly concordant sequencing of treatment escalation and switching (high behavioural similarity), and consistent prescribing patterns (high entropy-based similarity). These similarities indicate that the mapped treatment pathways in these ICBs were fully aligned. In contrast, a subset of ICBs showed marked divergence from ICB-17, with lower overlap in prescribed drug classes and transitions (low-moderate structural similarity), differing patterns of treatment progression (low behavioural similarity), and lower alignment in the distribution and sequence of prescribing patterns relative to ICB-17 (low entropy-based similarity). Variations in observed treatment pathways for LR-C and HR-C are presented in Supplementary Results Figures S8 and S9.

**Figure 4:**
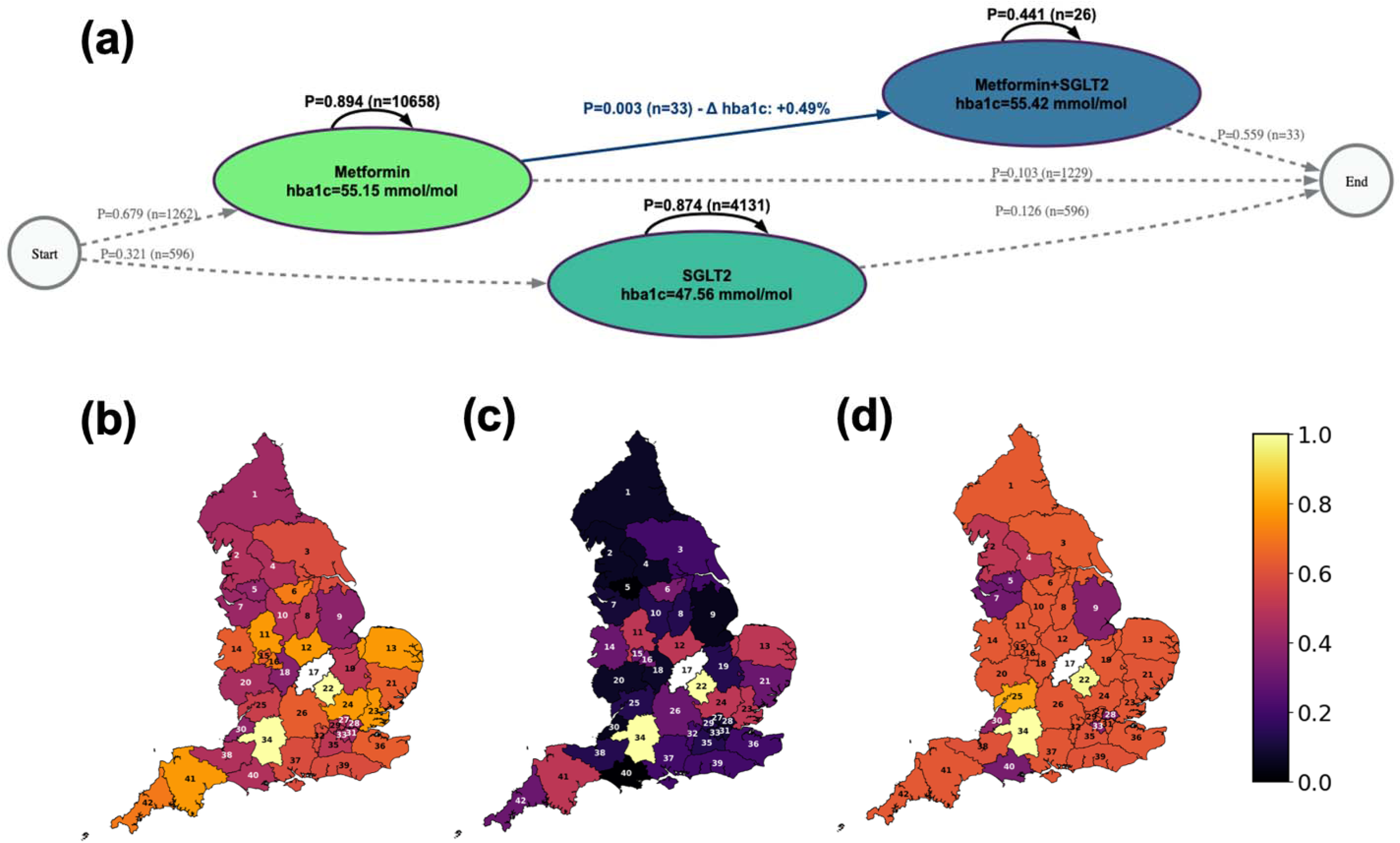
ICB-17 is used as eCVD-C referenced ICB, and the treatment pathways of other ICBs are compared to the reference. (a) Optimal T2DM treatment pathways for patients in eCVD-C for ICB-17 (Fitness = 0.99, Precision = 0.93, F-score = 0.96) (b) Structural, (c) Behavioural, and (d) Entropy-based similarity index between other ICBs and ICB-17 (shown in white), as reference.

## 4. Discussion

This study provides a comprehensive analysis of T2DM treatment pathways through three periods of COVID-19 pandemic across cardiovascular risk cohorts, using process mining of national linked EHR data from England. We compared population-based T2DM treatment pathways observed to the definition of NICE guidelines, published in February 2022, immediately following the COVID-19 pandemic when services were still disrupted. Despite uniform national clinical guidance, we demonstrate substantial non-conformance to the guidelines at national level, following the pandemic and variation in treatment pathways across regions, revealing important gaps between NICE-recommended care and routine prescribing practice. These findings are consistent with longstanding concerns raised by GIRFT regarding potentially unwarranted variation in diabetes care across the NHS. During the post-pandemic recovery phase, increased adoption of newer glucose-lowering therapies, the revised NICE NG28 (2022) guidelines supporting SGLT2 inhibitors[8], and evolving prescribing practices contributed to changes in treatment patterns.

Across all cardiovascular risk cohorts, metformin remained the predominant first-line therapy, with relatively low but gradually increasing uptake of newer medications. Use of SGLT2 inhibitors and DPP4 inhibitors were more common in higher-risk groups, particularly among individuals with established cardiovascular disease, and higher transitions to dual therapy in HR-C were observed, suggesting some adaptation in prescribing practice. SGLT2 inhibitors were preferentially prescribed in the eCVD cohort, reflecting their established benefits for cardiovascular and kidney risk reduction (in line with NICE NG28 guidelines published in 2022 [8]). This pattern indicates clinical uptake among practices towards more risk stratified, evidence-based disease modifying treatments in patients at greatest cardiorenal risk. However, overall uptake of SGLT2 inhibitors remained limited, highlighting ongoing substantial opportunities to accelerate implementation of guideline recommended therapies; perhaps particularly for those at high risk of, but not yet established, cardiovascular disease, who may benefit greatly from increased identification and treatment in line with the UK Governments strategy of shifting treatment to prevention in the 10-Year Health Plan [17].

GLP1 receptor agonists, although prescribed less frequently, were similarly targeted towards individuals with higher cardiovascular risk, and similar trends in the use of these drugs in this group may occur in coming years. Transitions involving less frequently observed treatment switches, such as from metformin to DPP4 inhibitors or SGLT2 inhibitors, were often followed by reductions in HbA1c, although some changes may reflect intolerance or contraindications rather than proactive intensification.

Importantly, national-level patterns do not fully capture the variation in how therapies are implemented at a local level. Regional analysis showed that regional variation was striking. Few ICBs showed close alignment with NICE-recommended sequencing, while others diverged markedly, particularly in the adoption of therapies with established cardiorenal benefit. These differences persisted even after stratification by cardiovascular risk, suggesting that conformance to NICE treatment guidelines vary systematically across patient risk strata, with the most clinically complex group exhibiting the greatest structural alignment with recommended care pathways. This pattern is unlikely to be explained solely by case-mix; rather, it may reflect the more protocolised nature of cardiovascular risk management, greater clinician familiarity with guideline-recommended therapies in higher-risk populations, or differential audit and accountability pressures across ICBs. Taken together, these findings point to meaningful variation in local perceptions of new medicines, prescribing cultures, formulary implementation, and service configuration. This finding echoes GIRFT reviews and aligns with national prescribing data from the NHS Business Services Authority, which have consistently demonstrated wide geographic variation in the uptake of newer diabetes therapies [18]. Together, these observations indicate that national guidance alone is insufficient to ensure consistent delivery of optimal care.

It is important to note that as process maps were generated after frequency-based filtering to retain the most representative treatment pathways, conformance reflects alignment of the dominant process maps with NICE guidance rather than complete guideline adherence at the individual patient level. Moreover, high conformance to NICE guidelines, does not imply that all ICBs follow identical treatment pathways. NICE conformance measures alignment with guideline-recommended sequences and permitted transitions, but it allows for multiple alternative pathways that are still considered compliant. As a result, an ICB can achieve high conformance (i.e., close to 1.0) while implementing a treatment sequence that differs from that observed in another region. Conversely, while our similarity analyses highlight regions with closely aligned treatment pathways, by comparing all ICBs directly to the reference region (ICB-17), high similarity between regions does not necessarily imply adherence to clinical guidelines. Two regions may exhibit highly concordant prescribing patterns relative to each other while still deviating from the recommended NICE sequences. As such, NICE conformance and interregional similarity capture distinct but complementary dimensions of variation. Conformance assesses alignment with recommended pathways, whereas similarity metrics capture real-world consistency in sequencing and intensification across regions. This distinction underscores the value of combining structural, behavioural, and entropy-based similarity measures with guideline conformance to provide a more comprehensive understanding of regional variation in diabetes management beyond guideline compliance alone. Moreover, as conformance to NICE guidance captures only one dimension of ICB performance, an important next step is to build on this framework to better distinguish warranted clinical variation from unwarranted differences, thereby supporting proportionate quality improvement action consistent with GIRFT principles.

Placing these findings in a broader context, our results align with international real-world evidence demonstrating divergence between guideline recommendations and observed treatment sequences in routine care [19]. Analyses using Scandinavian national registries [20], [21], US claims databases [22], and Italian administrative datasets [23], [24] have similarly reported delayed uptake of newer glucose lowering agents and substantial heterogeneity in treatment intensification pathways. In several countries, real-world data analyses have increasingly been incorporated into national diabetes strategy updates, recognising that methods such as process mining can reveal prescribing behaviours, system bottlenecks, and unwarranted variation that are not captured through traditional audit or performance metrics [25]. Our study adds to this international evidence base by demonstrating, at national scale, how linked routine data can be used not only to quantify variation but to visualise treatment pathways and identify where guideline-concordant care breaks down in practice. The persistence of marked regional variation has important policy implications in the context of current NHS practice in England. The proposed single national formulary (SNF), outlined in the NHS 10-year plan [17], offers a potential mechanism to reduce unwarranted variation, improve equitable access to evidence-based therapies, and support more consistent implementation of NICE recommendations. However, our findings suggest that formulary alignment alone will be insufficient without complementary data-driven approaches to monitor local implementation strategies. Embedding affordable real-world pathway analysis within a learning health system framework would enable continuous feedback on prescribing behaviour, support benchmarking across regions, and provide actionable intelligence for clinicians, commissioners, and policymakers.

**Limitations:** In this study, we considered medication dispensing data as a proxy for prescribed medications. However, the process mining methodology is sensitive to the precise sequencing of treatment events and during the COVID-19 pandemic period, there were clear disruptions to healthcare delivery, including delays between prescription and dispensing. Therefore, this may have introduced artificial sequencing anomalies that do not reflect true clinical decision-making. We attempted to minimise this bias by introducing an imputation process that accounted for the prescribed dosage ( discussed in Supplementary Information Methods 1.4); however, the possibility of residual bias from this source cannot be entirely excluded. As such, observed pathway differences during this period should therefore be interpreted with appropriate caution.

Moreover, the selection of incident treatment populations across the three comparator periods carries a risk of introducing collider bias, as the assumption of comparability between patients initiating therapy may not hold for the pandemic period, during which reduced access to primary and secondary care services likely altered the clinical profile of those entering treatment, meaning that pathway differences observed between periods may partly reflect changes in the patient population being treated rather than genuine shifts in prescribing practice.

Further, whilst process mining captures the structure and sequencing of prescribed treatments at a population level, it does not incorporate patient-level clinical nuance; specifically, clinically important drivers of treatment modification. As such, dose escalation, patient intolerance, hypoglycaemia, frailty, and individual patient preference, are not captured within the event log data, and their absence means that some observed pathway transitions may reflect complex clinical reasoning that cannot be fully distinguished from variation in prescribing behaviour within this framework. It is also important to note that there is a theoretical possibility that infrequent but guideline conformant pathways could be removed by frequency filtering and bias conformance scores, this is unlikely to be true as guideline-recommended treatment sequences, by virtue of being standard of care, are expected to constitute the most frequently observed pathways in the data. Rare or filtered pathways are more likely to represent outliers or data artefacts rather than guideline conformant behaviour.

## 5. Conclusion

In conclusion, this study demonstrates a novel methodology for measuring the adherence of NICE guidance, using routinely collected, linked health data through three periods of COVID-19. Using process mining of national linked EHR data from England, this study demonstrates that T2DM treatment pathways across cardiovascular risk cohorts diverged substantially from NICE guidance after the COVID-19 pandemic. Despite the existence of uniform national clinical guidance, published in February 2022 while services remained disrupted, we found considerable non-conformance at the national level and marked regional variation in prescribing practice, highlighting persistent gaps between recommended and routine care.

This methodology can be used not only for describing variation in care, but ultimately for enabling a learning health system capable of supporting continuous improvement in T2DM management. Process mining of population-level prescribing data provides a scalable approach to track uptake of innovative treatments, assess concordance with NICE and GIRFT recommendations, and identify where and for whom guideline-concordant care is not being realised. Crucially, the same linked data infrastructure can support more personalised care by identifying which patient groups are most likely to benefit from newer therapies, such as SGLT2 inhibitors and GLP1 receptor agonists, and how these treatments are being sequenced in routine practice. Embedding such analytic capability within NHS and NICE decision making would allow national priorities to be translated into local action, enabling innovation in care to be implemented safely, equitably, and at pace. T2DM is used as an example here, but these methods could be applied across care pathways incorporating non-pharmacological interventions and processes to improve care and its outcomes not only in the UK, but in other health economies with similarly linked data.

## Supplementary information

Supplementary Information is included.

## Data Availability

The data used in this study are available in NHS England Secure Data Environment (SDE) service for England, but as restrictions apply they are not publicly available.

## Acknowledgement

The authors thank Prof Spiros Denaxas and Prof Evangelos Kontopantelis for their insightful comments on this manuscript.

This work was carried out with the support of the BHF Data Science Centre led by Health Data Research UK (BHF Grant no. SP/19/3/34678). This study made use of anonymised data held in NHS England’s Secure Data Environment service for England and made available via the BHF Data Science Centre’s CVD-COVID-UK/COVID-IMPACT consortium. This work used data provided by patients and collected by the NHS as part of their care and support. We would also like to acknowledge all data providers who make health relevant data available for research.

## Declarations

PB and RS are funded by the National Institute for Health and Care Research (NIHR), RS’ research professorship, NIHR303160, for this research project. KK is supported by the National Institute for Health Research (NIHR) Applied Research Collaboration East Midlands (ARC EM), NIHR Global Research Centre for Multiple Long-Term Conditions, NIHR Cross NIHR Collaboration for Multiple Long-Term Conditions, NIHR Leicester Biomedical Research Centre (BRC) and the British Heart Foundation (BHF) Centre of Excellence. The views expressed are those of the author(s) and not necessarily those of the NIHR, NHS or the Department of Health and Social Care.

## Ethical approval

The North East - Newcastle and North Tyneside 2 research ethics committee provided ethical approval for the CVD-COVID-UK/COVID-IMPACT research programme (REC No 20/NE/0161) to access, within secure trusted research environments, unconsented, whole-population, anonymised data from electronic health records collected as part of patients’ routine healthcare.

## Data availability

The data used in this study are available in NHS England’s Secure Data Environment (SDE) service for England, but as restrictions apply they are not publicly available (https://digital.nhs.uk/services/secure-data-environment-service). The CVD-COVID-UK/COVID-IMPACT research programme, led by the BHF Data Science Centre (https://bhfdatasciencecentre.org/), received approval to access data in NHS England’s SDE service for England from the Advisory Group for Data (AGD) (https://digital.nhs.uk/about-nhs-digital/corporate-information-and-documents/advisory-group-for-data) – formerly the Independent Group Advising on the Release of Data (IGARD) – via an application made in the Data Access Request Service (DARS) Online system (ref. DARS-NIC-381078-Y9C5K) (https://digital.nhs.uk/services/data-access-request-service-dars/dars-products-and-services). The CVD-COVID-UK/COVID-IMPACT Approvals & Oversight Board (https://bhfdatasciencecentre.org/areas/cvd-covid-uk-covid-impact/) subsequently granted approval to this project (CCU014) to access the data within NHS England’s SDE service for England. The anonymised data used in this study were made available to accredited researchers only. Those wishing to gain access to the data should follow the application process of the relevant national data custodian.

