## Supplementary information for "Measuring implementation of clinical guidelines through the COVID-19 pandemic, using linked national health records: a national study of type 2 diabetes in England"

**Conformance to NICE guidelines in routine clinical practice: evidence from population data**

### 1 Methods

#### 1.1 Diabetes medication categorisation

Diabetes medications were identified using the British National Formulary (BNF) Chapter 6 Section 1 (BNF 6.1: Drugs used in diabetes). The drugs were further mapped to drugs used for T2DM treatment according to their chemical substance, as presented in Table S1.

#### 1.2 Chronic heart failure and established atherosclerotic CVD

The SNOMED and ICD10 codes for heart failure were taken from Phenotype library [1], presented in Code list Table S7. Furthermore, corresponding to NICE guidelines’ definition of established atherosclerotic CVD, the code lists for coronary heart disease, acute coronary syndrome, previous myocardial infarction [2], stable angina, prior coronary or other revascularisation, peripheral arterial disease [3], and cerebrovascular disease [4] such as ischaemic stroke and transient ischaemic attack [5]. The code list corresponding to chronic heart failure and established CVD are reported in Code list section Tables S7 and S8, respectively.

#### 1.3 QRISK3 calculation

QRISK3 score is calculated based on the algorithm developed by Hippisley-Cox et al. [6]. Accordingly, a separate dataset is curated to contain all the information required for QRISK3 calculation. This curated dataset will contain one row of data per individual, with each column representing one of the information required for the calculation. As such, the following information is extracted from the individual’s historical data:

- **Age:** defined at time *t*_0_.
- **Sex:** defined as Female or Male. The QRISK3 algorithm and its associated coefficients differ for each sex.
- **Ethnicity:** is considered as 9 categories in line with QRISK3 algorithm. Accordingly, the 18-ethnicity categorisation of multi-source demographic curated dataset,

**Table S1**: Diabetes drug categorisation according to their chemical substances

| **Drug Category** | **Chemical Substance** | **BNF code count** |
| --- | --- | --- |
| Metformin | Metformin hydrochloride | 47 |
| Insulin | Insulin | 232 |
| Glucosidase | Acarbose | 4 |
| DPP4 | Sitagliptin, Linagliptin,  Saxagliptine, Alogliptin,  Vildagliptine | 22 |
| PPARY | Pioglitazone, hydrochloride,  Rosiglitazone | 25 |
| SGLT2 | Dapagliflozin, Canagliflozin,  Empagliflozin, Ertugliflozin | 16 |
| Sulfonylurea | Glimepiride, Chlorpropamide,  Clibenclamide, Gliclazide, Glipizide,Gliquidone, Tolbutamide | 84 |
| Meglitinide | Repaglinide, Nateglinide | 17 |
| GLP1 | Liraglutide, Lixisenatide,  Dulaglutide, Albiglutide,  Semaglutide, Exenatide | 43 |
| Metformin/DPP4 | Sitagliptin+Metformin,  Linagliptin+Metformin,  Saxagliptine+Metformin, Alogliptin+Metformin,  Vildagliptine+Metformin | 16 |
| Metformin/PPARY | Rosiglitazone+Metformin,  Pioglitazone+Metformin | 10 |
| Metformin/SGLT2 | Dapagliflozin+Metformin,  Canagliflozin+Metformin,  Empagliflozin+Metformin | 16 |
| Combination | Empagliflozin+Linagliptin,  Insulin+Lixisenatide,  Insulin+Liraglutide,  Saxagliptin+Dapagliflozin | 12 |

developed quarterly by the Consortium’s health data science team, is mapped to include these 9 categories, as per Table S2.

- **Deprivation Index:** QRISK3 algorithm relates the post code to the Townsend score, which is a measure of material deprivation. However, in the SDE the Lower Layer Super Output Area (LSOA) information is mapped to the Index of Multiple Deprivation (IMD) score, which is a measure of multi-dimensional deprivation, and describes deprivation in more details. It has been indicated that both indices represent similar broad patterns of deprivation [7]. Therefore, given the availability of data in this study, we assumed a linear relationship between the two scores in each quintile, considering that both scores show higher deprivation with higher values. Accordingly, the IMD scores corresponding to individual’s LSOA are linearly

**Table S2**: Multi-source Ethnicity mapping to QRISK3 algorithm

| **QRISK3 ethnicity** | **Curated multi-source ethnicity (Code value)** |
| --- | --- |
| White or not stated | British (A)  Irish (B)  Any other white background (C)  Traveler (T)  Not stated (Null) |
| Indian | Indian (H) |
| Pakistani | Pakistani (J) |
| Bangladeshi | Bangladeshi (K) |
| Other Asian | Any other Asian background (L) |
| Black Caribbean | Caribbean (M) |
| Black African | African (N)  Any other black background (P) |
| Chinese | Chinese (R) |
| Other Ethnic Groups | White & black African (E)  White & black Caribbean (D)  Any other mixed background (G)  White & Asian (F)  Arab (W)  Any other ethnic group (S) |

projected to Townsend scores as per Eq. 1, to be used for our QRISK3 calculation.

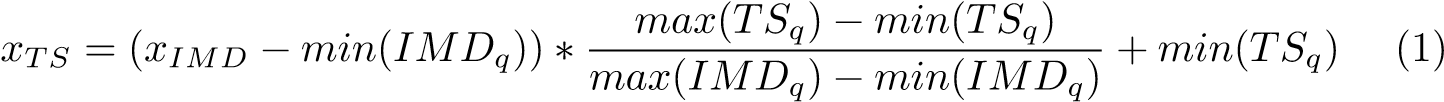

where *IMD_q_* and *TS_q_* represent the IMD and Townsend scores of each quintile, respectively, as presented in Table S3. *x_IMD_* and *x_TS_* are defined as the corresponding IMD and Townsend score of each LSOA. The values of *x_IMD_* for each LSOA is available in the SDE.

**Table S3**: Quintile Townsend and IMD band cutoffs

| **Quintile** | **Townsend band cutoffs [8]** | **IMD band cutoffs [9]** |
| --- | --- | --- |
| 1 | [-6.61, -3.01] | [0.4770 , 8.3720] |
| 2 | [-3.00 , -1.32] | [8.3730 , 13.9220] |
| 3 | [-1.31 , 0.95] | [13.9240, 21.4320] |
| 4 | [0.96 , 3.73] | [21.4340 , 33.8760] |
| 5 | [3.74 , 11.06] | [33.8810 , 92.6010] |

- **Smoking status:** QRISK3 considers 5 smoking classes as non-smoker, ex-smoker, light (*<* 10 cigarettes per day), moderate (10−19 cigarettes per day), and heavy smoker (*>*= 20 cigarettes per day). The associated SNOMED codes for each of the 5 categories are listed in Codelist Table S9. From the table, the severity of smoking can be implied in some cases when there is no definite mention of the cigs per day. However, there are few SNOMED codes which define a smoker status with unknown severity. To address the individuals who have these SNOMED codes in their historical data, we assumed the unknown severity status to be light smokers. Further, the individuals with no associated smoking code in their GDPPR historical data are considered as non-smokers.
- **Diabetes status:** Given the design of our study, all the individuals in our study population have T2DM.
- **Body Mass Index (BMI):** The associated SNOMED codes for BMI are presented in Codelist Table S10. Notably, QRISK3 algorithm requires continuous numerical values for BMI. Therefore, BMI values are taken from GDPPR dataset for the latest date of individual’s history. Further, in case where an individual has a BMI SNOMED code associated to them in their historical data, but with no value, the BMI is imputed using an average value in the range defined by each code. Also, any individual with no associated BMI code is considered to have a normal *BMI* = 22.
- **Angina or heart attack in a** 1*^st^* **degree relative:** SNOMED codes associated with Angina and heart attack in family members are collected from SNOMED CT library [10], as well as codes available in BHF caliber for stable Angina and Myocardial Infraction corresponding to family history. The Codelist was further searched in GDPPR dataset to check their availability at national level records. Accordingly, the list is detailed in Codelist Table S11. These criteria will make up a binary class where 1 is defined for individuals who have a code linked to family history of Angina or heart attack in their historical data.
- **Chronic kidney disease (CKD) stages 3, 4 or 5:** The associated Codelist for CKD is obtained from HDRUK Phenotype Library [11] and is listed in Table S12. Consequently, the CKD codes will form a binary classification where a value of 1 indicates that the individuals have had CKD in their history, prior to the analysis.
- **Atrial Fibrillation (AF):** The Codelist for AF is collected from HDRUK Phenotype Library [12] and is listed in Table S13. These criteria will also make a binary classification to denote the individuals with AF in their historical data.
- **Migraines:** The SNOMED and ICD10 codes for Migraine are obtained from CALIBER phenotyping library, as listed in S14. Migraine will also make up a binary classification in our curated dataset for QRISK3 calculation to denote the individuals with migraine in their historical data.
- **Systemic lupus erythematosus (SLE):** The SNOMED and ICD10 codes for SLE are obtained from CALIBER phenotyping library, as listed in S15. SLE will make a binary classification in our curated dataset to denote the individuals with SLE codes associated with them in their historical data.
- **Rheumatoid Arthritis:** SNOMED codes associated with RA are obtained by searching the GDPPR reference set for “*Rheumatoid Arthritis*”. The list of codes found in the first step were further searched in GDPPR to eliminate the ones that have not been used in the whole dataset. The final Codelist is presented in Table S16. These criterial will make a binary classification based on the historical data to enhance the QRISK3 calculation
- **Steroid tablets:** Individual’s historical data of NHSBSA are considered for the medications in BNF Chapter 6, section 3, paragraph 2 (BNF060302) “Glucocorticoid therapy” for Corticosteroid use [6]. This information is also presented as a binary data.
- **Erectile dysfunction:** The individual’s NHSBSA historical records are reviewed for medications listed under BNF Chapter 7, Section 4, Paragraph 5 (BNF070405), titled “Drugs for erectile dysfunction”, to identify as Erectile dysfunction medication use [6]. This data is represented in binary form.
- **Atypical antipsychotic medication:** This group of medication is identified from an individual’s NHSBSA historical data by examining medications categorised under BNF Chapter 4, Section 2, Paragraph 1 (BNF040201) “Antipsychotic drugs”. This information is, also, recorded as binary data.
- **Blood pressure treatment:** This information also represented as binary data is extracted from individual’s NHSBSA historical data corresponding to BNF Chapter 2, Section 5, Paragraph 1 (BNF020501) “Vasodilator antihypertensive drugs”.
- **Severe Mental Illness:** Phenotype library was used to collect the SNOMED and ICD10 codes for Schizophrenia [13], and bipolar affective disease [14]. The full list of mental illnesses considered in the study is presented in Codelist section S17. Individual’s historical GDDPR and HES data are considered for extracting information regarding mental illnesses. The data is presented in binary.

#### 1.4 Medication Data Processing and Imputation

Following NICE guidelines [5], T2DM treatment is optimised according to individual clinical circumstances, with switching or adding treatments from different drug classes (up to triple therapy) as appropriate. For process-mining analyses, it is crucial that event logs are as clean and coherent as possible to accurately capture treatment trajectories. However, EHR data can contain discontinuities, particularly in NHSBSA prescription records, where medications may be dispensed in bulk for multiple months. To address this, we applied an imputation method using a 3-month moving average of prescribed quantities. Briefly, if the moving average was zero, indicating no prescriptions in the preceding two months, a “loss cost” equivalent to one month’s supply (–28) was applied to account for potential missing prescriptions. If the prescribed quantity was below the moving average, no adjustment was made, as the medication was likely still being consumed. Conversely, if the prescribed quantity exceeded the moving average, a –28 adjustment was applied to correct for bulk prescribing. This step was repeated on the adjusted values, and months with positive values were considered as having a prescribed drug. This approach allows for a more consistent, continuous time series of medication use and is detailed further in the Supplementary Information. These assumptions were necessary due to the lack of established methods for imputing non-recorded prescription data in EHRs.

#### 1.5 HbA1c data processing and analysis

HbA1c measurements were extracted for individuals in the post-Covid19 cohort, considering values recorded within the 6 months preceding treatment initiation *t*0. At least one HbA1c measurement in this window was available for 683,806 individuals out of 708,935. Because HbA1c testing is performed intermittently in routine clinical practice and does not align with the monthly resolution of prescription data, a last observation carried forward (LOCF) approach was applied to impute HbA1c values for months without recorded measurements. This approach assumes stability of HbA1c between consecutive clinical assessments and is commonly used in longitudinal observational analyses.

For each treatment transition or intensification state identified through process mining, changes in glycaemic control were quantified by calculating the percentage difference between the HbA1c value immediately preceding the transition (*HbA*1*c_prev_*) and the subsequent available HbA1c measurement (*HbA*1*c_next_*), defined as:

*
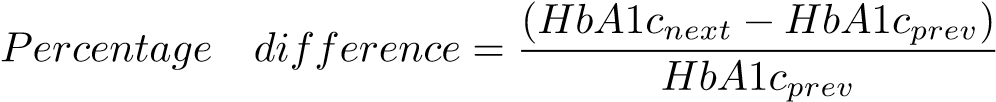
100	(2)

For each transition, the median percentage change in HbA1c was calculated across the population, alongside the mean and standard deviation to characterise variability. These estimates were integrated into process maps, where directional changes in HbA1c were visually encoded: red arrows indicated median decreases in HbA1c, while blue arrows indicated increases. Arrow labels displayed both the frequency of the transition and the corresponding mean ± standard deviation of HbA1c change, enabling simultaneous interpretation of treatment pathways and associated glycaemic outcomes.

#### 1.6 Process mining considerations

Process mining is the intersection of data science and business process management that involves computational approaches to extract structured information and process related knowledge from event logs [15]. It integrates principles from data mining and process modelling to reconstruct, analyse, and visualise real-world process flows. Given that EHR systems inherently generate time-stamped event data, they provide a suitable foundation for process mining applications in clinical research. In this study, we applied process mining techniques to EHR data to identify and visualise the longitudinal trajectories of antidiabetic drug use in the 12 months following the initiation of treatment of T2DM. The analysis leveraged the inherent event log structure of the EHR, wherein each drug prescription was treated as a discrete event associated with a timestamp and patient identifier.

To enhance interpretability and reduce visual complexity, the DFGs were optimised through frequency-based filtering. This step was essential to remove infrequent transitions and to emphasise the most representative pathways of drug utilisation. Without such filtering, the resulting process maps often suffer from excessive complexity, commonly referred to as “spaghetti models”, in which the density of nodes and edges obscures meaningful insights and hinders the analysis [16]. The optimisation was performed iteratively using a trial-and-error approach, where multiple filtering thresholds for activity and path frequencies were applied. The resulting process maps were evaluated based on their F scores to assess model quality. The model with the highest F score was selected as the optimal representation, where *F-score*, as the harmonic mean of Precision and Fitness, was calculated as:

$F-score=2* \frac{Fitness*Precision}{Fitness+Precision}$	(3)

where, Fitness quantifies the extent to which the discovered process model can reproduce the observed behaviour in the event log. Using token-based replay, fitness is defined as:

$Fitness= \frac{1}{2}(1-\frac{m}{c}+1-\frac{r}{p})$	(4)

where c and p denote the number of consumed and produced tokens, respectively, m represents the number of missing tokens required to replay the observed traces, and r denotes the number of remaining tokens after replay. Fitness values range from 0 to 1, with higher values indicating better agreement between the model and the observed event log.

Precision measures the extent to which the model restricts behaviour to that observed in the event log by penalising additional behaviour allowed by the model but not supported by the data. Precision was computed using an alignment-based approach and is defined as:

$Precision=1- \frac{\sum_{s\in S} w\left( s \right) esc(s)}{\sum_{s\in S} w\left( s \right) out(s)}$	(5)

where S denotes the set of reachable states during log replay, out(s) is the number of enabled transitions from state s, esc(s) is the number of transitions enabled by the model but not observed in the event log, and w(s) represents the frequency-based weight of state s. Precision values range from 0 to 1, with higher values indicating less allowance for unobserved behaviour.

#### 1.7 Similarity measures

Given the extent of the analysis in both phases, we calculated three similarity measures to quantify the variability and alignment between treatment pathways. This would enable easier comparison between time periods, regional, and socioeconomic status in phase 1 and 2 of the study, respectively. The three measures are defined as follows:

- **Structural similarity** assesses the degree of overlap in the topology of process pathways, considering the nodes (i.e. medication) and their connections (i.e. transitions). It is calculated using a Jaccard-like index [17]:

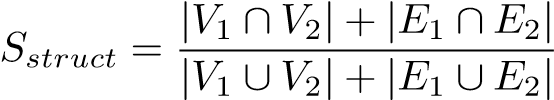
(4)

where *V_i_* and *E_i_* denote the set of nodes and edges in process path *i*, and |*.*| represents the cardinality of each set.

- **Behavioural similarity** evaluates the extent to which the pathways produce comparable outcomes, focusing on execution or functional equivalence. This measure focuses on behavioural profiles by capturing the characteristics of process models by means of relations between each activity (node) pair. It is defined as the proportion of shared computational paths or outcome similarity [18]:

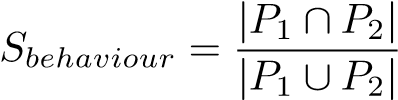
(5)

where *P_i_* denotes the set of execution paths or observable outputs for each process path *i*.

- **Entropy-based similarity** captures the degree of similarity in the information content and variability of the pathways, reflecting the distribution and diversity of actions or events. It can be quantified using the Jensen–Shannon divergence of the entropy distributions:

*Sentropy* = 1 − *JSD*(*H*1*,H*2)	(6)

where *H_i_* represents the entropy distribution of actions or interventions in process pathway *i*. *JSD* is a symmetric and finite divergence calculation, widely used in information theory [19]. This measure captures how similar the information content or complexity patterns are between the two process pathways, rather than just structure or outputs.

The similarity measures are defined in range of [0,1], with higher values representing greater similarity between the two process pathways under study. Table S4 represents the clinical interpretation of the combined similarity measures of T2DM treatment pathway.

Conformance of observed treatment pathways to NICE guideline recommendations was quantified using a structural similarity approach. Each real-world pathway was compared to the guideline-defined process, considering three components: edges (the allowed transitions between therapies), start activities (therapies permitted at pathway initiation), and end activities (therapies permitted at pathway termination). Similarity for each component was calculated as the proportion of observed elements that matched the guideline. An overall guideline conformance score was then computed as a weighted sum of the three components: edge similarity weighted 0.7, start activity similarity 0.15, and end activity similarity 0.15. This score ranges from 0 to 1, with higher values indicating greater adherence of observed pathways to guideline recommendations.

**Table S4**: Clinical interpretation of combined similarity measures of treatment pathways. Combinations of high and low values across these measures reflect differences in medication selection, sequencing, and variability. This framework allows clinicians to interpret whether pathways are standardised, predictable, or heterogeneous, and to identify areas of consistent versus variable treatment practice across patient subgroups.

| **Structural** | **Behavioural** | **Entropy-based** | **Clinical Interpretation** |
| --- | --- | --- | --- |
| High | High | High | Pathways are very similar in drugs, sequence, and predictability. Clinically, this indicates highly standardised care: most patients receive the same medications in the same order with minimal variation. |
| High | Low | High | Same medications used, but sequencing differs. Clinically, the choice of drugs is consistent, but timing of intensification or switching varies across clinicians or subgroups. |
| High | High | Low | Same medications and sequence, but greater dispersion in prescribing patterns. Clinically, suggests that although the same pathways are followed, there is some variability in frequency or adherence to sequences, possibly reflecting individualised treatment adjustments. |
| High | Low | Low | Same medications but differing sequence and variable patterns. Clinically, indicates predictable drug choices, but treatment progression and application are heterogeneous; patient level factors or clinician discretion may drive differences. |
| Low | High | High | Different medications, but similar sequence and predictable patterns. Clinically, suggests alternative standardised treatment strategies are being applied across groups, perhaps due to formulary differences or regional guidelines. |
| Low | Low | High | Different medications and sequences, but patterns are consistent within each group. Clinically, pathways diverge in drugs and order, yet each pathway is internally predictable, reflecting distinct but structured care approaches. |
| Low | High | Low | Different drugs with similar sequencing, but high variability. Clinically, treatments are heterogeneous in selection, and sequencing is not consistent in frequency, indicating both diverse clinical strategies and individualised adjustments. |
| Low | Low | Low | Highly variable pathways in drug choice, sequencing, and predictability. Clinically, reflects the greatest heterogeneity: patients experience very different therapies and treatment progressions, likely due to individualised care, complex comorbidities, or inconsistent guideline implementation. |

#### 1.8 Calculation of conformance to NICE guidelines

To quantify conformance with NICE guideline pathways, we computed a structural similarity index between the guideline-derived process model and observed real-world care pathways. Guideline pathways were represented as a directly flow graph (DFG) specifying allowed transitions (edges) between clinical activities, along with defined start and end activities. Real-world pathways were similarly encoded as a set of observed transitions and, where available, start and end activities, derived from process mining. Structural similarity was assessed across three components: **(i) edge similarity**, defined as the proportion of observed transitions that were present in the set of guideline-allowed transitions; (ii) **start activity similarity**, calculated as the proportion of observed starting activities that matched guideline-defined starting activities; and (iii) **end activity similarity**, defined as the proportion of observed ending activities that corresponded to any permissible terminal activity in the guideline model. An overall conformance score was computed as a weighted sum of these components, with weights of 0.7 for edge similarity, 0.15 for start similarity, and 0.15 for end similarity, reflecting the primary importance of transition structure in pathway adherence. All similarity measures were normalized to the range [0,1], where higher values indicate greater conformance to guideline-defined care processes.

### 2 Results

The demographic characteristics of the individuals who met inclusion criteria are summarised in Table S5. Optimal process maps for LR-C, eCVD-C, and HR-C, for pre and during-COVID-19 demonstrate high performance scores for all risk groups, as presented in Figure S1, and S2 respectively, indicating strong alignment between mapped and observed behaviours.

**Table S5:** The count and age range of patients with T2DM for the three cardiovascular risk groups, according to NICE NG28 2022 guidelines, in England.

|  |  | **LR-C** | **HR-C** | **eCVD-C** |
| --- | --- | --- | --- | --- |
| **Pre COVID-19** | Count (% female)  Mean Age (±std) | 10,505 (50.31%)  42.8 (±8.21) | 45,520 (42.11%)  59.25 (±11.22) | 12,950 (32,36%)  66.11 (±10.50) |
| **During COVID-19** | Count (% female)  Mean Age (±std) | 16,720 (50.72%)  42.35 (±8.38) | 72,325 (43.88%)  58.77 (±11.20) | 21,470 (34.47%)  65.55 (±10.51) |

(a) LR-C, Precision = 0.94, Fitness = 0.99, F-score = 0.96

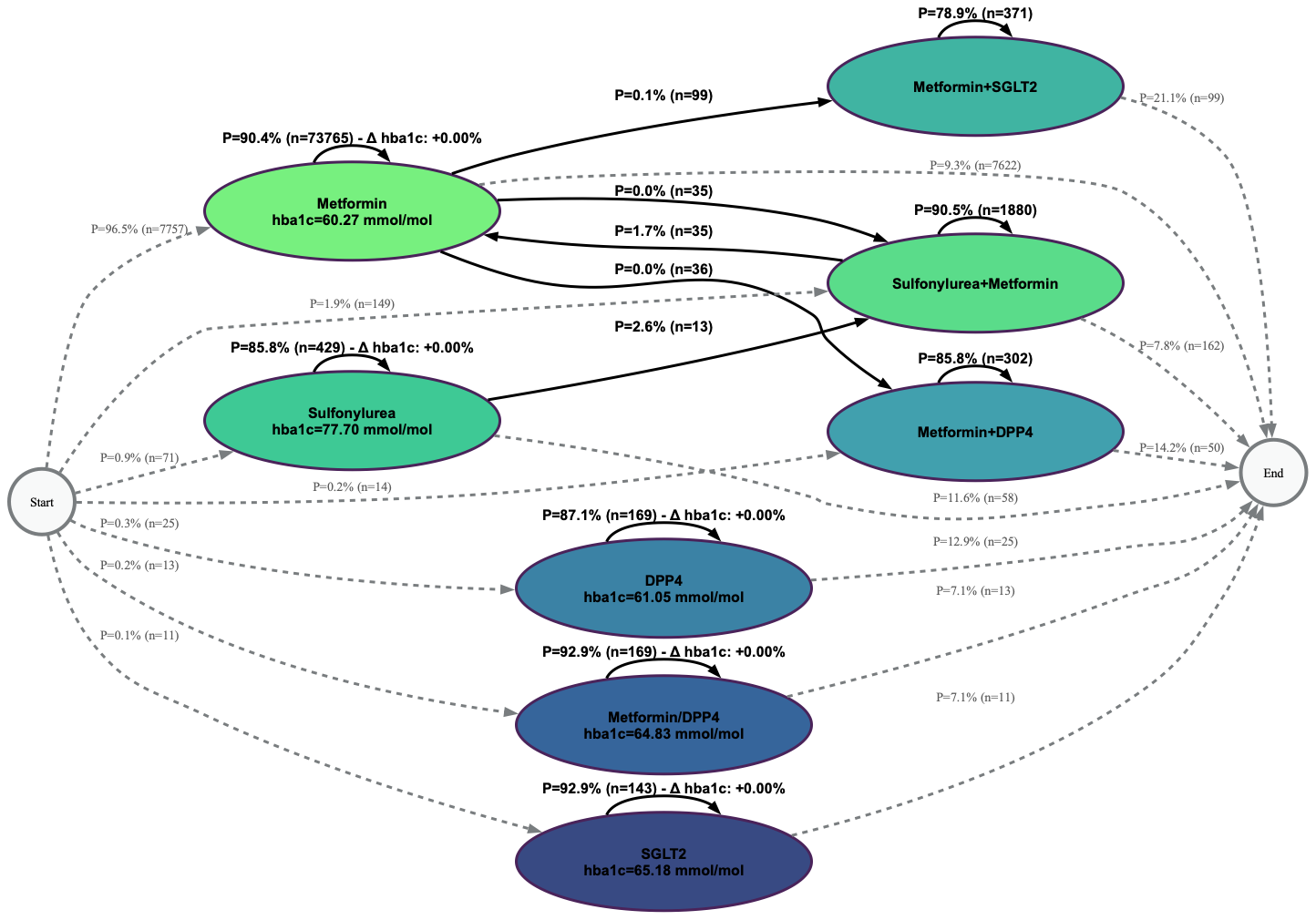

(b) HR-C, Precision = 0.96, Fitness = 0.99, F-score = 0.98

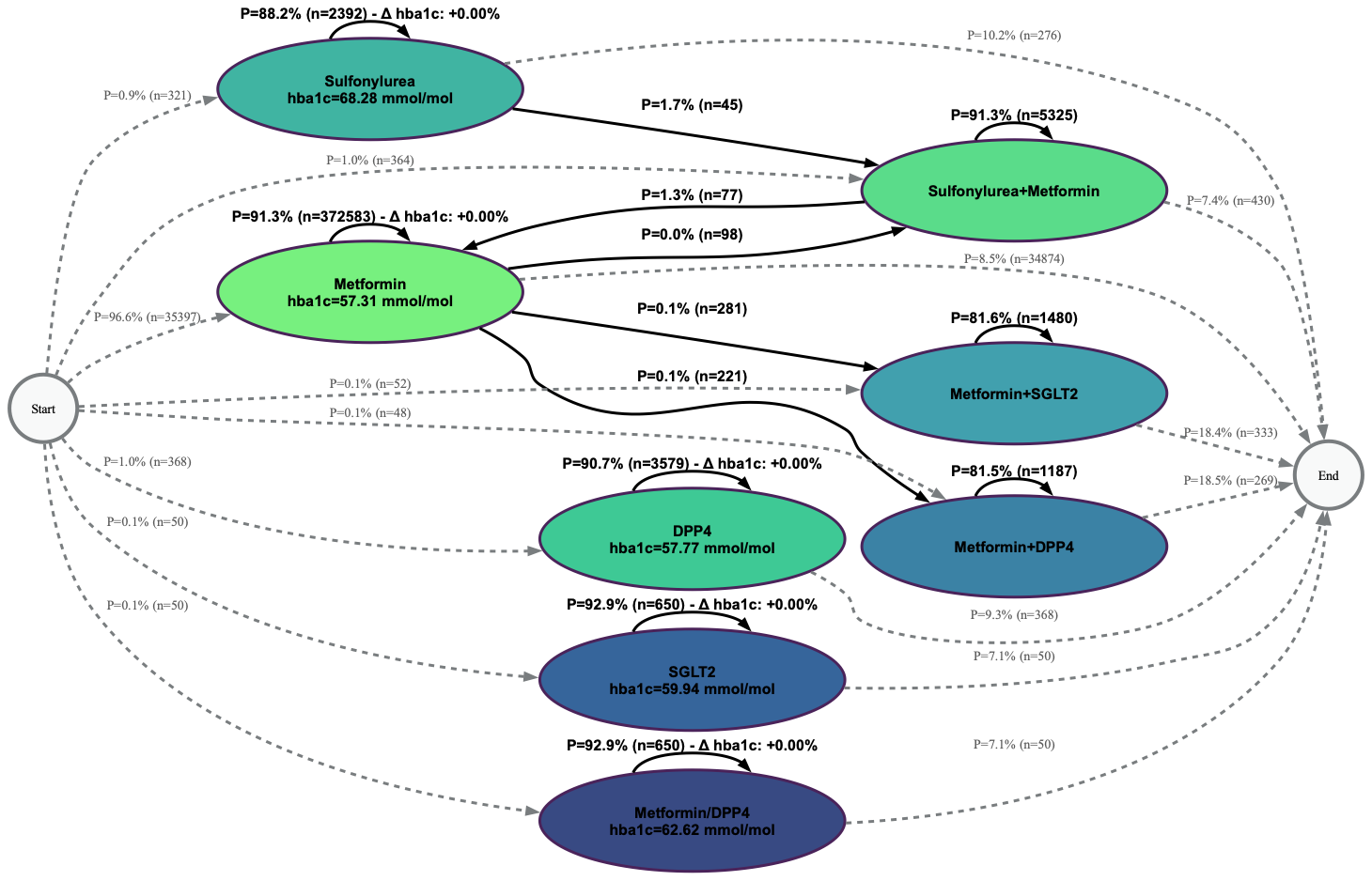

(c ) eCVD, Precision = 0.96, Fitness = 0.99, F-score = 0.98

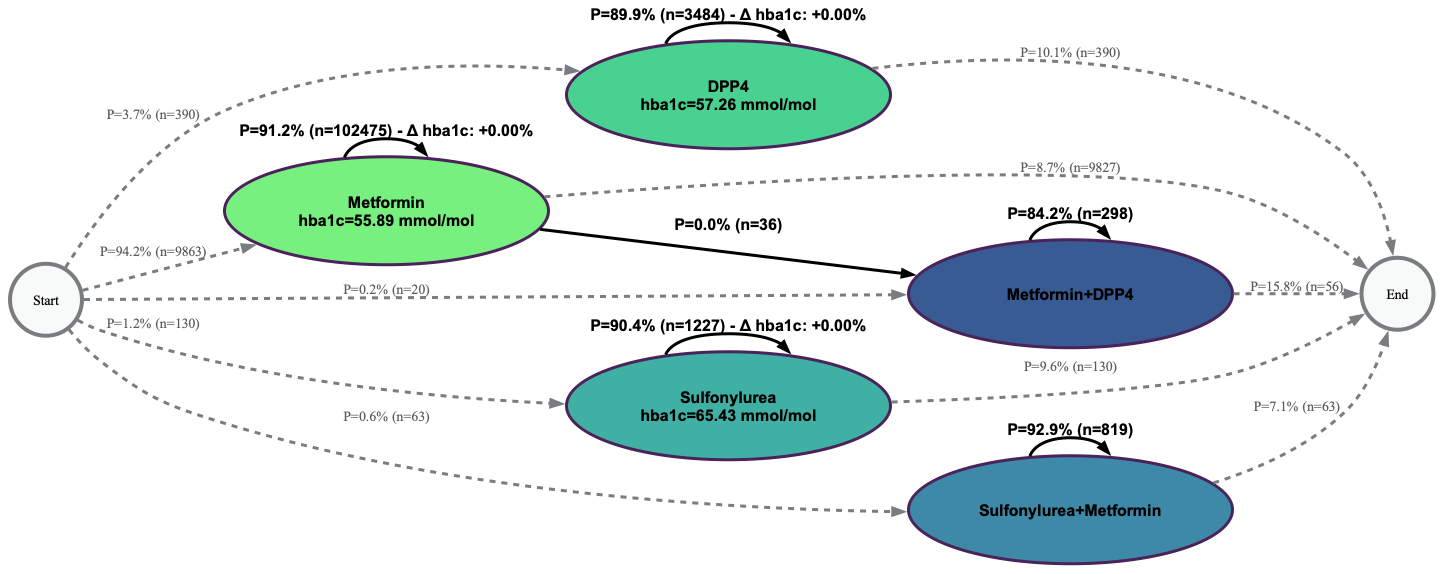

**Figure S1:** First-line treatment and further interventions for patients with T2DM in **pre COVID-19**, shown as optimal process maps in the format of Directly-Follows Graph (DFG), in (a) low risk of CVD cohort (LR-C), (b) high risk of CVD cohort (HR-C), and (c) patients with established CVD (eCVD-C).

(a) LR-C, Precision = 0.94, Fitness = 0.96, F-score = 0.98

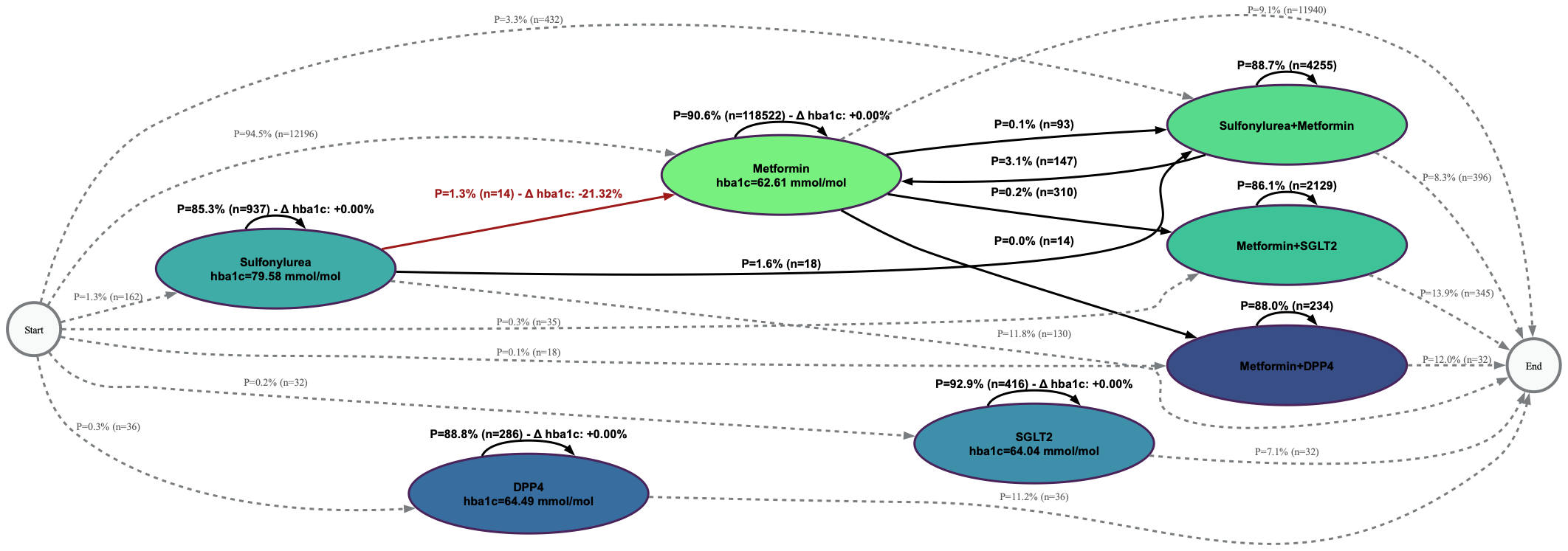

(b) HR-C, Precision = 0.96, Fitness = 0.99, F-score = 0.98

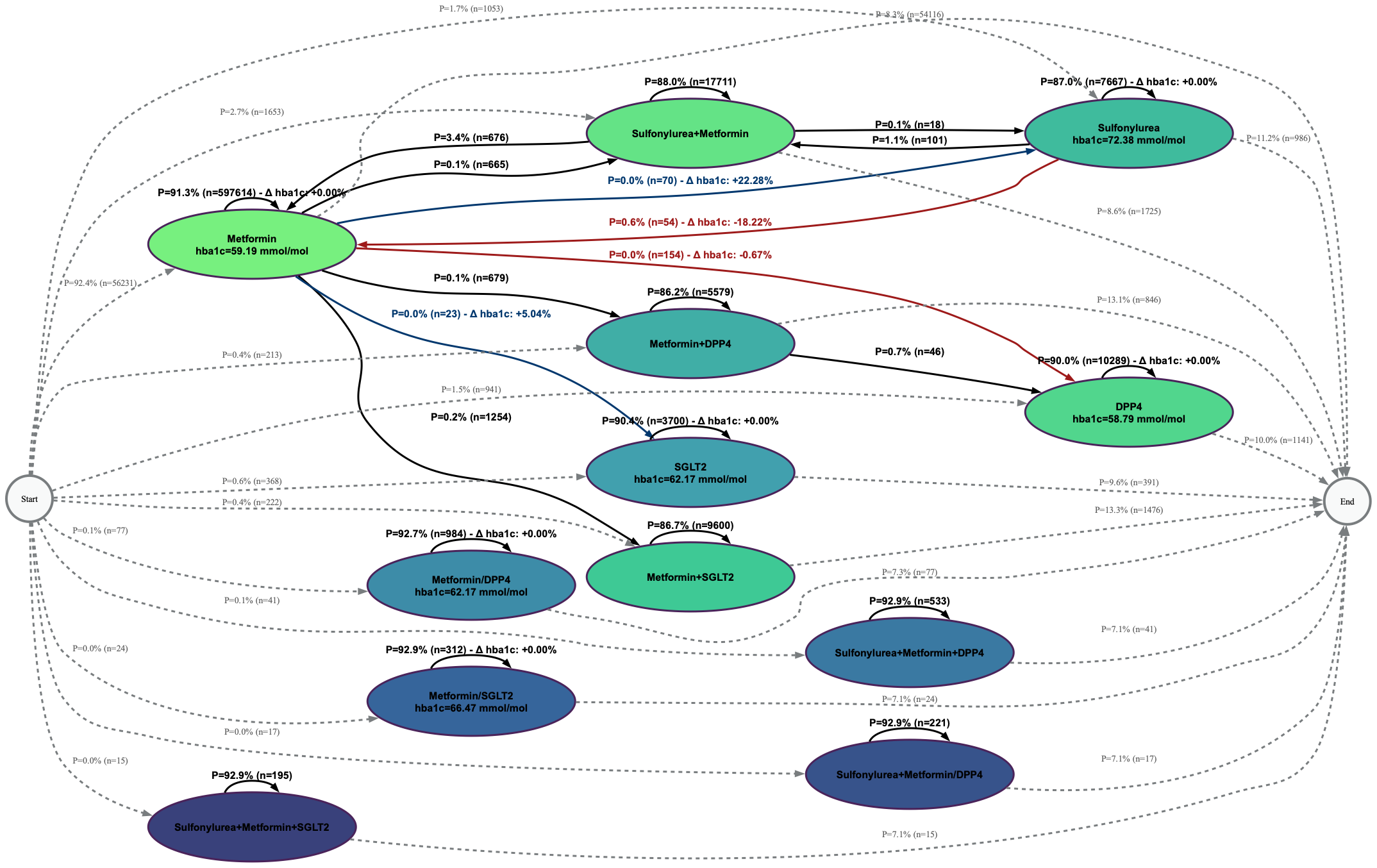

( c) eCVD-C, Precision = 0.96, Fitness = 0.99, F-score = 0.98

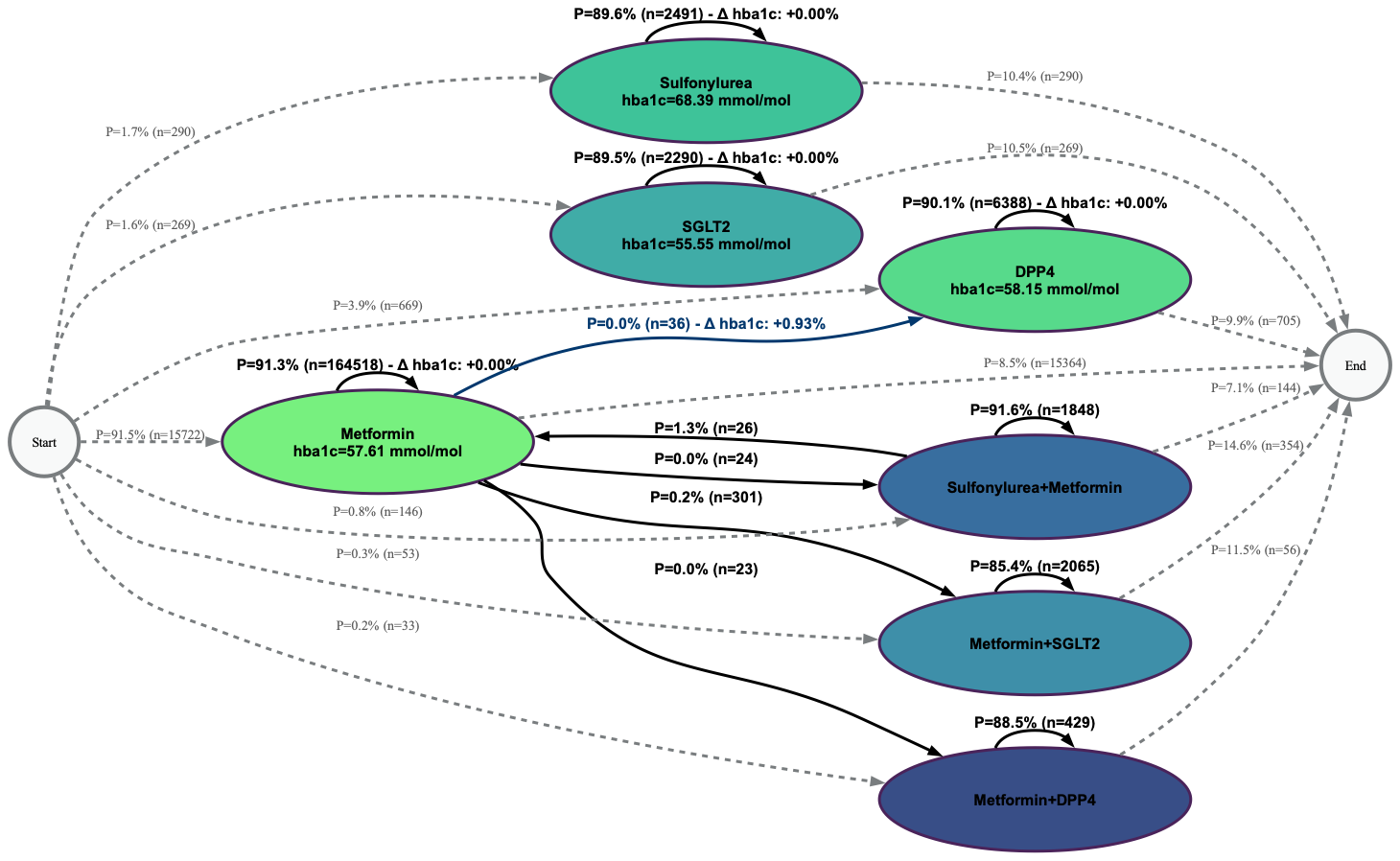

**Figure S2:** First-line treatment and further interventions for patients with T2DM in **during COVID-19**, shown as optimal process maps in the format of Directly-Follows Graph (DFG), in (a) low risk of CVD cohort (LR-C), (b) high risk of CVD cohort (HR-C), and (c) patients with established CVD (eCVD-C).

**2.1 NICE NG28 2022**

NICE T2DM treatment recommendations were reproduced as DFGs stratifying patients into the three CVD risk cohorts (LR-C, HR-C, e-CVD-C), as presented in Figure S2.

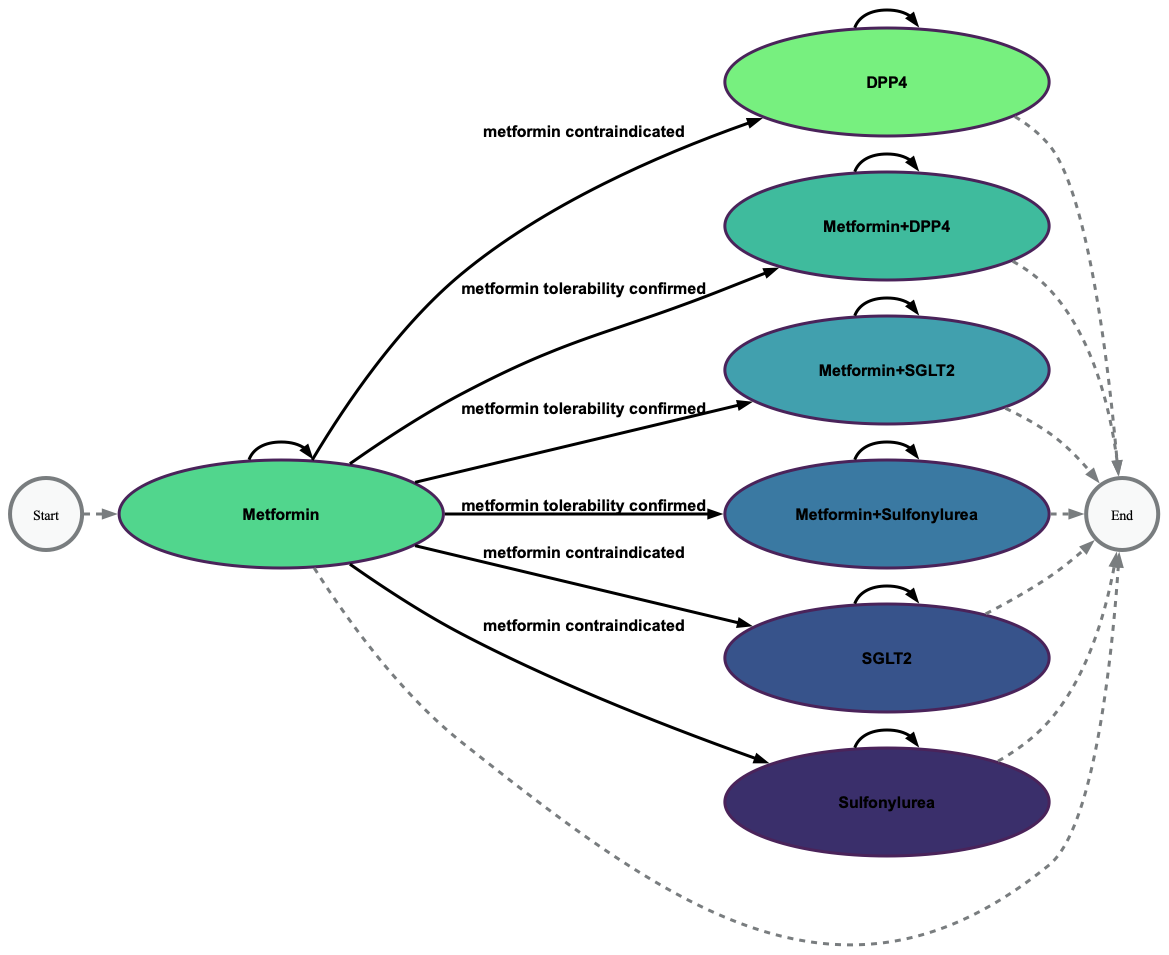

(a)

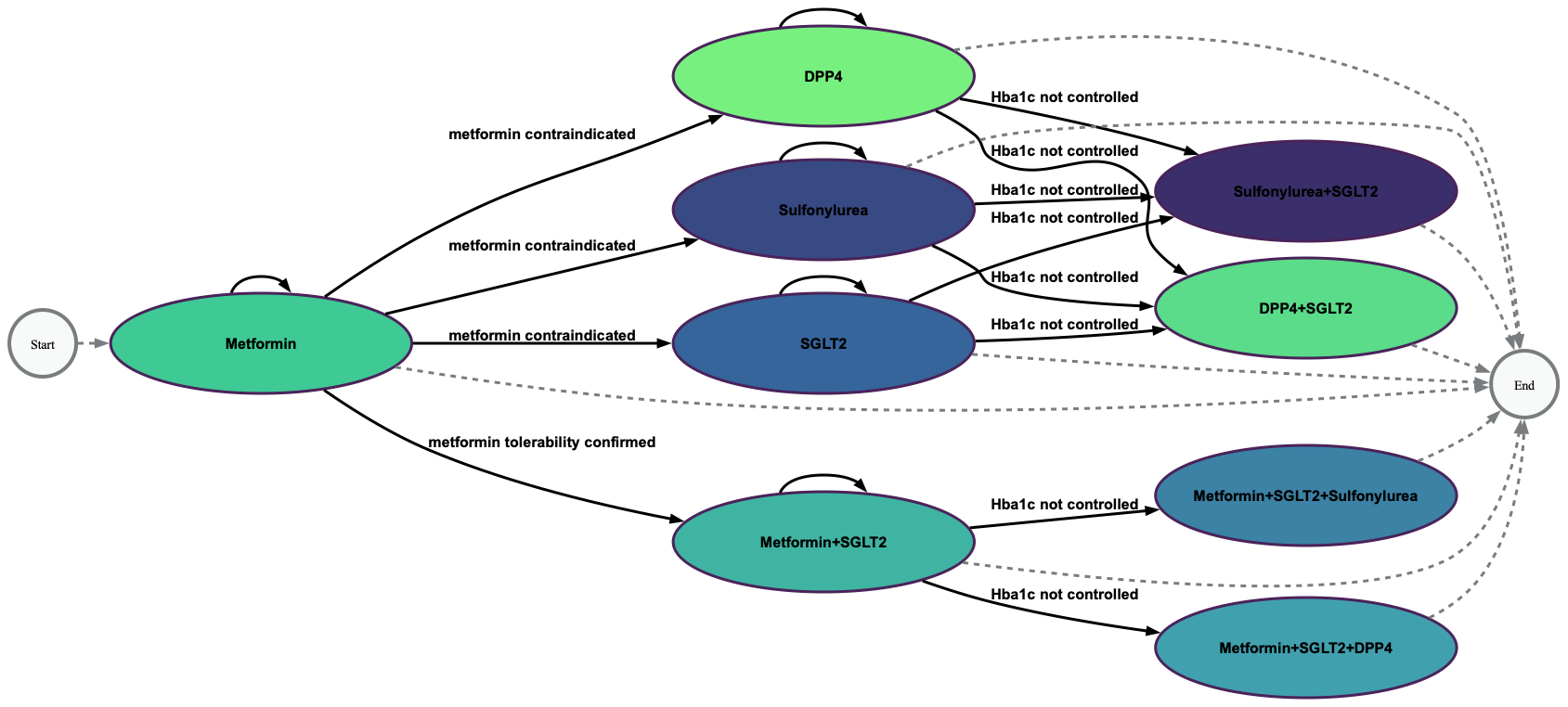

(b)

**Fig. S3**: NICE guidelines represented as a directly follow graph; first line treatments followed by intensification of medicines for patients with T2DM **(a)** at low risk of CVD and **(b)** at high risk of CVD or with established CVD [5].

#### 2.1 Phase 1: national level analysis

Figure S3 represents the changes in drug use in 1-Feb-2022 – 1-Nov-2025. To show the changes in treatment of the whole population in each cohort, alluvial plots were generated to show the switching and the intensification in each 3 month of treatment. Figures S2-S4 represent the changes in treatment in each 3- month duration for all the individuals in the post COVID-19 period.

(a) LR-C – Metformin initiation (90.56%)
(b)
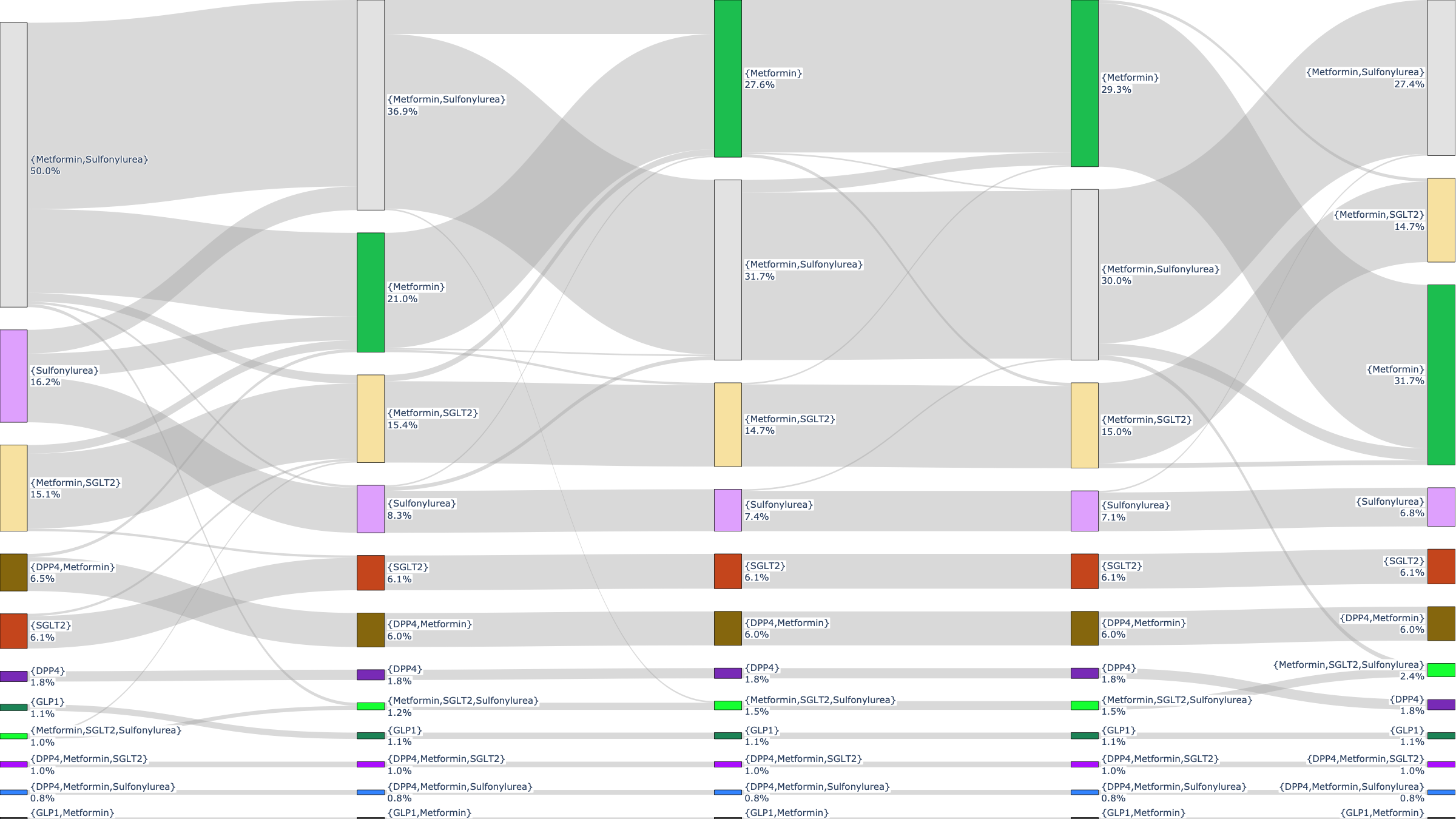

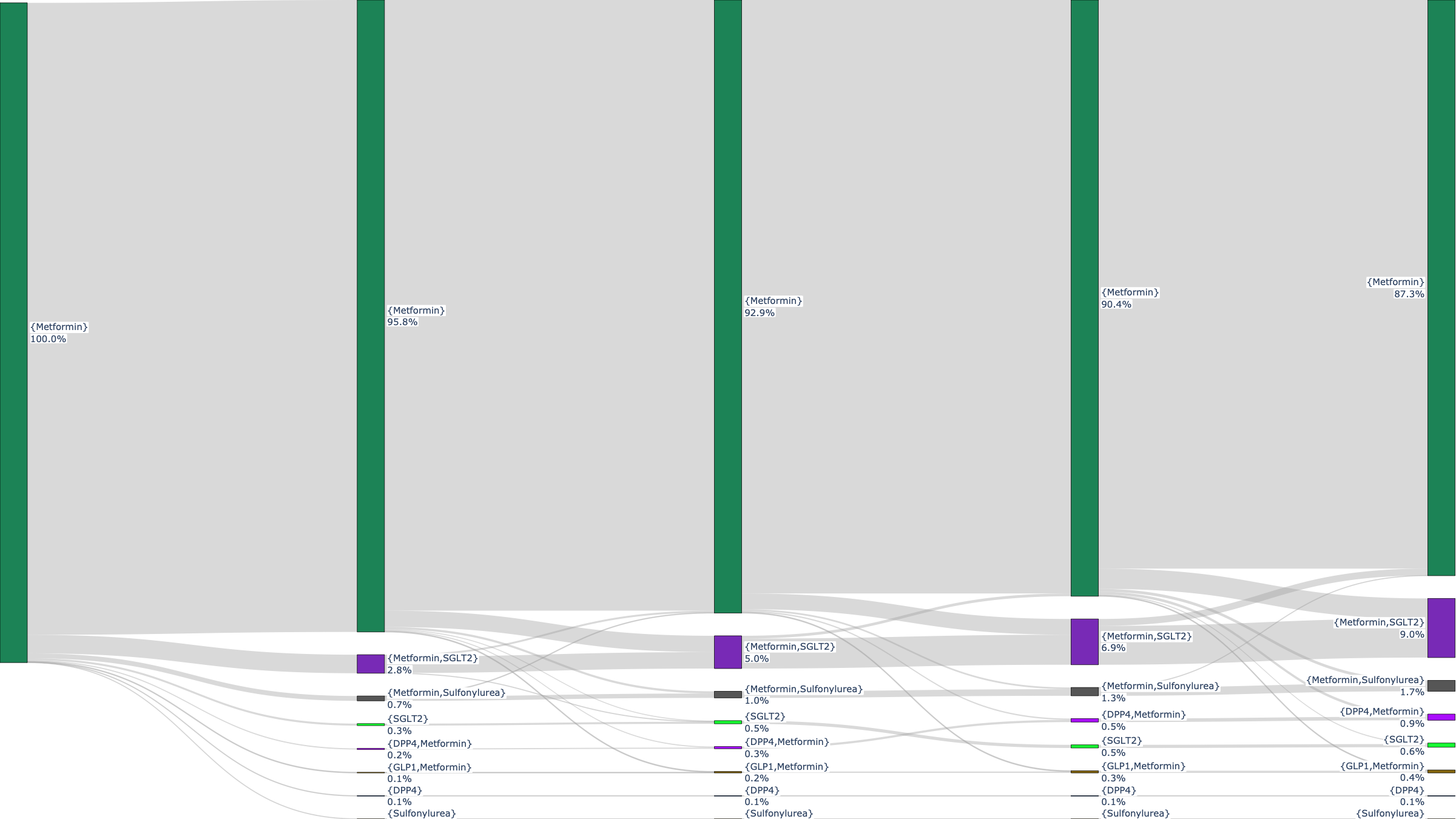
LR-C – Other drugs initiation (9.44%)

**Fig. S4**: Treatment switch and intensification of patients in LR-C in post pandemic period. a) represents the trajectory of treatment changes for those who initiated with metformin. b) represents the trajectory of treatment changes for those who initiated with any other drug.

(a) HR-C – Metformin initiation (91.07%)

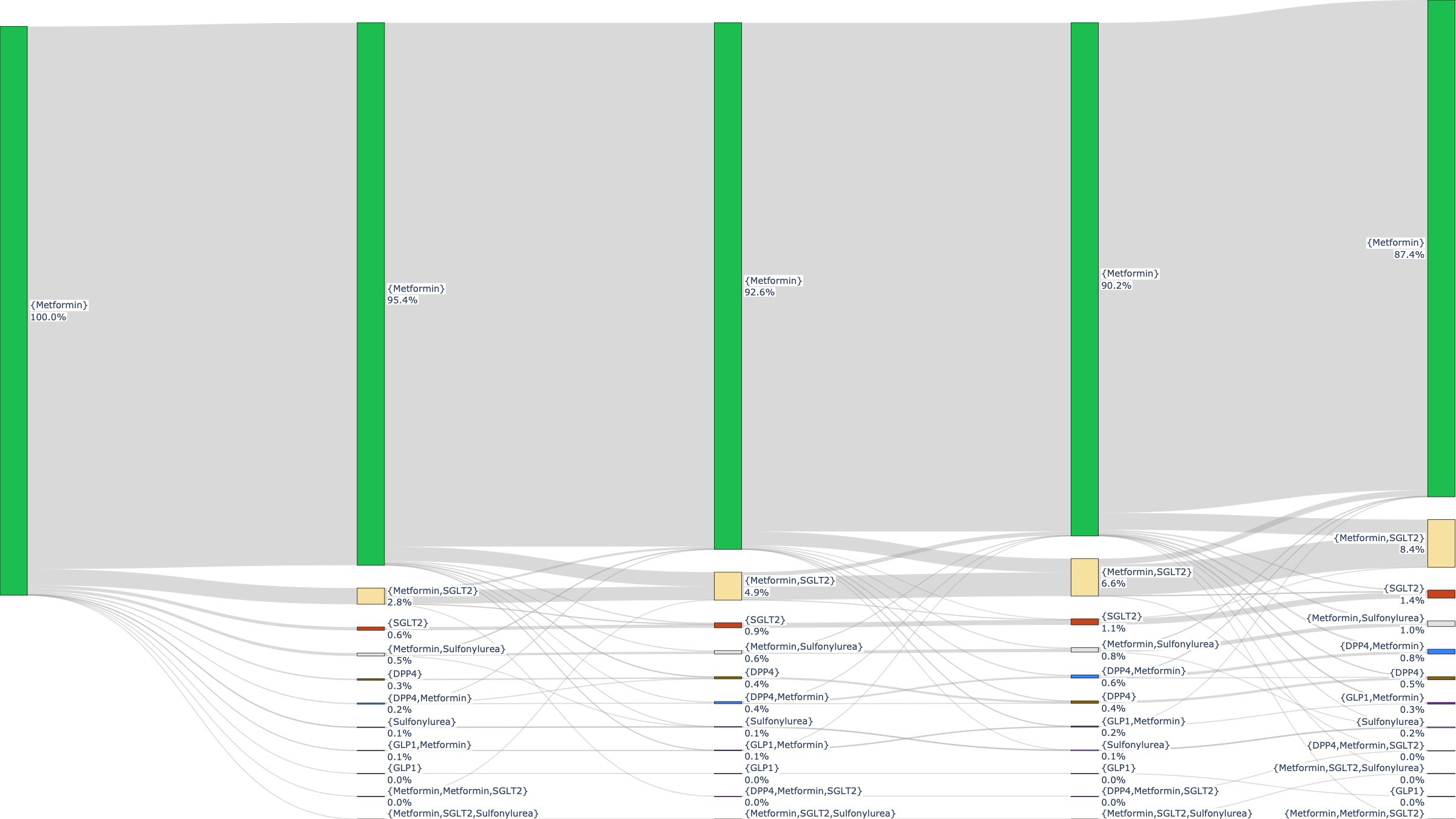

(b)
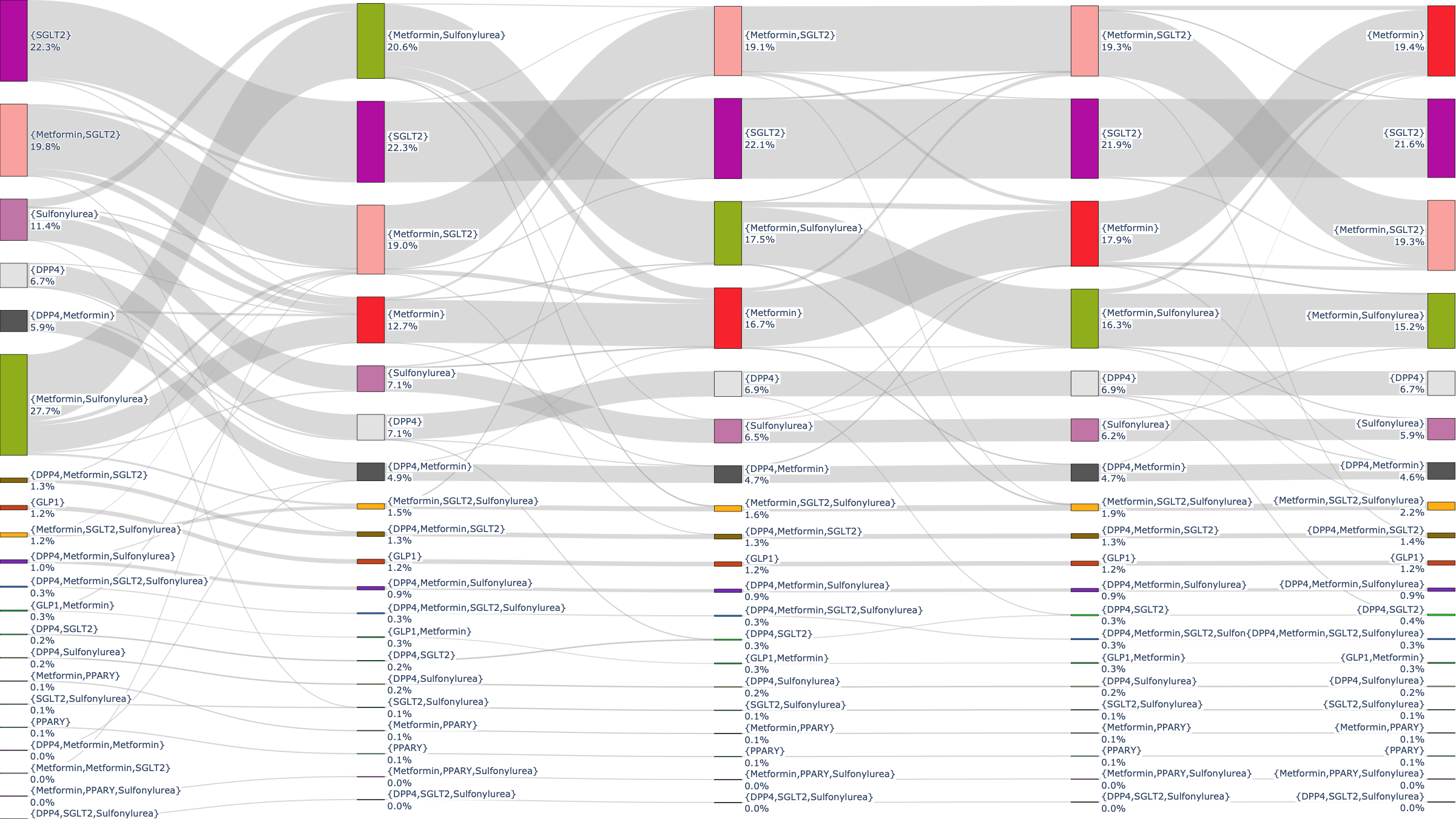
HR-C – Other drugs initiation (8.93%)

**Fig. S5**: Treatment switch and intensification of patients in HR-C in post pandemic period. a) represents the trajectory of treatment changes for those who initiated with metformin. b) represents the trajectory of treatment changes for those who initiated with any other drug.

(a)
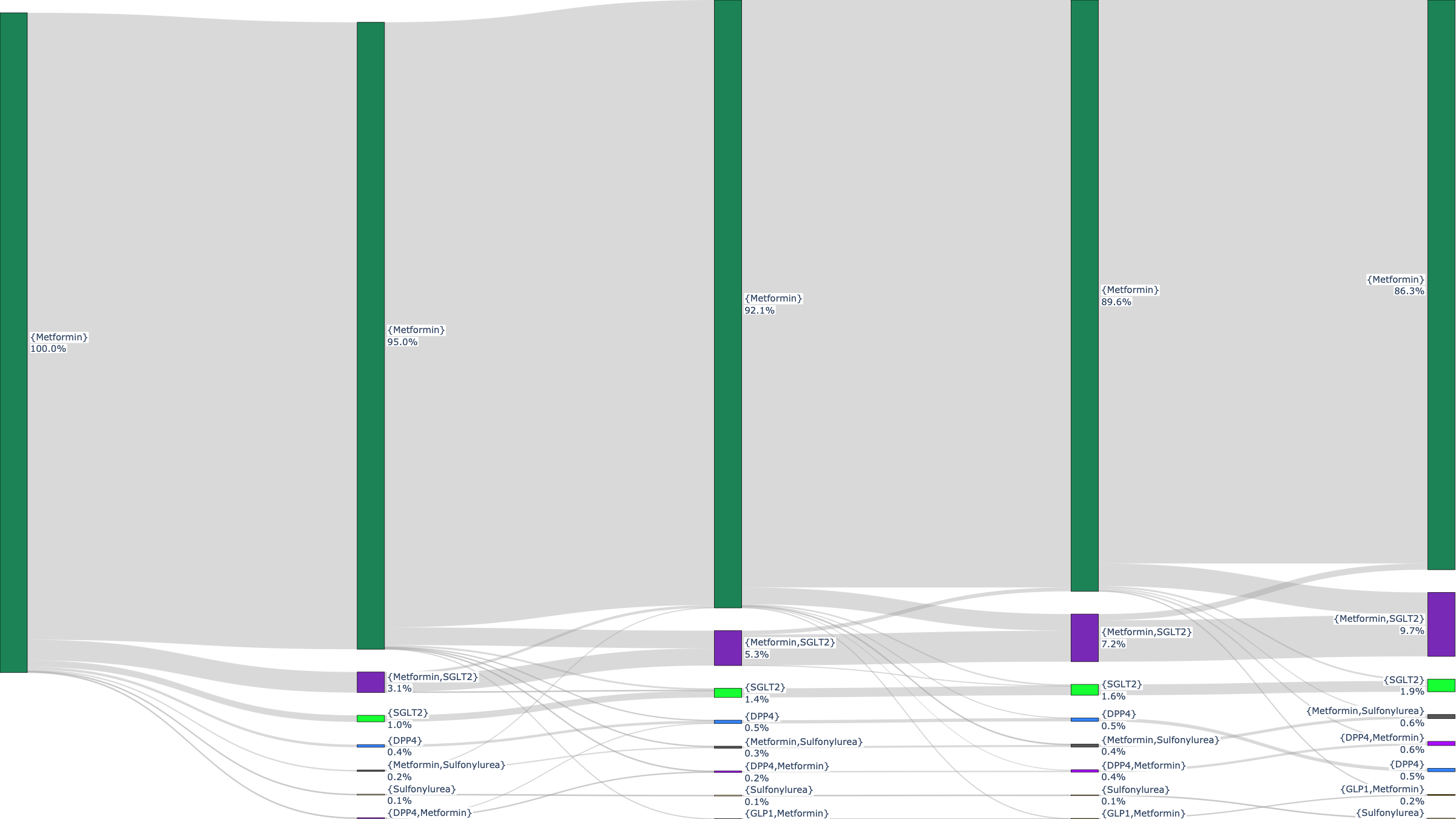
eCVD-C – Metformin initiation (77.97%)
(b)
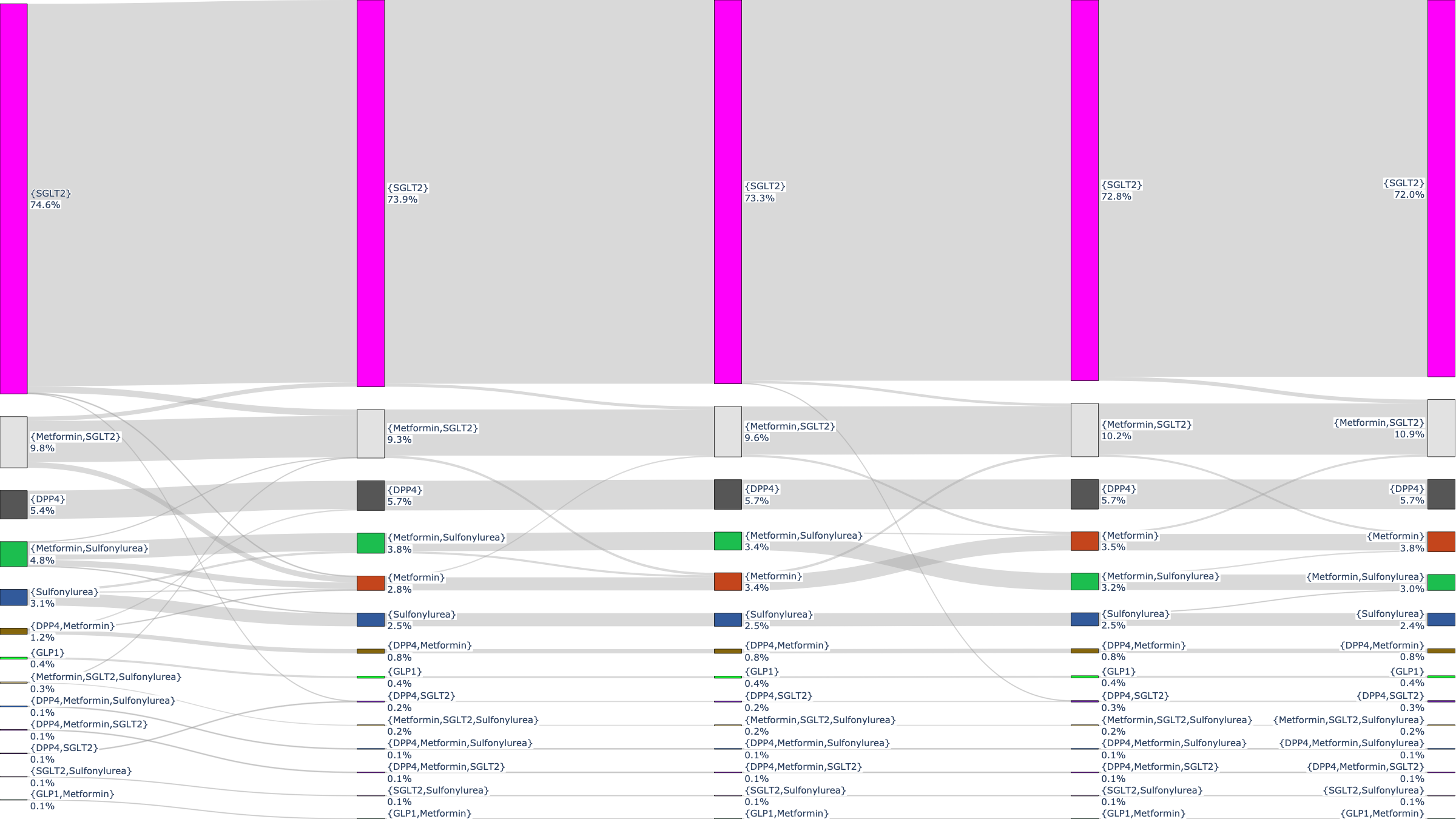
eCVD-C – Other drugs initiation (22.03%)

**Fig. S6**: Treatment switch and intensification of patients in eCVD-C in post pandemic period. a) represents the trajectory of treatment changes for those who initiated with metformin. b) represents the trajectory of treatment changes for those who initiated with any other drug.

#### 2.2 Phase 2: Post-pandemic variation in treatment pathways across regions

**Table S6**: The count and age range of patients with T2DM for the three cardiovascular risk groups, according to NICE [5], and the F-score of the resulted DFG, in each ICB, post Covid Period.

|  |  | | **LR-C** | | | | | | **HR-C** | | | | | | **eCVD-C** | | | | | |
| --- | --- | --- | --- | --- | --- | --- | --- | --- | --- | --- | --- | --- | --- | --- | --- | --- | --- | --- | --- | --- |
| No. | Name | | Count | | Mean Age  (±std) | | F-score | | Count | | Mean Age  (±std) | | F-score | | Count | | Mean Age  (±std) | | F-score | |
| 1 | QHM | | 5435 | | 41.74 (±8.04) | | 0.95 | | 31205 | | 59.74 (±11.30) | | 0.98 | | 10940 | | 67.00 (±10.22) | | 0.98 | |
| 2 | QE1 | | 2615 | | 41.64 (±8.31) | | 0.95 | | 16980 | | 59.22 (±11.46) | | 0.97 | | 6015 | | 67.39 (±10.51) | | 0.98 | |
| 3 | QOQ | | 2995 | | 43.22 (±8.55) | | 0.96 | | 16335 | | 61.10 (±11.15) | | 0.98 | | 5760 | | 68.17 (±10.03) | | 0.98 | |
| 4 | QWO | | 4810 | | 40.94 (±8.00) | | 0.95 | | 24695 | | 58.33 (±11.50) | | 0.98 | | 7685 | | 66.27 (±10.98) | | 0.98 | |
| 5 | QOP | | 5870 | | 41.45 (±7.94) | | 0.95 | | 29795 | | 57.79 (±11.44) | | 0.97 | | 9230 | | 65.96 (±10.77) | | 0.96 | |
| 6 | QF7 | | 2855 | | 41.36 (±8.05) | | 0.95 | | 13505 | | 58.86 (±11.47) | | 0.98 | | 4660 | | 66.38 (±10.63) | | 0.97 | |
| 7 | QYG | | 3780 | | 42.11 (±8.34) | | 0.94 | | 23155 | | 59.46 (±11.30) | | 0.97 | | 8465 | | 67.19 (±10.42) | | 0.97 | |
| 8 | QT1 | | 2265 | | 42.50 (±8.29) | | 0.96 | | 9915 | | 59.46 (±11.29) | | 0.98 | | 3275 | | 67.23 (±10.79) | | 0.97 | |
| 9 | QJM | | 1290 | | 42.72 (±8.28) | | 0.97 | | 8035 | | 61.09 (±11.21) | | 0.97 | | 2830 | | 68.23 (±10.14) | | 0.97 | |
| 10 | QJ2 | | 1970 | | 43.00 (±8.31) | | 0.93 | | 9880 | | 60.12 (±11.22) | | 0.97 | | 3425 | | 67.43 (±10.64) | | 0.98 | |
| 11 | QNC | | 2045 | | 42.52 (±8.35) | | 0.96 | | 11530 | | 60.27 (±11.51) | | 0.97 | | 3685 | | 68.04 (±10.40) | | 0.97 | |
| 12 | QK1 | | 2915 | | 42.52 (±8.17) | | 0.94 | | 12720 | | 59.47 (±11.31) | | 0.97 | | 3650 | | 67.30 (±10.68) | | 0.96 | |
| 13 | QMM | | 1765 | | 42.80 (±8.36) | | 0.94 | | 11425 | | 61.85 (±11.44) | | 0.97 | | 3595 | | 68.57 (±10.20) | | 0.96 | |
| 14 | QOC | | 825 | | 42.85 (±8.29) | | 0.93 | | 5105 | | 60.94 (±11.55) | | 0.95 | | 1665 | | 68.76 (±10.04) | | 0.96 | |
| 15 | QUA | | 3105 | | 40.45 (±7.84) | | 0.94 | | 14760 | | 58.03 (±11.51) | | 0.97 | | 4365 | | 65.99 (±10.72) | | 0.97 | |
| 16 | QHL | | 3410 | | 40.20 (±8.08) | | 0.94 | | 15095 | | 56.81 (±11.33) | | 0.96 | | 4110 | | 65.08 (±11.18) | | 0.96 | |
| 17 | QPM | | 1670 | | 43.75 (±8.17) | | 0.95 | | 8325 | | 59.62 (±11.32) | | 0.96 | | 2630 | | 67.27 (±10.49) | | 0.96 | |
| 18 | QWU | | 2155 | | 42.11 (±8.18) | | 0.94 | | 9840 | | 59.10 (±11.76) | | 0.96 | | 2630 | | 67.41 (±10.62) | | 0.96 | |
| 19 | QUE | | 2080 | | 42.85 (±8.18) | | 0.96 | | 8165 | | 59.85 (±11.39) | | 0.96 | | 2600 | | 67.32 (±10.64) | | 0.97 | |
| 20 | QGH | | 1250 | | 43.43 (±8.44) | | 0.92 | | 7190 | | 61.18 (±11.38) | | 0.96 | | 2330 | | 68.97 (±10.29) | | 0.95 | |
| 21 | QJG | | 1800 | | 43.05 (±8.39) | | 0.94 | | 9550 | | 61.46 (±11.44) | | 0.97 | | 3380 | | 68.62 (±10.38) | | 0.97 | |
| 22 | QHG | | 2950 | | 42.23 (±8.43) | | 0.93 | | 12545 | | 58.86 (±11.57) | | 0.97 | | 3665 | | 66.48 (±11.05) | | 0.95 | |
| 23 | QH8 | | 2100 | | 43.69 (±8.36) | | 0.94 | | 10975 | | 60.49 (±11.53) | | 0.97 | | 3225 | | 67.75 (±10.51) | | 0.96 | |
| 24 | QM7 | | 2655 | | 43.81 (±8.33) | | 0.95 | | 11970 | | 60.03 (±11.26) | | 0.97 | | 3545 | | 67.80 (±10.51) | | 0.96 | |
| 25 | QR1 | | 1175 | | 43.43 (±8.49) | | 0.95 | | 5780 | | 61.38 (±11.19) | | 0.97 | | 1945 | | 67.99 (±10.66) | | 0.96 | |
| 26 | QU9 | | 3475 | | 43.96 (±8.46) | | 0.95 | | 14055 | | 60.31 (±11.64) | | 0.97 | | 4050 | | 68.04 (±10.76) | | 0.97 | |
| 27 | | QMJ | | 2540 | | 42.96 (±8.48) | | 0.95 | | 11625 | | 58.55 (±11.07) | | 0.96 | | 2840 | | 65.80 (±10.90) | | 0.96 |
| 28 | | QMF | | 6030 | | 40.76 (±8.33) | | 0.96 | | 24540 | | 56.32 (±11.23) | | 0.98 | | 5080 | | 63.94 (±11.30) | | 0.96 |
| 29 | | QRV | | 6175 | | 41.62 (±8.22) | | 0.94 | | 24415 | | 57.59 (±11.27) | | 0.97 | | 5530 | | 65.28 (±10.93) | | 0.96 |
| 30 | | QUY | | 1780 | | 42.92 (±8.45) | | 0.95 | | 7280 | | 59.98 (±11.47) | | 0.96 | | 2685 | | 67.69 (±10.68) | | 0.98 |
| 31 | | QKK | | 4080 | | 43.81 (±8.72) | | 0.94 | | 16285 | | 59.16 (±10.80) | | 0.98 | | 3770 | | 65.60 (±10.71) | | 0.97 |
| 32 | | QNQ | | 1725 | | 42.87 (±8.59) | | 0.92 | | 7480 | | 58.93 (±11.64) | | 0.96 | | 1860 | | 67.35 (±10.76) | | 0.97 |
| 33 | | QWE | | 3050 | | 43.12 (±8.52) | | 0.95 | | 12380 | | 59.10 (±11.07) | | 0.96 | | 2905 | | 65.97 (±10.93) | | 0.96 |
| 34 | | QOX | | 1690 | | 43.49 (±8.49) | | 0.92 | | 7770 | | 61.00 (±11.13) | | 0.97 | | 2440 | | 68.22 (±10.28) | | 0.94 |
| 35 | | QXU | | 1570 | | 44.13 (±8.34) | | 0.94 | | 7230 | | 60.75 (±11.40) | | 0.97 | | 2320 | | 68.50 (±10.34) | | 0.98 |
| 36 | | QKS | | 3235 | | 43.04 (±8.21) | | 0.95 | | 17325 | | 60.14 (±11.39) | | 0.98 | | 5135 | | 67.64 (±10.64) | | 0.96 |
| 37 | | QRL | | 3325 | | 43.26 (±8.44) | | 0.96 | | 15425 | | 60.74 (±11.46) | | 0.97 | | 5280 | | 68.33 (±10.36) | | 0.97 |
| 38 | | QSL | | 815 | | 42.72 (±7.88) | | 0.92 | | 4920 | | 61.96 (±11.33) | | 0.97 | | 1945 | | 69.34 (±9.83) | | 0.98 |
| 39 | | QNX | | 2535 | | 43.04 (±8.51) | | 0.94 | | 14395 | | 61.10 (±11.44) | | 0.97 | | 4820 | | 68.36 (±10.57) | | 0.95 |
| 40 | | QVV | | 1115 | | 43.60 (±8.52) | | 0.94 | | 5985 | | 61.85 (±11.18) | | 0.97 | | 2415 | | 69.45 (±9.80) | | 0.96 |
| 41 | | QJK | | 1640 | | 42.76 (±8.39) | | 0.97 | | 9500 | | 61.40 (±11.26) | | 0.98 | | 3390 | | 68.89 (±9.90) | | 0.96 |
| 42 | | QT6 | | 635 | | 42.97 (±7.93) | | 0.97 | | 4485 | | 61.84 (±11.04) | | 0.97 | | 1985 | | 68.79 (±9.89) | | 0.98 |

The total percentage of dispensed drugs in each ICB per Cardiovascular risk group is shown in Figure S5. These figures show that the percentage of SGLT2 prescription were higher for eCVD cohort, overall, compared to LR-C and HR-C. Moreover, it is important to emphasise that, despite the high performance of the DFGs, the models do not capture the complete treatment pathway for every patient, but instead, they represent the most common treatment pathways of most patients.

(a) LR-C

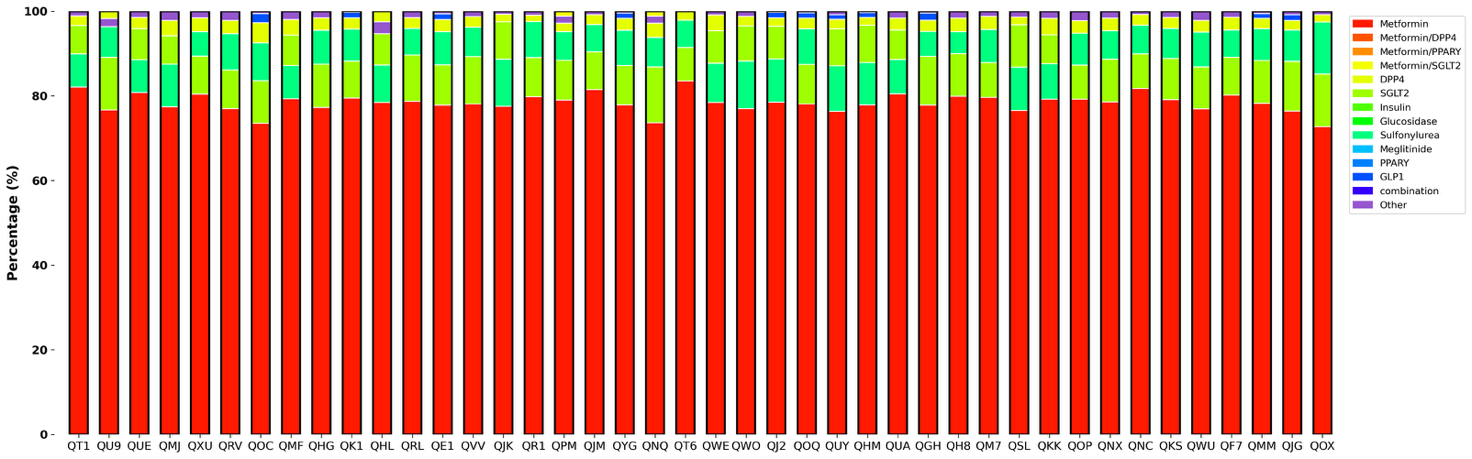

(b) HR-C

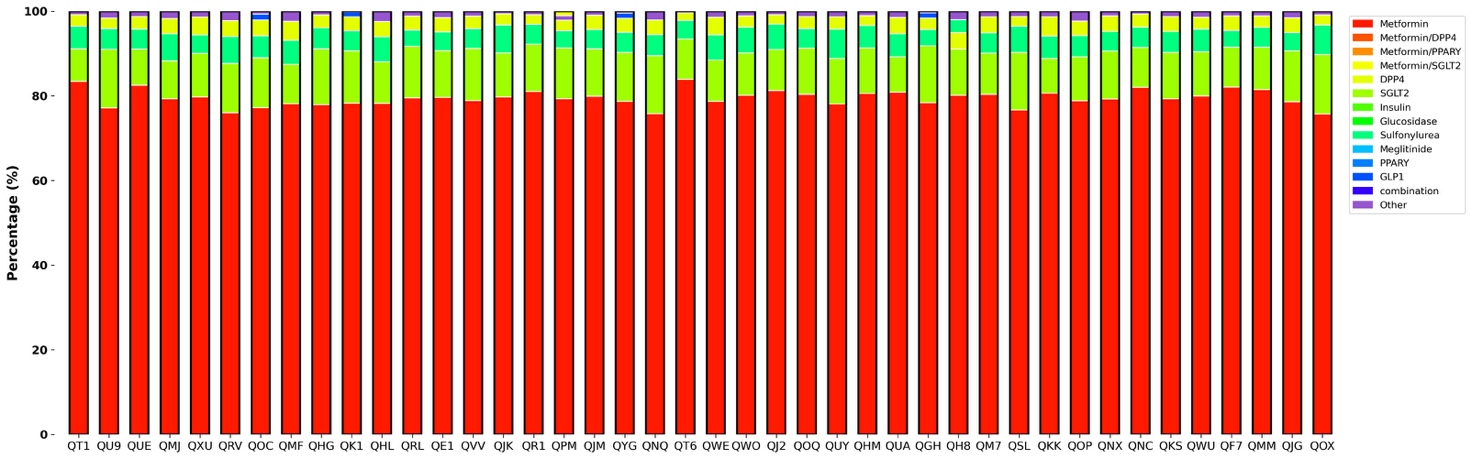

(c) eCVD-C

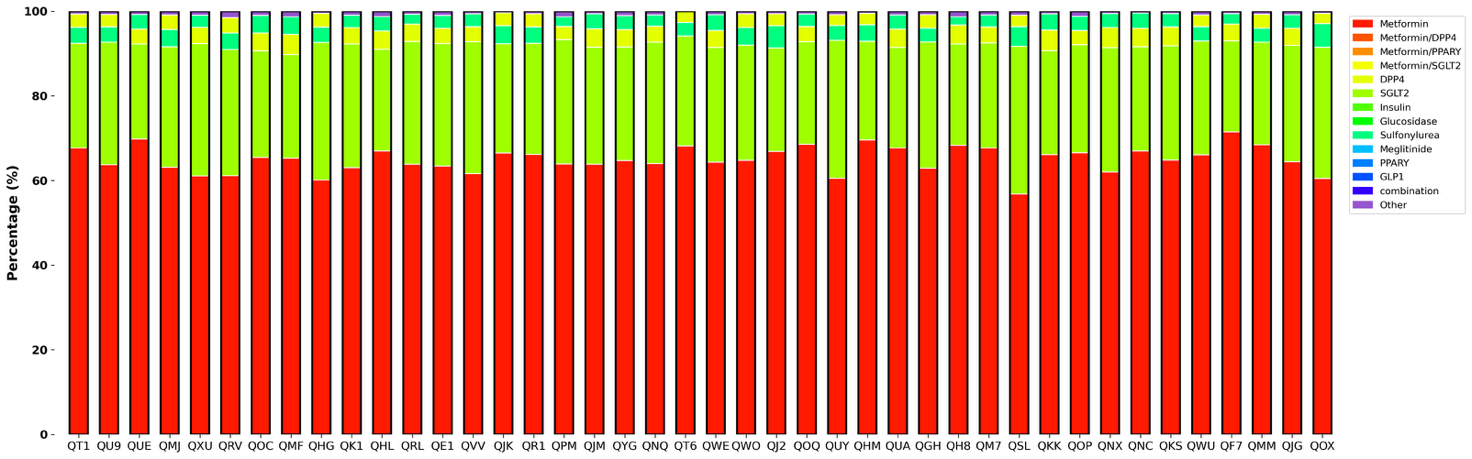

**Fig. S7**: Total percentage of dispensed drugs for the cohort (a) at low risk of CVD, (b) at high risk of CVD, and (c) with eCVD, in each ICB in 1 year after the onset of T2DM.

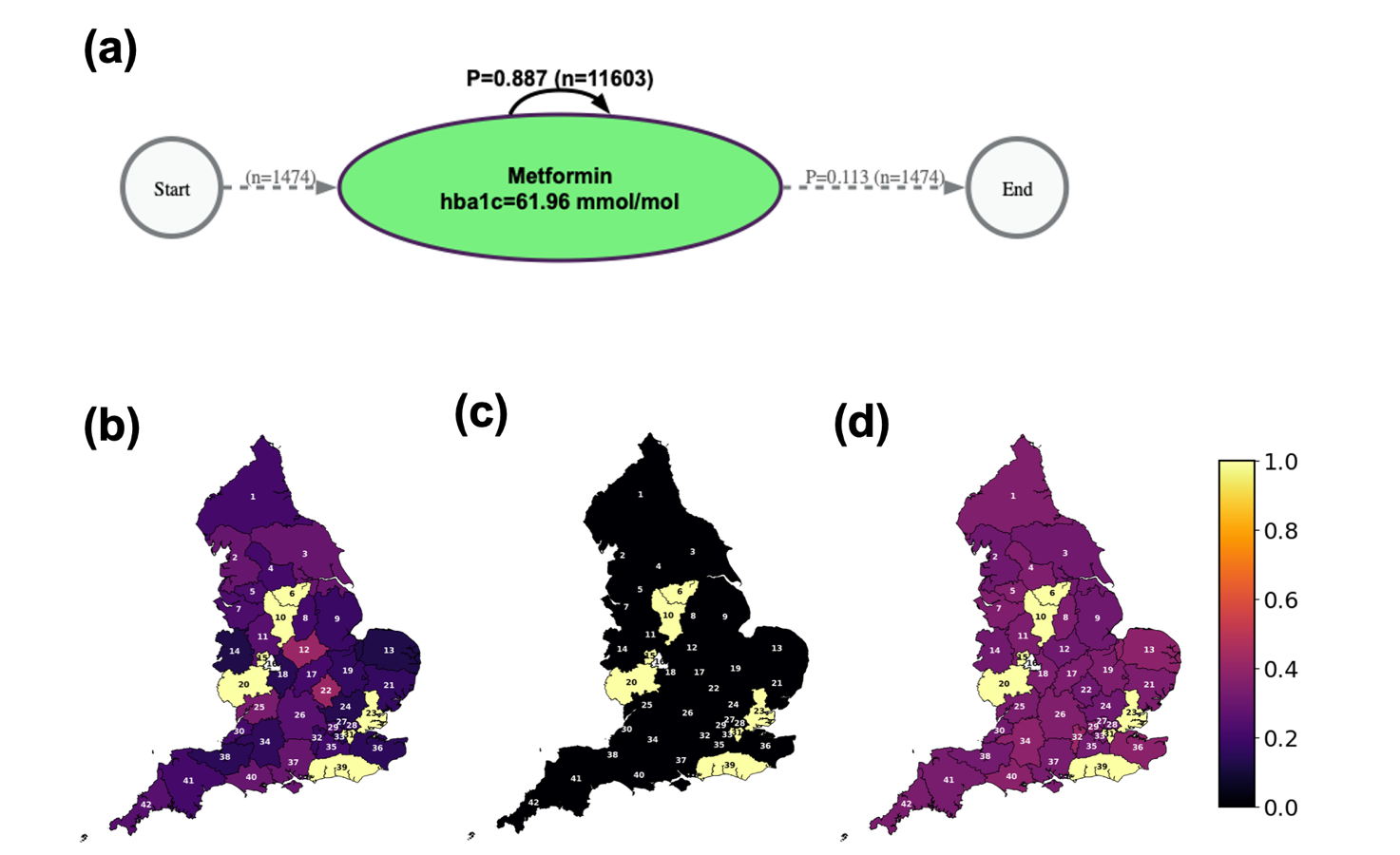

**Fig. S8**: ICB-16 is used as a reference, and the treatment pathways of other ICBs are

compared to the reference. (a) Optimal T2DM treatment pathways for patients in

**LR-C** for ICB-16 (b) Structural, (c) Behavioural, and (d) Entropy-based similarity

index between other ICBs and ICB-16 (shown in white), as reference.

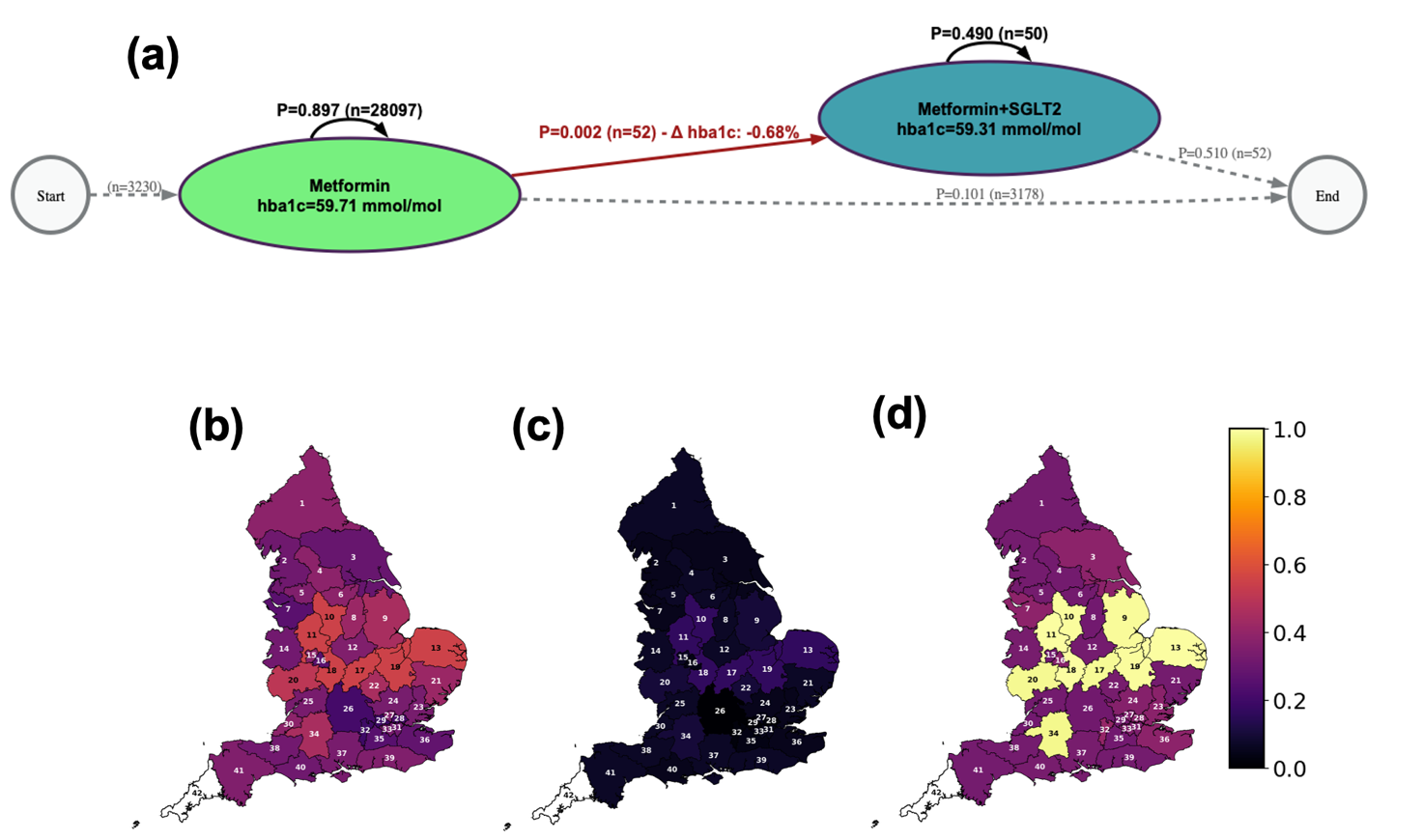

**Fig. S9**: ICB-42 is used as a reference, and the treatment pathways of other ICBs are

compared to the reference. (a) Optimal T2DM treatment pathways for patients in

**HR-C** for ICB-42 (b) Structural, (c) Behavioural, and (d) Entropy-based similarity

index between other ICBs and ICB-42 (shown in white), as reference

### Codelists

**Table S7**: Chronic Heart failure SNOMED and ICD10 code list [1]

|  | 1 | | 10335000 | | Chronic right-sided heart failure (disorder) | | | 33 | 426263006 | | Congestive heart failure due to left ventricular systolic dysfunction (disorder) |
| --- | --- | --- | --- | --- | --- | --- | --- | --- | --- | --- | --- |
|  | 2 | | 10633002 | | Acute congestive heart failure | | | 34 | 426611007 | | Congestive heart failure due to valvular disease (disorder) |
|  | 3 | | 111283005 | | Chronic left-sided heart failure (disorder) | | | 35 | 430396006 | | Chronic systolic dysfunction of left ventricle (disorder) |
|  | 4 | | 1204200007 | | Left ventricular failure with normal ejection fraction due to valvular heart disease  (disorder) | | | 36 | 43736008 | | Rheumatic left ventricular  failure |
|  | 5 | | 1204203009 | | Left ventricular failure with normal ejection fraction due to coronary arteriosclerosis (disorder) | | | 37 | 441481004 | | Chronic systolic heart failure  (disorder) |
|  | 6 | | 1204204003 | | Left ventricular failure with normal ejection fraction due to myocarditis (disorder) | | | 38 | 441530006 | | Chronic diastolic heart failure (disorder) |
|  | 7 | | 1204206001 | | Left ventricular failure with normal ejection fraction due to cardiomyopathy (disorder) | | | 39 | 446221000 | | Heart failure with normal ejection fraction (disorder) |
|  | 8 | | 1204462004 | | Left ventricular failure with sepsis (disorder) | | | 40 | 48447003 | | Chronic heart failure (disorder) |
|  | 9 | | 1204468000 | | Right ventricular failure with sepsis (disorder) | | | 41 | 5375005 | | Chronic left-sided congestive heart failure (disorder) |
|  | 10 | | 1208843003 | | Right ventricular failure due to heart valve disorder (disorder) | | | 42 | 56675007 | | Acute heart failure |
|  | 11 | | 1208846006 | | Right ventricular failure due to disorder of lung (disorder) | | | 43 | 66989003 | | Chronic right-sided congestive heart failure (disorder) |
|  | 12 | | 1208848007 | | Right ventricular failure due to disorder of pulmonary circulation (disorder) | | | 44 | 698592004 | | Asymptomatic left ventricular systolic dysfunction (disorder) |
|  | 13 | | 1208850004 | | Right ventricular failure due to right ventricular infarction (disorder) | | | 45 | 703272007 | | Heart failure with reduced ejection fraction (disorder) |
|  | 14 | | 128404006 | | Right heart failure | | | 46 | 71892000 | | Cardiac asthma |
|  | 15 | | 134401001 | | Left ventricular systolic dysfunction | | | 47 | 788950000 | | Heart failure with mid range ejection fraction (disorder) |
|  | 16 | | 194767001 | | Benign hypertensive heart disease with congestive cardiac failure | | | 48 | 79955004 | | Chronic cor pulmonale |
|  | 17 | | 194779001 | | Hypertensive heart and renal disease with (congestive) heart failure | | | 49 | 82523003 | | Congestive rheumatic heart failure (disorder) |
|  | 18 | | 194781004 | | Hypertensive heart and renal disease with both (congestive) heart failure and renal  failure | | | 50 | 83105008 | | Malignant hypertensive heart disease with congestive heart failure |
|  | 19 | | 195111005 | | Decompensated cardiac failure | | | 51 | 84114007 | | Heart failure |
|  | 20 | | 195112003 | | Compensated cardiac failure | | | 52 | 85232009 | | Left heart failure |
|  | 21 | | 195114002 | | Acute left ventricular failure | | | 53 | 871617000 | | Low output heart failure due to and following Fontan operation (disorder) |
|  | 22 | | 206586007 | | Congenital cardiac failure | | | 54 | 87837008 | | Chronic pulmonary heart  disease |
|  | 23 | | 233924009 | | Heart failure as a complication of care (disorder) | | | 55 | 88805009 | | Chronic congestive heart  failure |
| 24 | | 275514001 | | Impaired left ventricular function | | 56 | 898208007 | | | Heart failure due to thyrotoxicosis (disorder) | |
| 25 | | 314206003 | | Refractory heart failure (disorder) | | 57 | 92506005 | | | Biventricular congestive heart failure | |
| 26 | | 367363000 | | Right ventricular failure | | 58 | 153941000119100 | | | Chronic combined systolic and diastolic heart failure (disorder) | |
| 27 | | 407596008 | | Echocardiogram shows left ventricular systolic dysfunction (finding) | | 59 | 16838951000119100 | | | Acute on chronic right-sided congestive heart failure (disorder) | |
| 28 | | 420300004 | | New York Heart Association Classification - Class I (finding) | | 60 | I11.0 | | | Hypertensive heart disease with (congestive) heart failure | |
| 29 | | 420913000 | | New York Heart Association Classification - Class III (finding) | | 61 | I13.0 | | | Hypertensive heart and renal disease with (congestive) heart failure | |
| 30 | | 421704003 | | New York Heart Association Classification - Class II (finding) | | 62 | I13.2 | | | Hypertensive heart and renal disease with both (congestive) heart failure and renal  failure | |
| 31 | | 422293003 | | New York Heart Association Classification - Class IV (finding) | | 63 | I50 | | | Heart failure | |
| 32 | | 42343007 | | Congestive heart failure | |  |  | | |  | |

**Table S8**: Established atherosclerotic CVD SNOMED and ICD10 code list

|  | **Coronary heart disease** | | | | |
| --- | --- | --- | --- | --- | --- |
| 1 | 46109009 | Subendocardial ischemia | 19 | 472100003 | Ischemic dilated cardiomyopathy due to coronary  artery disease (disorder) |
| 2 | 53741008 | Coronary arteriosclerosis | 20 | 712866001 | Resting ischaemia |
| 3 | 67682002 | Coronary artery atheroma | 21 | 15629741000119100 | Systolic heart failure stage C due to ischaemic cardiomyopathy |
| 4 | 171223006 | Ischemic heart disease  screening | 22 | 15629641000119100 | Systolic heart failure stage B due to ischemic cardiomyopathy (disorder) |
| 5 | 194821006 | Coronary thrombosis not resulting in myocardial  infarction | 23 | 15629591000119100 | Congestive heart failure stage B due to ischemic  cardiomyopathy (disorder) |
| 6 | 194849004 | Generalized ischemic  myocardial dysfunction | 24 | 15629541000119100 | Congestive heart failure stage C due to Ischemic  cardiomyopathy (disorder) |
| 7 | 233817007 | Triple vessel disease of the heart | 25 | 15960061000119100 | Unstable angina cooccurrent and due to coronary arteriosclerosis (disorder) |
| 8 | 233823002 | Silent myocardial ischemia | 26 | 15960141000119100 | Angina co-occurrent and due to coronary arteriosclerosis (disorder) |
| 9 | 275931000 | Aspirin prophylaxis - IHD | 27 | 16754391000119100 | Stable angina due to coronary arteriosclerosis (disorder) |
| 10 | 315026000 | Transient myocardial  ischaemia | 28 | I250 | Atherosclerotic cardiovascular disease, so described |
| 11 | 315348000 | Asymptomatic coronary  heart disease | 29 | I251 | Atherosclerotic heart disease |
| 12 | 315610002 | Primary prevention of  ischemic heart disease | 30 | I253 | Aneurysm of heart |
| 13 | 414024009 | Disorder of coronary artery  (disorder) | 31 | I254 | Coronary artery aneurysm |
| 14 | 414545008 | Ischemic heart disease (disorder) | 32 | I255 | Ischaemic cardiomyopathy |

| 15 | 413838009 | Chronic ischemic heart disease (disorder) | 33 | I256 | Silent myocardial ischaemia |
| --- | --- | --- | --- | --- | --- |
| 16 | 413844008 | Chronic myocardial ischemia  (disorder) | 34 | I258 | Other forms of chronic  ischaemic heart disease |
| 17 | 426579008 | Coronary heart disease education (procedure) | 35 | I259 | Chronic ischaemic heart disease, unspecified |
| 18 | 443502000 | Atherosclerosis of coronary artery (disorder) |  |  |  |
| **Acute coronary syndrome** | | | | | |
| 1 | 394659003 | Acute coronary syndrome  (disorder) | 3 | 394659003 | Acute coronary syndrome  (disorder) |
| 2 | 837091000000100 | History of acute coronary syndrome (situation) | 4 | 16025471000119100 | Aneurysm of coronary artery due to and following acute febrile mucocutaneous lymph node syndrome  (disorder) |
| **Myocardial infarction** [2] | | | | | |
| 1 | 10273003 | Acute infarction of papillary muscle | 53 | 44821000087100 | Acute ST segment elevation myocardial infarction due to occlusion of mid portion of circumflex branch of left coronary artery (disorder) |
| 2 | 1077002 | Septal infarction by electrocardiogram (finding) | 54 | 44831000087103 | Acute ST segment elevation myocardial infarction due to occlusion of obtuse marginal branch of circumflex branch of left coronary artery (disorder) |
| 3 | 1089451000000100 | Acute nontransmural myocardial infarction  (disorder) | 55 | 44841000087109 | Acute ST segment elevation myocardial infarction due to occlusion of posterior lateral branch of circumflex branch of left coronary artery (disorder) |
| 4 | 1163440003 | Postoperative acute myocardial infarction (disorder) | 56 | 44851000087107 | Acute ST segment elevation myocardial infarction due to occlusion of proximal portion of circumflex branch of left coronary artery (disorder) |
| 5 | 1204151009 | Acute inferior non-ST segment elevation myocardial infarction of right ventricle (disorder) | 57 | 52035003 | Acute anteroapical myocardial infarction |
| 6 | 1204152002 | Acute inferior non-ST segment elevation myocardial infarction (disorder) | 58 | 54329005 | Acute myocardial infarction of anterior wall |
| 7 | 1204154001 | Acute anterior non-ST segment elevation myocardial infarction with right ventricular involvement (disorder) | 59 | 57054005 | Acute myocardial infarction |
| 8 | 1204155000 | Acute anterior non-ST segment elevation myocardial infarction (disorder) | 60 | 58612006 | Acute myocardial infarction of lateral wall |
| 9 | 1204222000 | Acute non-ST segment elevation myocardial infarction of right ventricle (disorder) | 61 | 59063002 | Acute myocardial infarction of apical-lateral wall (disorder) |
| 10 | 1208873007 | Subsequent inferior non-ST segment elevation myocardial infarction (disorder) | 62 | 62695002 | Acute anteroseptal myocardial infarction |
| 11 | 129574000 | Postoperative myocardial  infarction | 63 | 64627002 | Acute myocardial infarction of high lateral wall (disorder) |
| 12 | 15712841000119100 | Acute ST segment elevation myocardial infarction of posterolateral wall (disorder) | 64 | 65547006 | Acute myocardial infarction of inferolateral wall |

| 13 | 15713001000119100 | Acute ST segment elevation myocardial infarction of atrium (disorder) | 65 | 70211005 | Acute myocardial infarction of anterolateral wall |
| --- | --- | --- | --- | --- | --- |
| 14 | 15713161000119100 | Acute ST segment elevation myocardial infarction of septum (disorder) | 66 | 703164000 | Acute ST segment elevation myocardial infarction of anterior wall (disorder) |
| 15 | 15990001 | Acute myocardial infarction of posterolateral wall | 67 | 703165004 | Acute ST segment elevation myocardial infarction of anterior wall involving right ventricle (disorder) |
| 16 | 17531000119105 | Acute myocardial infarction due to left coronary artery  occlusion (disorder) | 68 | 703209002 | Subsequent ST segment elevation myocardial infarction of inferior wall (disorder) |
| 17 | 194802003 | True posterior myocardial  infarction | 69 | 703210007 | Subsequent ST segment elevation myocardial infarction of anterior wall (disorder) |
| 18 | 194809007 | Acute atrial infarction | 70 | 703211006 | Subsequent ST segment elevation myocardial infarction (disorder) |
| 19 | 194856005 | Subsequent myocardial  infarction | 71 | 703212004 | Acute myocardial infarction during procedure (disorder) |
| 20 | 194857001 | Subsequent myocardial  infarction of anterior wall | 72 | 703213009 | Acute ST segment elevation myocardial infarction of inferior wall (disorder) |
| 21 | 194858006 | Subsequent myocardial  infarction of inferior wall | 73 | 703251009 | Acute myocardial infarction of inferior wall involving right ventricle (disorder) |
| 22 | 22298006 | Myocardial infarction | 74 | 703252002 | Acute myocardial infarction of anterior wall involving right ventricle (disorder) |
| 23 | 23311000119105 | Acute myocardial infarction due to right coronary artery occlusion (disorder) | 75 | 703253007 | Acute ST segment elevation myocardial infarction of inferior wall involving right ventricle (disorder) |
| 24 | 233825009 | Acute Q wave infarction -  anteroseptal (disorder) | 76 | 703360004 | Subsequent non-ST segment elevation myocardial infarction (disorder) |
| 25 | 233826005 | Acute non-Q wave infarction  - anteroseptal (disorder) | 77 | 70422006 | Acute subendocardial infarction |
| 26 | 233827001 | Acute Q wave infarction -  anterolateral (disorder) | 78 | 70998009 | Acute myocardial infarction of basal inferior segment of left ventricle (disorder) |
| 27 | 233828006 | Acute non-Q wave infarction  - anterolateral (disorder) | 79 | 73795002 | Acute myocardial infarction of inferior wall |
| 28 | 233829003 | Acute Q wave infarction -  inferior (disorder) | 80 | 76593002 | Acute myocardial infarction of inferoposterior wall |
| 29 | 233830008 | Acute non-Q wave infarction  - inferior (disorder) | 81 | 79009004 | Acute myocardial infarction of septum |
| 30 | 233831007 | Acute Q wave infarction -  inferolateral (disorder) | 82 | 836293000 | Acute myocardial infarction of right ventricle (disorder) |
| 31 | 233832000 | Acute non-Q wave infarction  - inferolateral (disorder) | 83 | 836294006 | Acute myocardial infarction of apex of heart (disorder) |
| 32 | 233833005 | Acute Q wave infarction -  lateral (disorder) | 84 | 836295007 | Acute myocardial infarction of inferolateral wall with posterior extension (disorder) |
| 33 | 233834004 | Acute non-Q wave infarction  - lateral (disorder) | 85 | 840309000 | Acute ST segment elevation myocardial infarction due to occlusion of proximal portion of anterior descending branch of left coronary  artery (disorder) |

| 34 | 233835003 | Acute widespread myocardial infarction (disorder) | 86 | 840312002 | Acute ST segment elevation myocardial infarction due to occlusion of mid portion of anterior descending branch of left coronary artery (disorder) |
| --- | --- | --- | --- | --- | --- |
| 35 | 233838001 | Acute posterior myocardial infarction | 87 | 840316004 | Acute ST segment elevation myocardial infarction due to occlusion of distal portion of anterior descending branch of left coronary artery (disorder) |
| 36 | 233843008 | Silent myocardial infarction | 88 | 840609007 | Acute ST segment elevation myocardial infarction due to occlusion of anterior descending branch of left coronary artery (disorder) |
| 37 | 282006 | Acute myocardial infarction of basal-lateral wall (disorder) | 89 | 840680009 | Acute ST segment elevation myocardial infarction due to occlusion of septal branch of anterior descending branch of left coronary artery (disorder) |
| 38 | 285981000119103 | Acute ST segment elevation myocardial infarction involving left anterior descending coronary artery (disorder) | 90 | 846668006 | Acute ST segment elevation myocardial infarction due to occlusion of diagonal branch of anterior descending branch of left coronary  artery (disorder) |
| 39 | 285991000119100 | Acute ST segment elevation myocardial infarction involving left main coronary artery (disorder) | 91 | 846683001 | Acute ST segment elevation myocardial infarction due to occlusion of intermediate artery (disorder) |
| 40 | 304914007 | Acute Q wave myocardial  infarction | 92 | 868214006 | Acute ST segment elevation myocardial infarction due to occlusion of proximal portion of right coronary artery  (disorder) |
| 41 | 307140009 | Acute non-Q wave infarction | 93 | 868217004 | Acute ST segment elevation myocardial infarction due to occlusion of distal portion of right coronary artery (disorder) |
| 42 | 311792005 | Postoperative transmural myocardial infarction of anterior wall | 94 | 868220007 | Acute ST segment elevation myocardial infarction due to occlusion of mid portion of right coronary artery (disorder) |
| 43 | 311793000 | Postoperative transmural myocardial infarction of  inferior wall | 95 | 868224003 | Acute ST segment elevation myocardial infarction due to occlusion of marginal branch of right coronary artery (disorder) |
| 44 | 311796008 | Postoperative subendocardial myocardial  infarction | 96 | 868225002 | Acute ST segment elevation myocardial infarction due to occlusion of posterior descending branch of right coronary artery (disorder) |
| 45 | 314207007 | Non-Q wave myocardial  infarction (disorder) | 97 | 868226001 | Acute ST segment elevation myocardial infarction due to occlusion of posterior lateral branch of right coronary artery (disorder) |
| 46 | 315287002 | Diabetes mellitus insulinglucose infusion in acute myocardial infarction | 98 | 879955009 | Myocardial infarction with non-obstructive coronary  artery (disorder) |

| 47 | 371068009 | Myocardial infarction with complication (disorder) | 99 | 896689003 | Acute myocardial infarction due to occlusion of circumflex branch of left coronary artery (disorder) |
| --- | --- | --- | --- | --- | --- |
| 48 | 394710008 | First myocardial infarction  (disorder) | 100 | 896691006 | Acute ST segment elevation myocardial infarction due to occlusion of circumflex branch of left coronary artery (disorder) |
| 49 | 401303003 | Acute ST segment elevation myocardial infarction (disorder) | 101 | 896696001 | Acute ST segment elevation myocardial infarction of apex of heart (disorder) |
| 50 | 401314000 | Acute non-ST segment elevation myocardial infarction (disorder) | 102 | 896697005 | Acute ST segment elevation myocardial infarction of right ventricle (disorder) |
| 51 | 428196007 | Mixed myocardial ischemia and infarction (disorder) | 103 | I21 | Acute myocardial infarction |
| 52 | 44811000087108 | Acute ST segment elevation myocardial infarction due to occlusion of distal portion of circumflex branch of left coronary artery (disorder) | 104 | I22 | Subsequent myocardial  infarction |
| **Stable Angina** | | | | | |
| 1 | 3546002 | Aortocoronary artery bypass graft with saphenous vein  graft | 35 | 233821000 | New onset angina |
| 2 | 11101003 | Percutaneous transluminal coronary angioplasty | 36 | 265481001 | Double anastomosis of mammary arteries to coronary  arteries |
| 3 | 14201006 | Coronary angioplasty by open chest approach | 37 | 275215001 | LIMA single anastomosis |
| 4 | 20738002 | Endarterectomy of coronary artery | 38 | 275216000 | RIMA single anastomosis |
| 5 | 21470009 | Syncope anginosa | 39 | 275252001 | LIMA sequential anastomosis |
| 6 | 26141007 | ST segment depression | 40 | 275253006 | RIMA sequential anastomosis |
| 7 | 29843007 | Coronary atherectomy by laser | 41 | 276861004 | Percutaneous balloon angioplasty of artery |
| 8 | 31413008 | Operative procedure on  coronary arteries | 42 | 300995000 | Exercise-induced angina |
| 9 | 35928006 | Nocturnal angina | 43 | 308117007 | Antianginal therapy |
| 10 | 36969009 | Placement of stents in coronary artery | 44 | 312610006 | Prosthetic graft patch angioplasty |
| 11 | 59021001 | Angina decubitus | 45 | 312611005 | Percutaneous transluminal angioplasty of vascular graft |
| 12 | 59931005 | T wave inversion | 46 | 314116003 | Post infarct angina |
| 13 | 66812002 | T wave amplitude decreased | 47 | 371805005 | Significant coronary bypass graft disease (disorder) |
| 14 | 68466008 | Removal of coronary artery obstruction by percutaneous transluminal balloon, single vessel | 48 | 371808007 | Recurrent angina status post percutaneous transluminal coronary angioplasty (disorder) |
| 15 | 85053006 | Percutaneous transluminal coronary angioplasty,  multiple vessels | 49 | 371809004 | Recurrent angina status post coronary stent placement  (disorder) |
| 16 | 102594003 | Abnormal EKG finding | 50 | 371810009 | Recurrent angina status post coronary artery bypass graft (disorder) |
| 17 | 164861001 | ECG myocardial ischemia | 51 | 408546009 | Coronary artery bypass graft occlusion (disorder) |
| 18 | 164934002 | ECG: T wave abnormal | 52 | 412747004 | Radionuclide heart study abnormal (finding) |
| 19 | 175045009 | Connection of mammary  artery to coronary artery | 53 | 415071007 | Percutaneous endarterectomy (procedure) |

| 20 | 175047001 | Double implantation of mammary arteries into  coronary arteries | 54 | 426674009 | Percutaneous transluminal balloon angioplasty of artery (procedure) |
| --- | --- | --- | --- | --- | --- |
| 21 | 175050003 | Single implantation of mammary artery into coronary  artery | 55 | 429673002 | Arteriosclerosis in coronary artery bypass graft (disorder) |
| 22 | 175066001 | Percutaneous transluminal balloon angioplasty of bypass graft of coronary  artery | 56 | 429245005 | Recurrent coronary arteriosclerosis after percutaneous transluminal coronary angioplasty  (disorder) |
| 23 | 175764006 | Rotary blade angioplasty | 57 | 428781001 | Deep venous thrombosis associated with coronary artery bypass graft  (disorder) |
| 24 | 175882004 | Peroperative angioplasty | 58 | 429639007 | Percutaneous transluminal balloon angioplasty with insertion of stent into coronary artery (procedure) |
| 25 | 194828000 | Angina | 59 | 698378009 | Coronary artery bypass graft operation planned (situa-  tion) |
| 26 | 213037002 | Mechanical complication of coronary bypass | 60 | 707828002 | Percutaneous transluminal cutting balloon angioplasty of coronary artery (procedure) |
| 27 | 225566008 | Ischemic chest pain | 61 | 132111000119107 | Acute deep venous thrombosis of lower limb due to coronary artery bypass grafting |
| 28 | 232717009 | Coronary artery bypass graft | 62 | 132091000119104 | Chronic deep venous thrombosis of lower limb due to coronary artery bypass grafting |
| 29 | 232719007 | Coronary artery bypass graft  x 1 | 63 | 11018701000119100 | Coronary arteriosclerosis after percutaneous coronary angioplasty |
| 30 | 232720001 | CABG x 2 - Coronary artery bypass grafts x 2 | 64 | 139011000119104 | Coronary arteriosclerosis following coronary artery bypass graft |
| 31 | 232721002 | Coronary artery bypass grafts x 3 | 65 | 15960381000119100 | Angina co-occurrent and due to arteriosclerosis of coronary artery bypass graft  (disorder) |
| 32 | 232722009 | CABG x 4 - Coronary artery bypass grafts x 4 | 66 | I201 | Angina pectoris with documented spasm |
| 33 | 232726007 | Percutaneous endarterectomy of coronary  artery | 67 | I208 | Other forms of angina pectoris |
| 34 | 233819005 | Stable angina | 68 | I209 | Angina pectoris, unspecified |
| **Prior revascularisation** | | | | | |
| 1 | 15256002 | Transmyocardial revascularization by laser technique (procedure) | 6 | 275227003 | Myocardial revascularization  (procedure) |
| 2 | 25263003 | Grafting of heart for revascularization (procedure) | 7 | 287277008 | Indirect heart revascularization (procedure) |
| 3 | 81266008 | Heart revascularization (procedure) | 8 | 713617008 | Percutaneous transluminal revascularization of chronic total occlusion of coronary artery using fluoroscopic guidance (procedure) |
| 4 | 90205004 | Cardiac revascularization with bypass anastomosis  (procedure) | 9 | 924381000000104 | Percutaneous transluminal revascularisation of chronic total occlusion of coronary artery using fluoroscopic guidance (procedure) |

| 5 | 174911007 | Revascularization of wall of heart (procedure) |  |  |  |
| --- | --- | --- | --- | --- | --- |
| **Cerebrovascular disease** [4] | | | | | |
| 1 | 111297002 | Nonparalytic stroke (disorder) | 138 | 330421000119102 | Ataxic hemiparesis of right nondominant side due to and following lacunar infarction (disorder) |
| 2 | 1153543002 | Occlusion of anterior cerebral artery (disorder) | 139 | 330791000119108 | Cerebrovascular accident due to thrombus of left  carotid artery (disorder) |
| 3 | 1153544008 | Occlusion of right anterior cerebral artery (disorder) | 140 | 34781003 | Vertebral artery syndrome  (disorder) |
| 4 | 1153545009 | Occlusion of left anterior cerebral artery (disorder) | 141 | 371040005 | Thrombotic stroke (disorder) |
| 5 | 1153546005 | Occlusion of bilateral posterior cerebral arteries (disorder) | 142 | 371041009 | Embolic stroke (disorder) |
| 6 | 1153607003 | Occlusion of right posterior communicating artery (disorder) | 143 | 373606000 | Occlusive stroke (disorder) |
| 7 | 1153608008 | Occlusion of left posterior communicating artery (disorder) | 144 | 413102000 | Infarction of basal ganglia  (disorder) |
| 8 | 1153611009 | Embolism of left anterior cerebral artery (disorder) | 145 | 413758000 | Cardioembolic stroke (disorder) |
| 9 | 1153612002 | Embolism of right anterior cerebral artery (disorder) | 146 | 422504002 | Ischemic stroke (disorder) |
| 10 | 1153630009 | Embolism of left carotid  artery (disorder) | 147 | 426107000 | Acute lacunar infarction  (disorder) |
| 11 | 1153631008 | Embolism of right carotid artery (disorder) | 148 | 426814001 | Transient cerebral ischemia due to atrial fibrillation (disorder) |
| 12 | 1153632001 | Embolism of bilateral middle cerebral arteries (disorder) | 149 | 426983002 | Infarction of medulla oblongata (disorder) |
| 13 | 1153633006 | Embolism of bilateral posterior cerebral arteries (disorder) | 150 | 432504007 | Cerebral infarction (disorder) |
| 14 | 1153634000 | Embolism of bilateral anterior cerebral arteries (disorder) | 151 | 444172003 | Recurrent transient cerebral ischemic attack (disorder) |
| 15 | 1155688007 | Embolism of bilateral  carotid arteries (disorder) | 152 | 444657001 | Superior cerebellar artery syndrome (disorder) |
| 16 | 1155699009 | Embolism of bilateral vertebral arteries (disorder) | 153 | 49422009 | Cortical hemorrhage (disorder) |
| 17 | 1156016006 | Thrombosis of left posterior cerebral artery (disorder) | 154 | 52201006 | Internal capsule hemorrhage  (disorder) |
| 18 | 1156017002 | Thrombosis of right posterior cerebral artery (disorder) | 155 | 5571000124103 | Cerebrovascular accident with intracranial  hemorrhage (disorder) |
| 19 | 1156018007 | Thrombosis of left cerebellar artery (disorder) | 156 | 56267009 | Multi-infarct dementia (disorder) |
| 20 | 1156019004 | Thrombosis of right cerebellar artery (disorder) | 157 | 57981008 | Progressing stroke (disorder) |
| 21 | 1156027008 | Thrombus of dural sinus in pregnancy (disorder) | 158 | 64009001 | Basilar artery syndrome  (disorder) |
| 22 | 1156029006 | Thrombus of dural sinus in puerperium (disorder) | 159 | 71444005 | Cerebral arterial thrombosis  (disorder) |
| 23 | 116288000 | Paralytic stroke (disorder) | 160 | 723082006 | Silent cerebral infarct (disorder) |
| 24 | 1179360000 | Fetal epilepsy due to perinatal stroke (disorder) | 161 | 724424009 | Cerebral ischemic stroke due to small artery occlusion  (disorder) |
| 25 | 1197363004 | Pediatric arterial ischemic stroke (disorder) | 162 | 724429004 | Stroke co-occurrent with migraine (disorder) |

| 26 | 1204189004 | Asymptomatic occlusion of intracranial vertebral artery (disorder) | 163 | 732923001 | Hemorrhage of medulla  oblongata (disorder) |
| --- | --- | --- | --- | --- | --- |
| 27 | 1204202004 | Occlusion of cerebral artery due to infection (disorder) | 164 | 734383005 | Thrombosis of left middle cerebral artery (disorder) |
| 28 | 1208871009 | Transient ischemic attack cooccurrent with subarachnoid hemorrhage (disorder) | 165 | 734384004 | Thrombosis of right middle cerebral artery (disorder) |
| 29 | 1231168008 | Malignant middle cerebral artery syndrome (disorder) | 166 | 734961002 | Embolus of left posterior cerebral artery (disorder) |
| 30 | 125081000119106 | Cerebral infarction due to occlusion of precerebral artery (disorder) | 167 | 734963004 | Embolus of right posterior cerebral artery (disorder) |
| 31 | 1260271009 | Hemorrhagic cerebral infarction due to hypertension  (disorder) | 168 | 734964005 | Embolus of left middle cerebral artery (disorder) |
| 32 | 1263550001 | Infarction of brain due to migraine (disorder) | 169 | 734965006 | Embolus of right middle  cerebral artery (disorder) |
| 33 | 1269237007 | Cerebrovascular accident due to thrombosis of anterior cerebral artery (disorder) | 170 | 75038005 | Cerebellar hemorrhage (disorder) |
| 34 | 1269238002 | Thrombosis of anterior cerebral artery (disorder) | 171 | 751371000000107 | Personal history of transient ischaemic attack (situation) |
| 35 | 1269239005 | Thrombosis of left anterior cerebral artery (disorder) | 172 | 75543006 | Cerebral embolism (disorder) |
| 36 | 1269240007 | Thrombosis of right anterior cerebral artery (disorder) | 173 | 762629007 | Occlusion of right middle cerebral artery by embolus  (disorder) |
| 37 | 1269243009 | Thrombosis of bilateral  carotid arteries (disorder) | 174 | 762630002 | Occlusion of left middle cerebral artery by embolus (disorder) |
| 38 | 1269245002 | Thrombosis of bilateral posterior cerebral arteries (disorder) | 175 | 762651004 | Occlusion of right posterior cerebral artery by embolus  (disorder) |
| 39 | 1269246001 | Thrombosis of bilateral vertebral arteries (disorder) | 176 | 762652006 | Occlusion of left posterior cerebral artery by embolus  (disorder) |
| 40 | 1269248000 | Cerebrovascular accident due to occlusion of anterior choroidal artery (disorder) | 177 | 7713009 | Intrapontine hemorrhage  (disorder) |
| 41 | 140921000119102 | Ischemic stroke without coma (disorder) | 178 | 78569004 | Posterior inferior cerebellar artery syndrome (disorder) |
| 42 | 14309005 | Anterior choroidal artery syndrome (disorder) | 179 | 788455001 | Occlusion of bilateral pontine arteries (disorder) |
| 43 | 15258001 | Subclavian steal syndrome  (disorder) | 180 | 788880006 | Cerebral ischemic stroke due to dissection of artery (disorder) |
| 44 | 161511000 | History of transient ischemic attack (situation) | 181 | 788881005 | Cerebral ischemic stroke due to aortic arch embolism (disorder) |
| 45 | 16218291000119100 | Acute cerebral ischemia (disorder) | 182 | 788882003 | Cerebral ischemic stroke due to global hypoperfusion with watershed infarct (disorder) |
| 46 | 16371781000119100 | Cerebellar stroke (disorder) | 183 | 788883008 | Cerebral ischemic stroke due to hypercoagulable state  (disorder) |
| 47 | 195163003 | Intracerebral hemorrhage (& [cerebrovascular accident due to]) (disorder) | 184 | 788884002 | Cerebral ischemic stroke due to subarachnoid hemorrhage (disorder) |
| 48 | 195165005 | Basal ganglia hemorrhage  (disorder) | 185 | 870544005 | Occlusion of distal basilar artery (disorder) |
| 49 | 195167002 | External capsule  hemorrhage (disorder) | 186 | 870579007 | Occlusion of branch of basilar artery (disorder) |
| 50 | 195168007 | Intracerebral hemorrhage with intraventricular  hemorrhage (disorder) | 187 | 87555007 | Claude’s syndrome (disorder) |

| 51 | 195169004 | Intracerebral hemorrhage multiple localized (disorder) | 188 | 90099008 | Subcortical leukoen-  cephalopathy (disorder) |
| --- | --- | --- | --- | --- | --- |
| 52 | 195185009 | Cerebral infarct due to thrombosis of precerebral arteries (disorder) | 189 | 95454007 | Brain stem hemorrhage (disorder) |
| 53 | 195186005 | Cerebral infarction due to embolism of precerebral  arteries (disorder) | 190 | 95457000 | Brain stem infarction (disorder) |
| 54 | 195189003 | Cerebral infarction due to thrombosis of cerebral arteries (disorder) | 191 | 95460007 | Cerebellar infarction (disorder) |
| 55 | 195190007 | Cerebral infarction due to embolism of cerebral arteries (disorder) | 192 | 99451000119105 | Cerebral infarction due to stenosis of carotid artery  (disorder) |
| 56 | 195199008 | Vertebrobasilar artery syndrome (disorder) | 193 | G45 | Transient cerebral ischaemic attacks and related syndromes |
| 57 | 195200006 | Carotid artery syndrome  hemispheric (disorder) | 194 | G450 | Vertebro-basilar artery syndrome |
| 58 | 195201005 | Multiple and bilateral precerebral artery syndromes  (disorder) | 195 | G451 | Carotid artery syndrome  (hemispheric) |
| 59 | 195205001 | Impending cerebral ischemia  (disorder) | 196 | G452 | Multiple and bilateral precerebral artery syndromes |
| 60 | 195206000 | Intermittent cerebral  ischemia (disorder) | 197 | G453 | Amaurosis fugax |
| 61 | 195209007 | Middle cerebral artery syndrome (disorder) | 198 | G454 | Transient global amnesia |
| 62 | 195210002 | Anterior cerebral artery syndrome (disorder) | 199 | G458 | Other transient cerebral ischaemic attacks and  related syndromes |
| 63 | 195211003 | Posterior cerebral artery syndrome (disorder) | 200 | G459 | Transient cerebral ischaemic attack, unspecified |
| 64 | 195212005 | Brainstem stroke syndrome  (disorder) | 201 | I60 | Subarachnoid haemorrhage |
| 65 | 195213000 | Cerebellar stroke syndrome  (disorder) | 202 | I600 | Subarachnoid haemorrhage from carotid siphon and  bifurcation |
| 66 | 195216008 | Left sided cerebral hemisphere cerebrovascular accident (disorder) | 203 | I601 | Subarachnoid haemorrhage from middle cerebral artery |
| 67 | 195217004 | Right sided cerebral hemisphere cerebrovascular accident (disorder) | 204 | I602 | Subarachnoid haemorrhage from anterior communicating artery |
| 68 | 195230003 | Cerebral infarction due to cerebral venous thrombosis non-pyogenic (disorder) | 205 | I603 | Subarachnoid haemorrhage from posterior communicating artery |
| 69 | 20059004 | Occlusion of cerebral artery  (disorder) | 206 | I604 | Subarachnoid haemorrhage from basilar artery |
| 70 | 20908003 | Subcortical cerebral hemorrhage (disorder) | 207 | I605 | Subarachnoid haemorrhage from vertebral artery |
| 71 | 230518009 | Infarction of optic tract (disorder) | 208 | I606 | Subarachnoid haemorrhage from other intracranial  arteries |
| 72 | 230523009 | Infarction of optic radiation  (disorder) | 209 | I607 | Subarachnoid haemorrhage from intracranial artery, unspecified |
| 73 | 230690007 | Cerebrovascular accident  (disorder) | 210 | I608 | Other subarachnoid haemorrhage |
| 74 | 230691006 | Cerebrovascular accident due to occlusion of cerebral artery (disorder) | 211 | I609 | Subarachnoid haemorrhage, unspecified |
| 75 | 230692004 | Infarction - precerebral (disorder) | 212 | I61 | Intracerebral haemorrhage |
| 76 | 230693009 | Anterior cerebral circulation infarction (disorder) | 213 | I610 | Intracerebral haemorrhage in hemisphere, subcortical |

| 77 | 230694003 | Total anterior cerebral circulation infarction (disorder) | 214 | I611 | Intracerebral haemorrhage in hemisphere, cortical |
| --- | --- | --- | --- | --- | --- |
| 78 | 230695002 | Partial anterior cerebral circulation infarction (disorder) | 215 | I612 | Intracerebral haemorrhage in hemisphere, unspecified |
| 79 | 230696001 | Posterior cerebral circulation infarction (disorder) | 216 | I613 | Intracerebral haemorrhage in brain stem |
| 80 | 230698000 | Lacunar infarction (disorder) | 217 | I614 | Intracerebral haemorrhage in cerebellum |
| 81 | 230699008 | Pure motor hemiparesis due to and following lacunar  infarction (disorder) | 218 | I615 | Intracerebral haemorrhage, intraventricular |
| 82 | 230700009 | Pure sensory stroke due to and following lacunar infarction (disorder) | 219 | I616 | Intracerebral haemorrhage, multiple localized |
| 83 | 230701008 | Pure sensorimotor motor hemiparesis due to and following lacunar infarction  (disorder) | 220 | I618 | Other intracerebral haemorrhage |
| 84 | 230702001 | Ataxic hemiparesis due to and following lacunar infarction (disorder) | 221 | I619 | Intracerebral haemorrhage, unspecified |
| 85 | 230703006 | Dysarthria-clumsy hand syndrome due to and following lacunar infarction  (disorder) | 222 | I62 | Other nontraumatic  intracranial haemorrhage |
| 86 | 230704000 | Multi-infarct state (disorder) | 223 | I620 | Subdural haemorrhage  (acute)(nontraumatic) |
| 87 | 230706003 | Hemorrhagic cerebral infarction (disorder) | 224 | I621 | Nontraumatic extradural haemorrhage |
| 88 | 230707007 | Anterior cerebral circulation hemorrhagic infarction (disorder) | 225 | I629 | Intracranial haemorrhage  (nontraumatic), unspecified |
| 89 | 230708002 | Posterior cerebral circulation hemorrhagic infarction (disorder) | 226 | I63 | Cerebral infarction |
| 90 | 230709005 | Massive supratentorial cerebral hemorrhage (disorder) | 227 | I630 | Cerebral infarction due to thrombosis of precerebral  arteries |
| 91 | 230710000 | Lobar cerebral hemorrhage  (disorder) | 228 | I631 | Cerebral infarction due to embolism of precerebral  arteries |
| 92 | 230711001 | Thalamic hemorrhage (disorder) | 229 | I632 | Cerebral infarction due to unspecified occlusion or stenosis of precerebral  arteries |
| 93 | 230712008 | Lacunar hemorrhage (disorder) | 230 | I633 | Cerebral infarction due to thrombosis of cerebral arteries |
| 94 | 230713003 | Stroke of uncertain pathology (disorder) | 231 | I634 | Cerebral infarction due to embolism of cerebral arteries |
| 95 | 230714009 | Anterior circulation stroke of uncertain pathology (disorder) | 232 | I635 | Cerebral infarction due to unspecified occlusion or stenosis of cerebral arteries |
| 96 | 230715005 | Posterior circulation stroke of uncertain pathology (disorder) | 233 | I636 | Cerebral infarction due to cerebral venous thrombosis, nonpyogenic |
| 97 | 230716006 | Carotid territory transient ischemic attack (disorder) | 234 | I638 | Other cerebral infarction |
| 98 | 230717002 | Vertebrobasilar territory transient ischemic attack  (disorder) | 235 | I639 | Cerebral infarction, unspecified |
| 99 | 24654003 | Weber-Gubler syndrome  (disorder) | 236 | I64 | Stroke, not specified as haemorrhage or infarction |

| 100 | 25133001 | Completed stroke (disorder) | 237 | I65 | Occlusion and stenosis of precerebral arteries, not resulting in cerebral  infarction |
| --- | --- | --- | --- | --- | --- |
| 101 | 266257000 | Transient ischemic attack  (disorder) | 238 | I650 | Occlusion and stenosis of vertebral artery |
| 102 | 274100004 | Cerebral hemorrhage (disorder) | 239 | I651 | Occlusion and stenosis of basilar artery |
| 103 | 275434003 | Stroke in the puerperium  (disorder) | 240 | I652 | Occlusion and stenosis of carotid artery |
| 104 | 276219001 | Occipital cerebral infarction  (disorder) | 241 | I653 | Occlusion and stenosis of multiple and bilateral precerebral arteries |
| 105 | 276220007 | Foville syndrome (disorder) | 242 | I658 | Occlusion and stenosis of other precerebral artery |
| 106 | 276221006 | Millard-Gubler syndrome  (disorder) | 243 | I659 | Occlusion and stenosis of unspecified precerebral  artery |
| 107 | 276222004 | Top of basilar syndrome (disorder) | 244 | I66 | Occlusion and stenosis of cerebral arteries, not resulting in cerebral infarction |
| 108 | 276722003 | Intracerebellar and posterior fossa hemorrhage (disorder) | 245 | I660 | Occlusion and stenosis of middle cerebral artery |
| 109 | 281240008 | Extension of cerebrovascular accident (disorder) | 246 | I661 | Occlusion and stenosis of anterior cerebral artery |
| 110 | 290641000119107 | Dysphagia due to and following non-traumatic intracerebral hemorrhage (disorder) | 247 | I662 | Occlusion and stenosis of posterior cerebral artery |
| 111 | 291511000119103 | Spontaneous hemorrhage of deep cerebral hemisphere  (disorder) | 248 | I663 | Occlusion and stenosis of  cerebellar arteries |
| 112 | 291521000119105 | Spontaneous hemorrhage of cortical intracerebral hemisphere (disorder) | 249 | I664 | Occlusion and stenosis of multiple and bilateral cerebral arteries |
| 113 | 291531000119108 | Spontaneous hemorrhage of cerebral hemisphere (disorder) | 250 | I668 | Occlusion and stenosis of other cerebral artery |
| 114 | 291541000119104 | Spontaneous hemorrhage of brain stem (disorder) | 251 | I669 | Occlusion and stenosis of unspecified cerebral artery |
| 115 | 292851000119109 | Ataxic hemiparesis of right dominant side due to and following lacunar infarction (disorder) | 252 | I67 | Other cerebrovascular diseases |
| 116 | 292861000119106 | Ataxic hemiparesis of left dominant side due to and following lacunar infarction (disorder) | 253 | I670 | Dissection of cerebral arteries, nonruptured |
| 117 | 297138001 | Embolus of circle of Willis  (disorder) | 254 | I671 | Cerebral aneurysm, nonruptured |
| 118 | 302904002 | Infarction of visual cortex  (disorder) | 255 | I672 | Cerebral atherosclerosis |
| 119 | 307363008 | Multiple lacunar infarcts  (disorder) | 256 | I673 | Progressive vascular  leukoencephalopathy |
| 120 | 307766002 | Left sided cerebral infarction  (disorder) | 257 | I674 | Hypertensive encephalopathy |
| 121 | 307767006 | Right sided cerebral infarction (disorder) | 258 | I675 | Moyamoya disease |
| 122 | 308128006 | Right sided intracerebral hemorrhage unspecified  (disorder) | 259 | I676 | Nonpyogenic thrombosis of intracranial venous system |
| 123 | 329361000119107 | Cerebrovascular accident due to occlusion of right middle cerebral artery by  embolus (disorder) | 260 | I677 | Cerebral arteritis, not elsewhere classified |

|  | | 124 | 329371000119101 | | Cerebrovascular accident due to occlusion of left middle cerebral artery by  embolus (disorder) | | | 261 | I678 | | Other specified cerebrovascular diseases |
| --- | --- | --- | --- | --- | --- | --- | --- | --- | --- | --- | --- |
|  | | 125 | 329421000119107 | | Cerebrovascular accident due to occlusion of right posterior cerebral artery by embolus (disorder) | | | 262 | I679 | | Cerebrovascular disease, unspecified |
|  | | 126 | 329431000119105 | | Cerebrovascular accident due to occlusion of left posterior cerebral artery by  embolus (disorder) | | | 263 | I68 | | Cerebrovascular disorders in diseases classified elsewhere |
|  | | 127 | 329451000119104 | | Cerebrovascular accident due to occlusion of right cerebellar artery by embolus (disorder) | | | 264 | I680 | | Cerebral amyloid angiopathy |
|  | | 128 | 329461000119102 | | Cerebrovascular accident due to occlusion of left cerebellar artery by embolus  (disorder) | | | 265 | I681 | | Cerebral arteritis in infectious and parasitic diseases classified elsewhere |
|  | | 129 | 329481000119106 | | Occlusion of right middle cerebral artery (disorder) | | | 266 | I682 | | Cerebral arteritis in other diseases classified elsewhere |
|  | | 130 | 329491000119109 | | Occlusion of left middle cerebral artery (disorder) | | | 267 | I688 | | Other cerebrovascular disorders in diseases classified elsewhere |
|  | | 131 | 329501000119102 | | Occlusion of bilateral middle cerebral arteries (disorder) | | | 268 | I69 | | Sequelae of cerebrovascular disease |
|  | | 132 | 329541000119100 | | Occlusion of bilateral anterior cerebral arteries (disorder) | | | 269 | I690 | | Sequelae of subarachnoid haemorrhage |
|  | | 133 | 329561000119101 | | Occlusion of right posterior cerebral artery (disorder) | | | 270 | I691 | | Sequelae of intracerebral haemorrhage |
|  | | 134 | 329571000119107 | | Occlusion of left posterior cerebral artery (disorder) | | | 271 | I692 | | Sequelae of other nontraumatic intracranial haemorrhage |
|  | | 135 | 329641000119104 | | Cerebrovascular accident due to thrombus of basilar artery (disorder) | | | 272 | I693 | | Sequelae of cerebral infarction |
|  | | 136 | 329651000119102 | | Cerebrovascular accident due to thrombus of right  carotid artery (disorder) | | | 273 | I694 | | Sequelae of stroke, not specified as haemorrhage or  infarction |
|  | | 137 | 330411000119109 | | Ataxic hemiparesis of left nondominant side due to and following lacunar infarction (disorder) | | | 274 | I698 | | Sequelae of other and unspecified cerebrovascular diseases |
|  | | **Peripheral arterial disease** [3] | | | | | | | | | |
|  | | 1 | 63491006 | | Intermittent claudication | | | 14 | 400047006 | | Peripheral vascular disease |
|  | | 2 | 127014009 | | Peripheral angiopathy due to diabetes mellitus | | | 15 | 713412006 | | Ischemic foot with rest pain |
|  | | 3 | 233958001 | | Peripheral ischemia | | | 16 | 723870003 | | Acute occlusion of artery of lower limb co-occurrent and due to thromboembolus |
|  | | 4 | 233961000 | | Lower limb ischemia | | | 17 | 792844003 | | Limb pain at rest due to atherosclerosis of artery of lower limb |
|  | | 5 | 233962007 | | Critical lower limb ischemia | | | 18 | 792846001 | | Ulcer of heel due to atherosclerosis of artery of lower limb |
|  | | 6 | 275520000 | | Claudication | | | 19 | 792850008 | | Ulcer of ankle due to atherosclerosis of artery of lower limb |
|  | | 7 | 300917007 | | Ischemia of feet | | | 20 | 792855003 | | Ulcer of calf due to atherosclerosis of artery of lower limb |
|  | | 8 | 301755001 | | Ischemic foot | | | 21 | 840580004 | | Peripheral arterial disease |
| 9 | 307406004 | | | Trash foot | | 22 | 31211000119101 | | | Peripheral angiopathy due to type 1 diabetes mellitus | |
| 10 | 307408003 | | | Ischemic toe | | 23 | 34881000119105 | | | Peripheral vascular disease associated with another disorder | |
| 11 | 312822006 | | | Critical ischemia of foot | | 24 | 836711000000108 | | | Ischaemic lower limb pain at rest | |
| 12 | 314902007 | | | Peripheral angiopathy due to type 2 diabetes mellitus | | 25 | 10665871000119100 | | | Ischemic ulcer of left ankle due to atherosclerotic disease | |
| 13 | 399957001 | | | Peripheral arterial occlusive disease | |  |  | | |  | |

**Table S9**: Smoking SNOMED code list

|  |  | | | **Former smoker** | | | | | | | |
| --- | --- | --- | --- | --- | --- | --- | --- | --- | --- | --- | --- |
| # | Code | | | Term | | # | Code | | | Term | |
| 1 | 160617001 | | | Stopped smoking (finding) | | 14 | 360900008 | | | Aggressive ex-smoker (finding) | |
| 2 | 160620009 | | | Ex-pipe smoker (finding) | | 15 | 53896009 | | | Tolerant ex-smoker (finding) | |
| 3 | 160621008 | | | Ex-cigar smoker (finding) | | 16 | 735112005 | | | Date ceased using moist  tobacco (observable entity) | |
| 4 | 160625004 | | | Date ceased smoking  (observable entity) | | 17 | 735128000 | | | Ex-smoker for less than 1  year (finding) | |
| 5 | 228486009 | | | Time since stopped smoking  (observable entity) | | 18 | 8517006 | | | Ex-smoker (finding) | |
| 6 | 266921000 | | | Ex-trivial cigarette smoker  (*<*1/day) (finding) | | 19 | 1092031000000100 | | | Ex-smoker amount unknown  (finding) | |
| 7 | 266922007 | | | Ex-light cigarette smoker (1-  9/day) (finding) | | 20 | 1092041000000100 | | | Ex-very heavy smoker  (40+/day) (finding) | |
| 8 | 266923002 | | | Ex-moderate cigarette smoker (10-19/day)  (finding) | | 21 | 1092071000000100 | | | Ex-heavy smoker  (20-39/day) (finding) | |
| 9 | 266924008 | | | Ex-heavy cigarette smoker  (20-39/day) (finding) | | 22 | 1092091000000100 | | | Ex-moderate smoker (10-  19/day) (finding) | |
| 10 | 266925009 | | | Ex-very heavy cigarette smoker (40+/day) (finding) | | 23 | 1092111000000100 | | | Ex-light smoker (1-9/day)  (finding) | |
| 11 | 266928006 | | | Ex-cigarette smoker amount unknown (finding) | | 24 | 1092131000000100 | | | Ex-trivial smoker (¡1/day)  (finding) | |
| 12 | 281018007 | | | Ex-cigarette smoker (finding) | | 25 | 48031000119106 | | | Ex-smoker for more than 1  year (finding) | |
| 13 | 360890004 | | | Intolerant ex-smoker (finding) | | 26 | 492191000000103 | | | Ex roll-up cigarette smoker  (finding) | |
|  |  | | | **Current smoker** | | | | | | | |
| 1 | 134406006 | | | Smoking reduced (finding) | | 21 | 266920004 | | | Trivial cigarette smoker (less than one cigarette/day)  (finding) | |
| 2 | 160603005 | | | Light cigarette smoker (1-9 cigs/day) (finding) | | 22 | 266929003 | | | Smoking started (finding) | |
| 3 | 160604004 | | | Moderate cigarette smoker  (10-19 cigs/day) (finding) | | 23 | 308438006 | | | Smoking restarted (finding) | |
| 4 | 160605003 | | | Heavy cigarette smoker (20-  39 cigs/day) (finding) | | 24 | 394871007 | | | Thinking about stopping smoking (finding) | |
| 5 | 160606002 | | | Very heavy cigarette smoker  (40+ cigs/day) (finding) | | 25 | 394872000 | | | Ready to stop smoking (finding) | |
| 6 | 160612007 | | | Keeps trying to stop smoking (finding) | | 26 | 394873005 | | | Not interested in stopping smoking (finding) | |
| 7 | 160613002 | | | Admitted tobacco consumption possibly untrue (finding) | | 27 | 401159003 | | | Reason for restarting smoking (observable entity) | |
| 8 | 160616005 | | | Trying to give up smoking  (finding) | | 28 | 401201003 | | | Cigarette pack-years  (observable entity) | |
|  | | 9 | 160619003 | | Rolls own cigarettes (finding) | | | 29 | 413173009 | | Minutes from waking to first tobacco consumption (observable entity) |
|  | | 10 | 225934006 | | Smokes in bed (finding) | | | 30 | 446172000 | | Failed attempt to stop smoking (finding) |
|  | | 11 | 230056004 | | Cigarette consumption  (observable entity) | | | 31 | 449868002 | | Smokes tobacco daily (finding) |
|  | | 12 | 230057008 | | Cigar consumption (observable entity) | | | 32 | 56578002 | | Moderate smoker (20 or less per day) (finding) |
|  | | 13 | 230058003 | | Pipe tobacco consumption  (observable entity) | | | 33 | 56771006 | | Heavy smoker (over 20 per  day) (finding) |
|  | | 14 | 230059006 | | Occasional cigarette smoker  (finding) | | | 34 | 59978006 | | Cigar smoker (finding) |
|  | | 15 | 230060001 | | Light cigarette smoker (finding) | | | 35 | 65568007 | | Cigarette smoker (finding) |
|  | | 16 | 230062009 | | Moderate cigarette smoker  (finding) | | | 36 | 77176002 | | Smoker (finding) |
|  | | 17 | 230063004 | | Heavy cigarette smoker  (finding) | | | 37 | 82302008 | | Pipe smoker (finding) |
|  | | 18 | 230064005 | | Very heavy cigarette smoker  (finding) | | | 38 | 836001000000109 | | Water pipe tobacco consumption (observable entity) |
|  | | 19 | 230065006 | | Chain smoker (finding) | | | 39 | 428041000124106 | | Occasional tobacco smoker  (finding) |
|  | | 20 | 266918002 | | Tobacco smoking consumption (observable entity) | | | 40 | 203191000000107 | | Wants to stop smoking (finding) |
|  | |  |  | | **Non-smoker** | | | | | |  |
|  | | 1 | 266919005 | | Never smoked tobacco (finding) | | | 2 | 221000119102 | | Never smoked any substance  (finding) |

**Table S10**: BMI SNOMED code list

|  | | # | Code | | Term | | | # | Code | | Term |
| --- | --- | --- | --- | --- | --- | --- | --- | --- | --- | --- | --- |
|  | | 1 | 788996008 | | Obesity in adolescence (disorder) | | | 16 | 427090001 | | Body mass index less than  16.5 (finding) |
|  | | 2 | 819948005 | | Obese class III (finding) | | | 17 | 301331008 | | Finding of body mass index  (finding) |
|  | | 3 | 35425004 | | Normal body mass index  (finding) | | | 18 | 846931000000101 | | Baseline body mass index  (observable entity) |
|  | | 4 | 412768003 | | Body mass index 20-24 - normal (finding) | | | 19 | 408512008 | | Body mass index 40+ -  severely obese (finding) |
|  | | 5 | 162864005 | | Body mass index 30+ - obe-  sity (finding) | | | 20 | 60621009 | | Body mass index (observable entity) |
|  | | 6 | 248342006 | | Underweight (finding) | | | 21 | 162764001 | | On examination - weight greater than 20% below ideal (finding) |
|  | | 7 | 268915006 | | On examination - weight 10-  20% over ideal (finding) | | | 22 | 162765000 | | On examination - weight 10-  20% below ideal (finding) |
|  | | 8 | 268916007 | | On examination - weight greater than 20% over ideal  (finding) | | | 23 | 162769006 | | On examination - Under-  weight (finding) |
|  | | 9 | 275947003 | | On examination - overweight  (finding) | | | 24 | 238131007 | | Overweight (finding) |
|  | | 10 | 190966007 | | Extreme obesity with alveolar hypoventilation (disorder) | | | 25 | 443371000124107 | | Obese class I (finding) |
|  | | 11 | 162690006 | | On examination - obese  (finding) | | | 26 | 443381000124105 | | Obese class II (finding) |
|  | | 12 | 414915002 | | Obese (finding) | | | 27 | 914721000000105 | | Obese class I (body mass  index 30.0 - 34.9) (finding) |
|  | | 13 | 162863004 | | Body mass index 25-29 -  overweight (finding) | | | 28 | 914731000000107 | | Obese class II (body mass index 35.0 - 39.9) (finding) |
| 14 | 722595002 | | | Overweight in adulthood with body mass index of 25 or more but less than 30  (finding) | | 29 | 914741000000103 | | | Obese class III (body mass index equal to or greater than 40.0) (finding) | |
| 15 | 310252000 | | | Body mass index less than 20  (finding) | | 30 |  | | |  | |

**Table S11**: Angina or heart attack in a 1*^st^* degree relative, SNOMED and ICD-

10 code list

| # | Code | Term |  | # | Code | Term |
| --- | --- | --- | --- | --- | --- | --- |
| 1 | 401122004 | Angina in 1*^st^*  *<* 55 | degree male | 6 | 315627005 | Angina in 1*^st^* degree female age unknown |
| 2 | 401065001 | Angina in female *<* 65 | 1*^st^* degree | 7 | 275121006 | F/H angina |
| 3 | 315623009 | Angina in 1*^st^* age known | degree male | 8 | 275932007 | F/H angina less than 60 |
| 4 | 315625002 | Angina in 1*^st^* age unknown | degree make | 9 | 161502000 | H/O: myocardial infarct at less than 60 |
| 5 | 315626001 | Angina in female age kn | 1*^st^* degree own | 10 | 308065005 | H/O: Myocardial infarction in last year |

**Table S12**: Chronic Kidney Disease (CKD) SNOMED and ICD-10 code list [11]

|  | **CKD Stage 3** | | | | |
| --- | --- | --- | --- | --- | --- |
| 1 | 700379002 | chronic kidney disease stage 3b | 13 | 324371000000106 | chronic kidney disease stage 3b with proteinuria |
| 2 | 324251000000105 | chronic kidney disease stage 3 with proteinuria | 14 | 950081000000107 | ckd g3ba2 - chronic kidney disease with glomerular filtration rate category g3b and albuminuria category a2 |
| 3 | 324311000000101 | chronic kidney disease stage 3a with proteinuria | 15 | 96731000119100 | Hypertensive heart AND  Chronic kidney disease stage  3 (disorder) |
| 4 | 994421000006107 | chronic kidney disease stage 3 | 16 | 284991000119104 | Chronic kidney disease stage 3 due to benign hypertension  (disorder) |
| 5 | 700378005 | chronic kidney disease stage 3a | 17 | 368441000119102 | Chronic kidney disease stage 3 due to drug induced dia-  betes mellitus (disorder) |
| 6 | 949901000000109 | ckd g3aa2 - chronic kidney disease with glomerular filtration rate category g3a and albuminuria category a2 | 18 | 129171000119106 | Chronic kidney disease stage 3 due to hypertension (disorder) |
| 7 | 949921000000100 | ckd g3aa3 - chronic kidney disease with glomerular filtration rate category g3a and albuminuria category a3 | 19 | 90741000119107 | Chronic kidney disease stage 3 due to type 1 diabetes mellitus (disorder) |
| 8 | 950061000000103 | ckd g3ba1 - chronic kidney disease with glomerular filtration rate category g3b and albuminuria category a1 | 20 | 731000119105 | Chronic kidney disease stage 3 due to type 2 diabetes mellitus (disorder) |
| 9 | 950101000000101 | ckd g3ba3 - chronic kidney disease with glomerular filtration rate category g3b and albuminuria category a3 | 21 | 949881000000106 | chronic kidney disease with glomerular filtration rate category g3a and albuminuria category a1 |

| 10 | 433144002 | chronic kidney disease stage 3 | 22 | 324341000000100 | ckd (chronic kidney disease) stage 3a without proteinuria |
| --- | --- | --- | --- | --- | --- |
| 11 | 324411000000105 | chronic kidney disease stage 3b without proteinuria | 23 | 285871000119106 | Malignant Hypertensive Chronic kidney disease stage  3 (disorder) |
| 12 | 324281000000104 | ckd (chronic kidney disease) stage 3 without proteinuria | 24 | N18.3 | ICD-10 chronic kidney disease stage 3 |
| **CKD Stage 4** | | | | | |
| 1 | 994431000006105 | chronic kidney disease stage 4 | 8 | 90751000119109 | Chronic kidney disease stage 4 due to type 1 diabetes mellitus (disorder) |
| 2 | 950231000000104 | ckd g4a3 - chronic kidney disease with glomerular filtration rate category g4 and albuminuria category a3 | 9 | 721000119107 | Chronic kidney disease stage 4 due to type 2 diabetes mellitus (disorder) |
| 3 | 431857002 | chronic kidney disease stage 4 | 10 | 950211000000107 | chronic kidney disease with glomerular filtration rate category g4 and albuminuria category a2 |
| 4 | 96721000119103 | Hypertensive heart AND  Chronic kidney disease stage  4 (disorder) | 11 | 324441000000106 | ckd (chronic kidney disease) stage 4 with proteinuria |
| 5 | 324471000000100 | chronic kidney disease stage 4 without proteinuria | 12 | 950181000000106 | ckd g4a1 - chronic kidney disease with glomerular filtration rate category g4 and albuminuria category a1 |
| 6 | 285001000119105 | Chronic kidney disease stage 4 due to benign hypertension  (disorder) | 13 | 285881000119109 | Malignant Hypertensive Chronic kidney disease stage  4 (disorder) |
| 7 | 368451000119100 | Chronic kidney disease stage 4 due to drug induced dia-  betes mellitus (disorder) | 14 | N18.4 | ICD-10 chronic kidney disease stage 4 |
| **CKD Stage 5** | | | | | |
| 1 | 423062001 | Stenosis of arteriovenous  dialysis fistula | 38 | 153851000119106 | Malignant Hypertensive Chronic kidney disease stage  5 (disorder) |
| 2 | 438546008 | Ligation of arteriovenous  dialysis fistula | 39 | 285841000119104 | Malignant Hypertensive end stage renal disease (disorder) |
| 3 | 180272001 | Placement ambulatory dialysis apparatus - compens  renal fail | 40 | 286371000119107 | Malignant Hypertensive end stage renal disease on dialysis (disorder) |
| 4 | 385971003 | [V]Preparatory care for dialysis | 41 | 991521000000102 | Dialysis fluid glucose level |
| 5 | 302497006 | Haemodialysis | 42 | 426361000000104 | [V]Unspecified aftercare involving intermittent  dialysis |
| 6 | 251859005 | Dialysis finding | 43 | 96701000119107 | Hypertensive heart AND Chronic kidney disease on  dialysis (disorder) |
| 7 | 108241001 | Dialysis procedure | 44 | 285011000119108 | Chronic kidney disease stage 5 due to benign hypertension  (disorder) |
| 8 | 161693006 | H/O: renal dialysis | 45 | 368461000119103 | Chronic kidney disease stage 5 due to drug induced dia-  betes mellitus (disorder) |
| 9 | 17778006 | Mechanical complication of dialysis catheter | 46 | 410511000000103 | [V]Aftercare involving renal dialysis NOS |
| 10 | 216878005 | Accidental cut, puncture, perforation or haemorrhage during kidney dialysis | 47 | 426351000000102 | [V]Aftercare involving peritoneal dialysis |
| 11 | 276883000 | Peritoneal dialysis-  associated peritonitis | 48 | 5571000001109 | solution haemodialysis stage 5 |
| 12 | 265764009 | Renal dialysis | 49 | 129161000119100 | Chronic kidney disease stage 5 due to hypertension (disorder) |

| 13 | 180277007 | Insertion of temporary peritoneal dialysis catheter | 50 | 90761000119106 | Chronic kidney disease stage 5 due to type 1 diabetes mellitus (disorder) |
| --- | --- | --- | --- | --- | --- |
| 14 | 79827002 | cadaveric renal transplant | 51 | 711000119100 | Chronic kidney disease stage 5 due to type 2 diabetes mellitus (disorder) |
| 15 | 216933008 | Failure of sterile precautions during kidney dialysis | 52 | 950251000000106 | ckd g5a1 - chronic kidney disease with glomerular filtration rate category g5 and albuminuria category a1 |
| 16 | 238318009 | Continuous ambulatory peritoneal dialysis | 53 | 96711000119105 | Hypertensive heart AND  Chronic kidney disease stage  5 (disorder) |
| 17 | 269698004 | failure of sterile precautions during kidney dialysis | 54 | 324501000000107 | ckd (chronic kidney disease) stage 5 with proteinuria |
| 18 | 269691005 | very mild acute rejection of renal transplant | 55 | 950311000000102 | ckd g5a3 - chronic kidney disease with glomerular filtration rate category g5 and albuminuria category a3 |
| 19 | 216904007 | acute rejection of renal transplant - grade i | 56 | 324541000000105 | chronic kidney disease stage 5 without proteinuria |
| 20 | 70536003 | Renal transplant stage 5 | 57 | 994441000006100 | chronic kidney disease stage 5 |
| 21 | 427053002 | Extracorporeal albumin haemodialysis stage 5 | 58 | 950291000000103 | chronic kidney disease with glomerular filtration rate category g5 and albuminuria category a2 |
| 22 | 225283000 | Priming haemodialysis lines stage 5 | 59 | N18.5 | ICD-10 chronic kidney disease stage 5 |
| 23 | 420106004 | Renal transplant venogram stage 5 | 60 | Z49.1 | ICD-10 extracorporeal dialysis |
| 24 | 708932005 | Emergency haemodialysis stage 5 | 61 | Z49.2 | ICD-10 other dialysis |
| 25 | 714153000 | chronic kidney disease 5t | 62 | Z94.0 | ICD-10 kidney transplant  status |
| 26 | 714152005 | ckd (chronic kidney disease) stage 5d | 63 | Z99.2 | ICD-10 dependence on renal dialysis//on dialysis treatment |
| 27 | 433146000 | chronic kidney disease stage 5 | 64 | N18.6 | ICD-10 patients with CKD  requiring dialysis |
| 28 | 180273006 | Removal of ambulatory peritoneal dialysis catheter | 65 | I77.0 | ICD-10 arteriovenous fistula |
| 29 | 426340003 | Creation of graft fistula for dialysis | 66 | N16.5 | ICD-10 renal tubulointerstitial disorders in  transplant rejection |
| 30 | 71192002 | Peritoneal dialysis NEC | 67 | T82.4 | ICD-10 mechanical complication of vascular dialysis catheter |
| 31 | 116223007 | Kidney dialysis with complication, without blame | 68 | T86.1 | ICD-10 kidney transplant failure and rejection |
| 32 | 428648006 | Automated peritoneal dialysis | 69 | Y60.2 | ICD-10 during kidney dialysis or other perfusion |
| 33 | 85223007 | Complication of haemodialysis stage 5 | 70 | Y61.2 | ICD-10 during kidney dialysis or other perfusion |
| 34 | 57274006 | Initial haemodialysis stage 5 | 71 | Y62.2 | ICD-10 during kidney dialysis or other perfusion |
| 35 | 233575001 | Intermittent haemodialysis stage 5 | 72 | Y84.1 | ICD-10 kidney dialysis |
| 36 | 398471000000102 | [V]Aftercare involving intermittent dialysis | 73 | Z49 | ICD-10 care involving dialysis |
| 37 | 366961000000102 | Renal transplant recipient stage 5 | 74 | Z49.0 | ICD-10 preparatory care for  dialysis |

**Table S13**: Atrial Fibrillation SNOMED and ICD-10 code list [12]

| # | Code | Term | # | Code | Term |
| --- | --- | --- | --- | --- | --- |
| 1 | 134377004 | Atrial fibrillation monitoring | 16 | 440028005 | Permanent atrial fibrillation (disorder) |
| 2 | 164889003 | ECG: atrial fibrillation | 17 | 440059007 | Persistent atrial fibrillation  (disorder) |
| 3 | 164890007 | ECG: atrial flutter | 18 | 49436004 | Atrial fibrillation |
| 4 | 195080001 | Atrial fibrillation and flutter | 19 | 5370000 | Atrial flutter |
| 5 | 233910005 | Lone atrial fibrillation (disorder) | 20 | 711411000000101 | Atrial fibrillation monitoring invitation (procedure) |
| 6 | 233911009 | Non-rheumatic atrial fib-  rillation | 21 | 716181000000109 | Atrial fibrillation monitoring third letter (procedure) |
| 7 | 720448006 | Typical atrial flutter (disorder) | 22 | 716721000000107 | Atrial fibrillation monitoring telephone invitation (procedure) |
| 8 | 282825002 | AF - Paroxysmal atrial fib-  rillation | 23 | 716981000000106 | Atrial fibrillation monitoring second letter (procedure) |
| 9 | 300996004 | Controlled atrial fibrillation (disorder) | 24 | 717011000000100 | Atrial fibrillation monitoring verbal invitation (procedure) |
| 10 | 312442005 | H/O: atrial fibrillation | 25 | 717221000000101 | Atrial fibrillation monitoring first letter (procedure) |
| 11 | 314208002 | Rapid atrial fibrillation  (disorder) | 26 | 248411000000105 | Atrial fibrillation annual review (regime/therapy) |
| 12 | 425615007 | Chronic atrial flutter (disorder) | 27 | 1596490100011910 | 0 Atypical atrial flutter (disorder) |
| 13 | 426749004 | Chronic atrial fibrillation  (disorder) | 28 | 1110851000000100 | Quality and Outcomes Framework atrial fibrillation quality indicator-related care invitation (procedure) |
| 14 | 427665004 | Paroxysmal atrial flutter  (disorder) | 29 | 120041000119109 | Atrial fibrillation with rapid ventricular response (disorder) |
| 15 | 428076002 | History of atrial flutter  (situation) | 30 | I48 | Atrial fibrillation and flutter |

**Table S14**: Migraine SNOMED and ICD-10 code list

|  | | # | Code | | Term | | | # | Code | | Term | |
| --- | --- | --- | --- | --- | --- | --- | --- | --- | --- | --- | --- | --- |
|  | | 1 | 4473006 | | Migraine with aura | | | 11 | 161481007 | | H/O: migraine | |
|  | | 2 | 23186000 | | Menstrual migraine | | | 12 | 193030005 | | Migraine variants | |
|  | | 3 | 37796009 | | Migraine | | | 13 | 193039006 | | Complicated migraine | |
|  | | 4 | 49605003 | | Ophthalmoplegic migraine | | | 14 | 230462002 | | Migraine with typical aura | |
|  | | 5 | 56097005 | | Migraine without aura | | | 15 | 230467008 | | Status migrainosus | |
|  | | 6 | 59292006 | | Hemiplegic migraine | | | 16 | 408381007 | | Migraine prophylaxis (procedure) | |
|  | | 7 | 75879005 | | Abdominal migraine | | | 17 | 608837004 | | History of migraine with aura | |
| 8 | 82639001 | | | Premenstrual tension syndrome | | 18 | 124001000119104 | | | Status migrainosus occurrent and due | | coto |
|  |  | | |  | |  |  | | | migraine without  (disorder) | | aura |
| 9 | 83351003 | | | Basilar migraine | | 19 | G43 | | | Migraine | |  |
| 10 | 95655001 | | | Ophthalmic migraine | |  |  | | |  | |  |

**Table S15**: Systemic lupus erythematosus SNOMED and ICD-10 code list

| # | Code | Term | # | Code | Term |
| --- | --- | --- | --- | --- | --- |
| 1 | 4676006 | SLE glomerulonephritis syndrome, WHO class II | 39 | 403492006 | Hypertrophic type discoid lupus erythematosus (disorder) |
| 2 | 7119001 | Cutaneous lupus  erythematosus | 40 | 403493001 | Rosaceous type discoid lupus erythematosus (disorder) |
| 3 | 11013005 | SLE glomerulonephritis syndrome, WHO class VI | 41 | 403494007 | Discoid lupus erythematosus of mucous membranes (dis-  order) |
| 4 | 15084002 | Lupus erythematosus profundus | 42 | 403495008 | Discoid lupus erythematosus of oral mucosa (disorder) |
| 5 | 36402006 | SLE glomerulonephritis syndrome, WHO class IV | 43 | 403496009 | Discoid lupus erythematosus of genital mucous membranes (disorder) |
| 6 | 52042003 | SLE glomerulonephritis syndrome, WHO class V | 44 | 403497000 | Discoid lupus erythematosus of scalp (disorder) |
| 7 | 54072008 | Nonbacterial verrucal endocardiosis | 45 | 403498005 | Discoid lupus erythematosus of face (disorder) |
| 8 | 55464009 | Systemic lupus erythematosus | 46 | 403499002 | Discoid lupus erythematosus of lip (disorder) |
| 9 | 68815009 | SLE glomerulonephritis syndrome | 47 | 403500006 | Discoid lupus erythematosus of hands (disorder) |
| 10 | 73286009 | SLE glomerulonephritis syndrome, WHO class I | 48 | 403501005 | Discoid lupus erythematosus of foot (disorder) |
| 11 | 76521009 | SLE glomerulonephritis syndrome, WHO class III | 49 | 403502003 | Disseminated discoid lupus erythematosus (disorder) |
| 12 | 79291003 | Discoid lupus erythematosus of eyelid | 50 | 403503008 | Lupus erythematosusassociated anetoderma  (disorder) |
| 13 | 95332009 | Rash of systemic lupus erythematosus | 51 | 403504002 | Lupus erythematosusassociated subcutaneous nodules (disorder) |
| 14 | 95644001 | Systemic lupus erythematosus encephalitis | 52 | 403505001 | Lupus erythematosusassociated calcinosis  (disorder) |
| 15 | 193178008 | Polyneuropathy in disseminated lupus erythematosus | 53 | 403506000 | Lupus erythematosusassociated papulonodular mucinosis (disorder) |
| 16 | 196138005 | Lung disease with systemic lupus erythematosus | 54 | 403507009 | Lupus erythematosusassociated hypermelanosis (disorder) |
| 17 | 200936003 | Lupus erythematosus | 55 | 403508004 | Lupus erythematosusassociated nailfold  telangiectasia (disorder) |

| 18 | 200937007 | Lupus erythematosus chronicus | 56 | 403510002 | Lupus erythematosusassociated urticarial  vasculitis (disorder) | |
| --- | --- | --- | --- | --- | --- | --- |
| 19 | 200938002 | Discoid lupus erythematosus | 57 | 403511003 | Lupus erythematosusassociated necrotizing  vasculitis (disorder) | |
| 20 | 200939005 | Lupus erythematosus  migrans | 58 | 403512005 | Lupus associated (disorder) | erythematosuspoikiloderma |
| 21 | 200940007 | Lupus erythematosus nodularis | 59 | 403513000 | Lupus associated (disorder) | erythematosusnail dystrophy |
| 22 | 200941006 | Lupus erythematosus  tumidus | 60 | 417303004 | Retinal vasculitis due to systemic lupus erythematosus  (disorder) | |
| 23 | 200942004 | Lupus erythematosus  unguium mutilans | 61 | 425951002 | Shrinking lung syndrome  (disorder) | |
| 24 | 200944003 | Lupus erythematosus NOS | 62 | 700269001 | Discoid lupus erythematosus of lower eyelid | |
| 25 | 201422008 | [X]Other local lupus erythematosus | 63 | 700334001 | Discoid lupus erythematosus of upper eyelid | |
| 26 | 238926009 | Lupus erythematosus and erythema multiforme-like syndrome | 64 | 430961000124105 | Cheilitis due to lupus erythematosus | |
| 27 | 238927000 | Discoid lupus erythematosus | 65 | 72181000119109 | Endocarditis due to systemic lupus erythematosus | |
| 28 | 238928005 | Chilblain lupus erythematosus | 66 | 295101000119105 | Nephropathy co-occurrent and due to systemic lupus erythematosus (disorder) | |
| 29 | 239887007 | Systemic lupus erythematosus with organ/system involvement | 67 | 308751000119106 | Glomerular disease due to systemic lupus erythematosus (disorder) | |
| 30 | 239889005 | Bullous systemic lupus erythematosus | 68 | 713225000 | Gingival disease cooccurrent and due to lupus erythematosus | |
| 31 | 239891002 | Subacute cutaneous lupus erythematosus | 69 | 295121000119101 | SLE (systemic lupus erythematosus) with nephrosis | |
| 32 | 239944008 | Lupus vasculitis | 70 | 295111000119108 | Systemic lupus erythematosus co-occurrent and due to nephrotic syndrome (disorder) | |
| 33 | 307755009 | Renal tubulo-interstitial disorder in systemic lupus erythematosus | 71 | 724767000 | Chorea co-occurrent and due to systemic lupus erythematosus (disorder) | |
| 34 | 309762007 | Systemic lupus erythematosus with pericarditis | 72 | 724781003 | Demyelination of central nervous system co-occurrent and due to systemic lupus erythematosus (disorder) | |
| 35 | 402865003 | Systemic lupus  erythematosus-associated  antiphospholipid syndrome  (disorder) | 73 | 732960002 | Secondary autoimmune haemolytic anaemia co-occurrent and due to systemic lupus  erythematosus | |
| 36 | 403489007 | Subacute cutaneous lupus erythematosus, annular/polycyclic type  (disorder) | 74 | L93 | Lupus erythematosus | |
| 37 | 403490003 | Subacute cutaneous lupus erythematosus, papulosquamous type (disorder) | 75 | M32 | Systemic lupus erythematosus | |
| 38 | 403491004 | Erythema multiformelike lupus erythematosus  (disorder) | 76 |  |  | |

**Table S16**: Rheumatoid arthritis SNOMED code list

| # | Code | | | Term | | | # | Code | | | Term | | |
| --- | --- | --- | --- | --- | --- | --- | --- | --- | --- | --- | --- | --- | --- |
| 1 | 398640008 | | | Rheumatoid with pneumoconiosis (disorder) | | | 36 | 400054000 | | | Rheumatoid with vasculitis  (disorder) | | |
| 2 | 54867000 | | | Rheumatoid with fibrosing  alveolitis (disorder) | | | 37 | 402433007 | | | Rheumatoid with nailfold and finger-pulp infarcts (disorder) | | |
| 3 | 398640008 | | | Rheumatoid with pneumoconiosis (disorder) | | | 38 | 427770001 | | | Rheumatoid of temporomandibular joint  (disorder) | | |
| 4 | 54867000 | | | Rheumatoid with fibrosing  alveolitis (disorder) | | | 39 | 429192004 | | | Rheumatoid of foot (disorder) | | |
| 5 | 398640008 | | | Rheumatoid with pneumoconiosis (disorder) | | | 40 | 69896004 | | | Rheumatoid (disorder) | | |
| 6 | 54867000 | | | Rheumatoid with fibrosing  alveolitis (disorder) | | | 41 | 735599007 | | | Rheumatoid with erosion of joint (disorder) | | |
| 7 | 201764007 | | | Rheumatoid of cervical spine  (disorder) | | | 42 | 735600005 | | | Rheumatoid without erosion  (disorder) | | |
| 8 | 201766009 | | | Rheumatoid of shoulder (disorder) | | | 43 | 7607008 | | | Rheumatoid with pericarditis (disorder) | | |
| 9 | 201767000 | | | Rheumatoid of sternoclavicular joint (disorder) | | | 44 | 781206002 | | | Rheumatoid of joint of spine  (disorder) | | |
| 10 | 201768005 | | | Rheumatoid of acromioclavicular joint (disorder) | | | 45 | 1149218009 | | | Rheumatoid with isolated  nailfold vasculitis (disorder) | | |
| 11 | 201769002 | | | Rheumatoid of elbow (disorder) | | | 46 | 1149219001 | | | Rheumatoid with systemic  vasculitis (disorder) | | |
| 12 | 201770001 | | | Rheumatoid of distal  radioulnar joint (disorder) | | | 47 | 129563009 | | | Rheumatoid with osteope-  riostitis (disorder) | | |
| 13 | 201771002 | | | Rheumatoid of wr  der) | | ist (disor- | 48 | 38877003 | | | Rheumatoid  (disorder) | | with aortitis |
| 14 | 201772009 | | | Rheumatoid of  pophalangeal (disorder) | | metacarjoint | 49 | 402432002 | | | Rheumatoid  trophilic (disorder) | | with neudermatitis |
| 15 | 201773004 | | | Rheumatoid of  interphalangeal finger (disorder) | | proximal joint of | 50 | 54867000 | | | Rheumatoid with fibrosing  alveolitis (disorder) | | |
| 16 | 201774005 | | | Rheumatoid of distal interphalangeal joint of finger  (disorder) | | | 51 | 59165007 | | | Rheumatoid with scleritis  (disorder) | | |
| 17 | 201775006 | | | Rheumatoid of hip (disor-  der) | | | 52 | 77522006 | | | Rheumatoid with episcleritis  (disorder) | | |
| 18 | 201776007 | | | Rheumatoid of sacroiliac  joint (disorder) | | | 53 | 1162677006 | | | Rheumatoid with rheumatoid lung disease (disorder) | | |
| 19 | 201777003 | | | Rheumatoid of knee (disorder) | | | 54 | 847261000000104 | | | Rheumatoid annual review  (regime/therapy) | | |
| 20 | 201778008 | | | Rheumatoid of tibiofibular joint (disorder) | | | 55 | 1073711000119100 | | | Rheumatoid of left hand  (disorder) | | |
|  | | 21 | 201779000 | | Rheumatoid of ankle (disorder) | | | | 56 | 1073721000119100 | | Rheumatoid of left hip (disorder) | |
|  | | 22 | 201780002 | | Rheumatoid of subtalar joint  (disorder) | | | | 57 | 1073731000119100 | | Rheumatoid of left knee (disorder) | |
|  | | 23 | 201781003 | | Rheumatoid of talonavicular joint (disorder) | | | | 58 | 1073741000119100 | | Rheumatoid of left shoulder  (disorder) | |
|  | | 24 | 201783000 | | Rheumatoid of first metatarsophalangeal joint (disorder) | | | | 59 | 1073791000119100 | | Rheumatoid of right hand  (disorder) | |
|  | | 25 | 201784006 | | Rheumatoid of lesser metatarsophalangeal joint (disorder) | | | | 60 | 1073801000119100 | | Rheumatoid of right hip  (disorder) | |
|  | | 26 | 201785007 | | Rheumatoid of interphalangeal joint of toe  (disorder) | | | | 61 | 1073811000119100 | | Rheumatoid of right knee  (disorder) | |
|  | | 27 | 239793008 | | Rheumatoid with organ / system involvement (disorder) | | | | 62 | 1073821000119100 | | Rheumatoid of right shoulder (disorder) | |
|  | | 28 | 239795001 | | Rheumatoid with multisystem involvement (disorder) | | | | 63 | 1073701000119100 | | Rheumatoid of left foot (disorder) | |
|  | | 29 | 239943002 | | Rheumatoid with necrotizing vasculitis (disorder) | | | | 64 | 1073751000119100 | | Rheumatoid of left wrist  (disorder) | |
|  | | 30 | 287006005 | | Rheumatoid of multiple  joints (disorder) | | | | 65 | 1073781000119100 | | Rheumatoid of right foot  (disorder) | |
|  | | 31 | 287007001 | | Rheumatoid of hand joint  (disorder) | | | | 66 | 1073831000119100 | | Rheumatoid of right wrist  (disorder) | |
|  | | 32 | 287008006 | | Rheumatoid of ankle and/or foot (disorder) | | | | 67 | 15686281000119100 | | Rheumatoid of bilateral feet  (disorder) | |
|  | | 33 | 28880005 | | Rheumatoid with carditis  (disorder) | | | | 68 | 15687201000119100 | | Rheumatoid of bilateral  knees (disorder) | |
|  | | 34 | 398640008 | | Rheumatoid with pneumoconiosis (disorder) | | | | 69 | 15687321000119100 | | Rheumatoid of bilateral hands (disorder) | |
|  | | 35 | 399923009 | | Rheumatoid with arteritis  (disorder) | | | |  |  | |  | |

**Table S17**: Severe mental illness SNOMED and ICD-10 code list

|  |  | **Schizophrenia** [13] | | |  |
| --- | --- | --- | --- | --- | --- |
| 1 | 111482003 | Subchronic schizophrenia with acute exacerbations  (disorder) | 34 | 270901009 | Schizoaffective disorder, mixed type (disorder) |
| 2 | 111483008 | Catatonic schizophrenia in remission (disorder) | 35 | 271428004 | Schizoaffective disorder, manic type (disorder) |

| 3 | 111484002 | Undifferentiated schizophrenia (disorder) | 36 | 27387000 | Subchronic disorganized schizophrenia (disorder) |
| --- | --- | --- | --- | --- | --- |
| 4 | 12939007 | Chronic disorganized  schizophrenia (disorder) | 37 | 29599000 | Chronic undifferentiated schizophrenia (disorder) |
| 5 | 14291003 | Subchronic disorganized schizophrenia with acute exacerbations (disorder) | 38 | 30336007 | Chronic residual schizophrenia with acute exacerbations (disorder) |
| 6 | 16990005 | Subchronic schizophrenia  (disorder) | 39 | 31373002 | Disorganized schizophrenia in remission (disorder) |
| 7 | 191526005 | Schizophrenic disorders (disorder) | 40 | 31658008 | Chronic paranoid  schizophrenia (disorder) |
| 8 | 191527001 | Simple schizophrenia (disorder) | 41 | 35218008 | Chronic disorganized schizophrenia with acute exacerbation (disorder) |
| 9 | 191530008 | Acute exacerbation of subchronic schizophrenia (disorder) | 42 | 35252006 | Disorganized schizophrenia  (disorder) |
| 10 | 191531007 | Acute exacerbation of chronic schizophrenia  (disorder) | 43 | 38368003 | Schizoaffective disorder, bipolar type (disorder) |
| 11 | 191538001 | Acute exacerbation of subchronic hebephrenic  schizophrenia (disorder) | 44 | 39610001 | Undifferentiated schizophrenia in remission (disorder) |
| 12 | 191539009 | Acute exacerbation of chronic hebephrenic  schizophrenia (disorder) | 45 | 416340002 | Late onset schizophrenia  (disorder) |
| 13 | 191542003 | Catatonic schizophrenia  (disorder) | 46 | 42868002 | Subchronic catatonic  schizophrenia (disorder) |
| 14 | 191547009 | Acute exacerbation of subchronic catatonic  schizophrenia (disorder) | 47 | 4926007 | Schizophrenia in remission  (disorder) |
| 15 | 191548004 | Acute exacerbation of chronic catatonic  schizophrenia (disorder) | 48 | 51133006 | Residual schizophrenia in remission (disorder) |
| 16 | 191554003 | Acute exacerbation of subchronic paranoid schizophrenia (disorder) | 49 | 58214004 | Schizophrenia (disorder) |
| 17 | 191555002 | Acute exacerbation of chronic paranoid  schizophrenia (disorder) | 50 | 63181006 | Paranoid schizophrenia in remission (disorder) |
| 18 | 191559008 | Latent schizophrenia (disorder) | 51 | 64905009 | Paranoid schizophrenia (disorder) |
| 19 | 191561004 | Subchronic latent  schizophrenia (disorder) | 52 | 68890003 | Schizoaffective disorder (disorder) |
| 20 | 191562006 | Chronic latent schizophrenia  (disorder) | 53 | 68995007 | Chronic catatonic  schizophrenia (disorder) |
| 21 | 191563001 | Acute exacerbation of subchronic latent schizophrenia (disorder) | 54 | 7025000 | Subchronic undifferentiated schizophrenia with acute exacerbations (disorder) |
| 22 | 191564007 | Acute exacerbation of chronic latent schizophrenia (disorder) | 55 | 70814008 | Subchronic residual schizophrenia with acute exacerbations (disorder) |
| 23 | 191565008 | Latent schizophrenia in  remission (disorder) | 56 | 71103003 | Chronic residual schizophrenia (disorder) |
| 24 | 191567000 | Schizoaffective schizophrenia  (disorder) | 57 | 76566000 | Subchronic residual  schizophrenia (disorder) |
| 25 | 191569002 | Subchronic schizoaffective schizophrenia (disorder) | 58 | 79204003 | Chronic undifferentiated schizophrenia with acute exacerbations (disorder) |
| 26 | 191570001 | Chronic schizoaffective schizophrenia (disorder) | 59 | 79866005 | Subchronic paranoid  schizophrenia (disorder) |
| 27 | 191571002 | Acute exacerbation of subchronic schizoaffective schizophrenia (disorder) | 60 | 83746006 | Chronic schizophrenia (disorder) |

| 28 | 191572009 | Acute exacerbation of chronic schizoaffective schizophrenia (disorder) | 61 | 84760002 | Schizoaffective disorder, depressive type (disorder) |
| --- | --- | --- | --- | --- | --- |
| 29 | 191574005 | Schizoaffective schizophrenia in remission (disorder) | 62 | 85861002 | Subchronic undifferentiated schizophrenia (disorder) |
| 30 | 191577003 | Cenesthopathic schizophrenia (disorder) | 63 | 88975006 | Schizophreniform disorder  (disorder) |
| 31 | 231485007 | Post-schizophrenic depression (disorder) | 64 | 1089481000000100 | Cataleptic schizophrenia  (disorder) |
| 32 | 26025008 | Residual schizophrenia (disorder) | 65 | F20 | Schizophrenia |
| 33 | 268617001 | Acute schizophrenic episode  (disorder) | 66 | F25 | Schizoaffective disorders |
| **Bipolar affective disease** [14] | | | | | |
| 1 | 111485001 | Mixed bipolar I disorder in full remission (disorder) | 38 | 28663008 | Severe manic bipolar I disorder with psychotic features (disorder) |
| 2 | 13313007 | Mild bipolar disorder (disorder) | 39 | 31446002 | Bipolar I disorder, most recent episode hypomanic  (disorder) |
| 3 | 13746004 | Bipolar disorder (disorder) | 40 | 371596008 | Bipolar I disorder (disorder) |
| 4 | 1499003 | Bipolar I disorder, single manic episode with postpartum onset (disorder) | 41 | 41836007 | Bipolar disorder in full  remission (disorder) |
| 5 | 162004 | Severe manic bipolar I disorder without psychotic features (disorder) | 42 | 4441000 | Severe bipolar disorder with psychotic features (disorder) |
| 6 | 16506000 | Mixed bipolar I disorder  (disorder) | 43 | 53049002 | Severe bipolar disorder without psychotic features (disorder) |
| 7 | 17782008 | Bipolar I disorder, most recent episode manic with catatonic features (disorder) | 44 | 53607008 | Depressed bipolar I disorder in remission (disorder) |
| 8 | 191583000 | Single manic episode, mild  (disorder) | 45 | 55516002 | Bipolar I disorder, most recent episode manic with postpartum onset (disorder) |
| 9 | 191584006 | Single manic episode, moderate (disorder) | 46 | 61403008 | Severe depressed bipolar I disorder without psychotic features (disorder) |
| 10 | 191586008 | Single manic episode, severe, with psychosis (disorder) | 47 | 63249007 | Manic bipolar I disorder in partial remission (disorder) |
| 11 | 191588009 | Single manic episode in full remission (disorder) | 48 | 765176007 | Psychosis and severe depression co-occurrent and due to bipolar affective disorder (disorder) |
| 12 | 191590005 | Recurrent manic episodes  (disorder) | 49 | 767631007 | Bipolar disorder, most recent episode depression  (disorder) |
| 13 | 191592002 | Recurrent manic episodes, mild (disorder) | 50 | 767632000 | Bipolar disorder, most recent episode manic  (disorder) |
| 14 | 191593007 | Recurrent manic episodes, moderate (disorder) | 51 | 767633005 | Bipolar affective disorder, most recent episode mixed  (disorder) |
| 15 | 191595000 | Recurrent manic episodes, severe, with psychosis (disorder) | 52 | 767635003 | Bipolar I disorder, most recent episode manic (disorder) |
| 16 | 191597008 | Recurrent manic episodes, in full remission (disorder) | 53 | 767636002 | Bipolar I disorder, most recent episode depression  (disorder) |
| 17 | 191618007 | Bipolar affective disorder, current episode manic (disorder) | 54 | 789061003 | Rapid cycling bipolar II disorder (disorder) |
| 18 | 191620005 | Bipolar affective disorder, currently manic, mild (disorder) | 55 | 79584002 | Moderate bipolar disorder  (disorder) |

| 19 | 191621009 | | | Bipolar affective disorder, currently manic, moderate  (disorder) | | 56 | 83225003 | | | Bipolar II disorder (disorder) | |
| --- | --- | --- | --- | --- | --- | --- | --- | --- | --- | --- | --- |
| 20 | 191623007 | | | Bipolar affective disorder, currently manic, severe, with psychosis (disorder) | | 57 | 85248005 | | | Bipolar disorder in remission  (disorder) | |
| 21 | 191625000 | | | Bipolar affective disorder, currently manic, in full  remission (disorder) | | 58 | 87950005 | | | Bipolar I disorder, single manic episode with catatonic features (disorder) | |
| 22 | 191627008 | | | Bipolar affective disorder, current episode depression (disorder) | | 59 | F30 | | | Manic episode | |
| 23 | 191629006 | | | Bipolar affective disorder, currently depressed, mild  (disorder) | | 60 | F31 | | | Bipolar affective disorder | |
| 24 | 191630001 | | | Bipolar affective disorder, currently depressed, moderate (disorder) | | 61 | 1089661000000100 | | | Mania with mood-congruent psychotic features (disorder) | |
| 25 | 191632009 | | | Bipolar affective disorder, currently depressed, severe, with psychosis (disorder) | | 62 | 1089671000000100 | | | Mania with moodincongruent psychotic  features (disorder) | |
| 26 | 191634005 | | | Bipolar affective disorder, currently depressed, in full remission (disorder) | | 63 | 1089681000000100 | | | Mania with psychotic features (disorder) | |
| 27 | 191636007 | | | Mixed bipolar affective disorder (disorder) | | 64 | 16238741000119100 | | | Bipolar disorder caused by drug (disorder) | |
| 28 | 191638008 | | | Mixed bipolar affective disorder, mild (disorder) | | 65 | 467361000000105 | | | [X] Manic-depressive psychosis, depressed type without psychotic symptoms (disorder) | |
| 29 | 191639000 | | | Mixed bipolar affective disorder, moderate (disorder) | | 66 | 760721000000109 | | | Mixed bipolar affective disorder, in partial remission (disorder) | |
| 30 | 191641004 | | | Mixed bipolar affective disorder, severe, with psychosis (disorder) | | 67 | 764591000000108 | | | Mixed bipolar affective disorder, severe (disorder) | |
| 31 | 191643001 | | | Mixed bipolar affective disorder, in full remission (disorder) | | 68 | 764621000000106 | | | Recurrent manic episodes, severe (disorder) | |
| 32 | 191658009 | | | Atypical manic disorder (disorder) | | 69 | 764641000000104 | | | Single manic episode, severe  (disorder) | |
| 33 | 192362008 | | | Bipolar affective disorder, current episode mixed (disorder) | | 70 | 764671000000105 | | | Recurrent manic episodes, in partial remission (disorder) | |
| 34 | 231494001 | | | Mania (disorder) | | 71 | 764681000000107 | | | Recurrent manic episodes, in remission (disorder) | |
| 35 | 231495000 | | | Manic stupor (disorder) | | 72 | 764731000000103 | | | Single manic episode in partial remission (disorder) | |
| 36 | 231496004 | | | Hypomania (disorder) | | 73 | 764741000000107 | | | Single manic episode in  remission (disorder) | |
| 37 | 268619003 | | | Manic disorder, single  episode (disorder) | |  |  | | |  | |
| **Psychosis** | | | | | | | | | | | |
| 1 | 5464005 | | | Brief reactive psychosis (disorder) | | 15 | 71961003 | | | Childhood disintegrative disorder (disorder) | |
| 2 | 268622001 | | | Chronic paranoid psychosis  (disorder) | | 16 | 69322001 | | | Psychotic disorder (disorder) | |
| 3 | 191525009 | | | Chronic paranoid psychosis  (disorder) | | 17 | 231487004 | | | Persistent delusional disorder (disorder) | |
| 4 | 712824002 | | | Acute polymorphic psychotic disorder without symptoms of schizophrenia  (disorder) | | 18 | 278853003 | | | Acute schizophrenia-like psychotic disorder (disorder) | |
| 5 | 191613003 | | | Recurrent major depressive episodes, severe, with psychosis (disorder) | | 19 | 48500005 | | | Delusional disorder (disorder) | |
|  | | 6 | 61831009 | | Induced psychotic disorder  (disorder) | | | 20 | 231437006 | | Reactive psychoses (disorder) |
|  | | 7 | 231489001 | | Acute transient psychotic disorder (disorder) | | | 21 | 31027006 | | Schizotypal personality disorder (disorder) |
|  | | 8 | 191676002 | | Reactive depressive  psychosis (disorder) | | | 22 | 191667009 | | Paranoid disorder (disorder) |
|  | | 9 | 274953007 | | Acute polymorphic psychotic disorder (disorder) | | | 23 | 191677006 | | Acute hysterical psychosis  (disorder) |
|  | | 10 | 712850003 | | Acute polymorphic psychotic disorder cooccurrent with symptoms of schizophrenia (disorder) | | | 24 | 231536004 | | Atypical autism (disorder) |
|  | | 11 | 441704009 | | Affective psychosis (disorder) | | | 25 | 307417003 | | Cycloid psychosis (disorder) |
|  | | 12 | 274952002 | | Borderline schizophrenia  (disorder) | | | 26 | 231449007 | | Epileptic psychosis (disorder) |
|  | | 13 | 35919005 | | Pervasive developmental disorder (disorder) | | | 27 | 417601000000102 | | [X]Schizophrenia, schizotypal and delusional disorders (disorder) |
|  | | 14 | 191680007 | | Pervasive developmental disorder (disorder) | | |  |  | |  |
